# Exploring the causal role of the human gut microbiome in breast cancer risk

**DOI:** 10.1101/2025.02.12.25322118

**Authors:** Grace L. Edmunds, Tim Robinson, James Yarmolinsky, Matthew A Lee, Bryony Hayes, David A Hughes, Lara Gibbs, Jeremy P. Braybrooke, Jeroen Raes, Nicholas J. Timpson, Kaitlin H. Wade

## Abstract

The relationship between the human gut microbiome and breast cancer (BC) risk is uncertain. We used Mendelian randomization (MR) and multivariable linear regression to investigate the causal relationship between the human gut microbiome and BC risk (overall and subtype-specific) and interrogate the role of circulating metabolites as potential mediators. Summary-level data from genome-wide association studies of gut microbial traits (N=3,890), circulating metabolites (N=7,824), overall (N=247,175) and subtype-specific (N=200,898) BC risk, including triple negative BC (TNBC; N=120,493), alongside individual-level data from the Flemish Gut Flora Project (N=2,242). Sensitivity analyses were applied to examine the robustness of causal estimates. Initial two-sample MR analyses suggested that the presence of bacteria within the *Ruminococcus* genus increased overall BC risk, that higher abundances of bacteria within the *Parabacteroides* genus decreased and presence of unclassified bacteria within the *Bacteroidales* order increased TNBC risk, and that several circulating metabolites altered BC risk (e.g., homostachydrine with overall BC and Luminal A BC, and betaine with TNBC). Whilst, in multivariable linear regression analyses, several of these metabolites were also associated with the microbial traits that influenced BC risk, importantly, given directional inconsistency of associations across analyses of microbial traits, metabolites and BC coupled with sensitivity analyses implying violations of MR assumptions, associations were unlikely to reflect causality nor a causal pathway between the gut microbiome, metabolites, and BC risk.

## INTRODUCTION

Breast cancer (BC) is the most common malignancy worldwide, with >2.2 million new cases and 684,000 deaths reported in 2020 (1). Although the mortality rate of BC is decreasing (2), greater understanding of the modifiable causal factors of BC risk is required to inform prevention. There has been increasing acknowledgement of the potential role of the human gut microbiome – a dynamic and complex system of approximately 100 trillion microorganisms including bacteria, eukaryotes, viruses and archaea – in cancer risk (3,4). Understanding of the human gut microbiome and its potential effects on cancer has rapidly increased (5–7). For example, there have been multiple direct mechanisms proposed that may link the gut microbiome to colorectal cancer, perhaps the most studied cancer type in this context, including promotion of a localised inflammatory response (8) and the generation of reactive oxygen species and intestinal stem cell proliferation (9). For most cancers that are distal to the gut, such as BC, causal relationships may be mediated by systemic mechanisms such as metabolism (and subsequent adiposity and insulin resistance (10)), inflammation (11) and immune function (6) – all of which are key hallmarks of cancer initiation and progression (12). There is evidence that bacteria residing in the gut can produce and alter levels of circulating metabolites and proteins that have been linked to the risk of one or more subtypes of BC, including hormones (e.g., oestrogen), cytokines (13,14), and levels of numerous circulating proteins (15).

Most studies investigating relationships between the gut microbiome and BC risk have been of case-control or cross-sectional design, often involving relatively low numbers of participants (range of cases and controls: 37-148) (16–19). However, conventional observational studies may provide inaccurate estimates of the relationship between the gut microbiome and cancer as they are generally prone to, predominantly, confounding and reverse causation. Mendelian randomization (MR) is a form of instrumental variable analysis that uses human germline genetic variation, most commonly single nucleotide polymorphisms (SNPs), to reduce some of the potential limitations of conventional observational studies (20). As germline genetic variants are inherited randomly and fixed at gametogenesis, analyses using these variants as instruments in an instrumental variable analysis should be both largely independent from confounding and not influenced by reverse causation (21). Importantly, the comparison of estimates obtained from MR and conventional observational studies, which have differing limitations and key sources of bias, can provide improved causal inference (22). Given the growing evidence for host genetic contributions to gut microbiome composition (23), there have been a number of recent studies that have used MR to investigate causal links between the gut microbiome, or gut microbiome-associated metabolites, and various traits. However, as the gut microbiome is influenced by the host’s environment and genetics, both of which influence host phenotypic development, MR analyses assessing the downstream impact of the microbiome and derived causal estimates of these relationships require careful examination and interpretation.

Here, we firstly used summary-level data from genome-wide association studies (GWASs) and two-sample MR analyses to interrogate whether there was a causal impact of the human gut microbiome and circulating metabolites on both overall and subtype-specific BC risk. Secondly, multivariable linear regression using individual-level data from the Flemish Gut Flora Project (FGFP) was then conducted to assess the relationship between the gut microbiome and circulating metabolites, results of which were compared to MR analyses. We then assessed the directional consistency of estimates across all analyses to provide insight into the possible causal pathways between gut microbial features, metabolites and BC risk.

## METHODS

We applied two-sample MR using summary-level data to estimate the causal effects of the gut microbiome and circulating metabolites on BC risk. We then undertook multivariable linear regression and two-sample MR analyses to estimate the association between the gut microbiome and circulating metabolites. Finally, to assess whether metabolites could mediate the association between microbial traits and BC risk, we explored the consistency of effect estimates across the associations between the gut microbiome, circulating metabolites and BC that persisted across all analyses (Figure 1). For MR analyses, sensitivity analyses were used to examine the robustness of causal estimates to violations of MR assumptions. In all analyses, we focused on both overall and subtype-specific BC outcomes. This study and all methods have been conducted in line with the Strengthening the Reporting of Observational Studies in Epidemiology MR (STROBE-MR) reporting guidelines (Supplementary Note) for MR studies (24).

**Figure 1.**
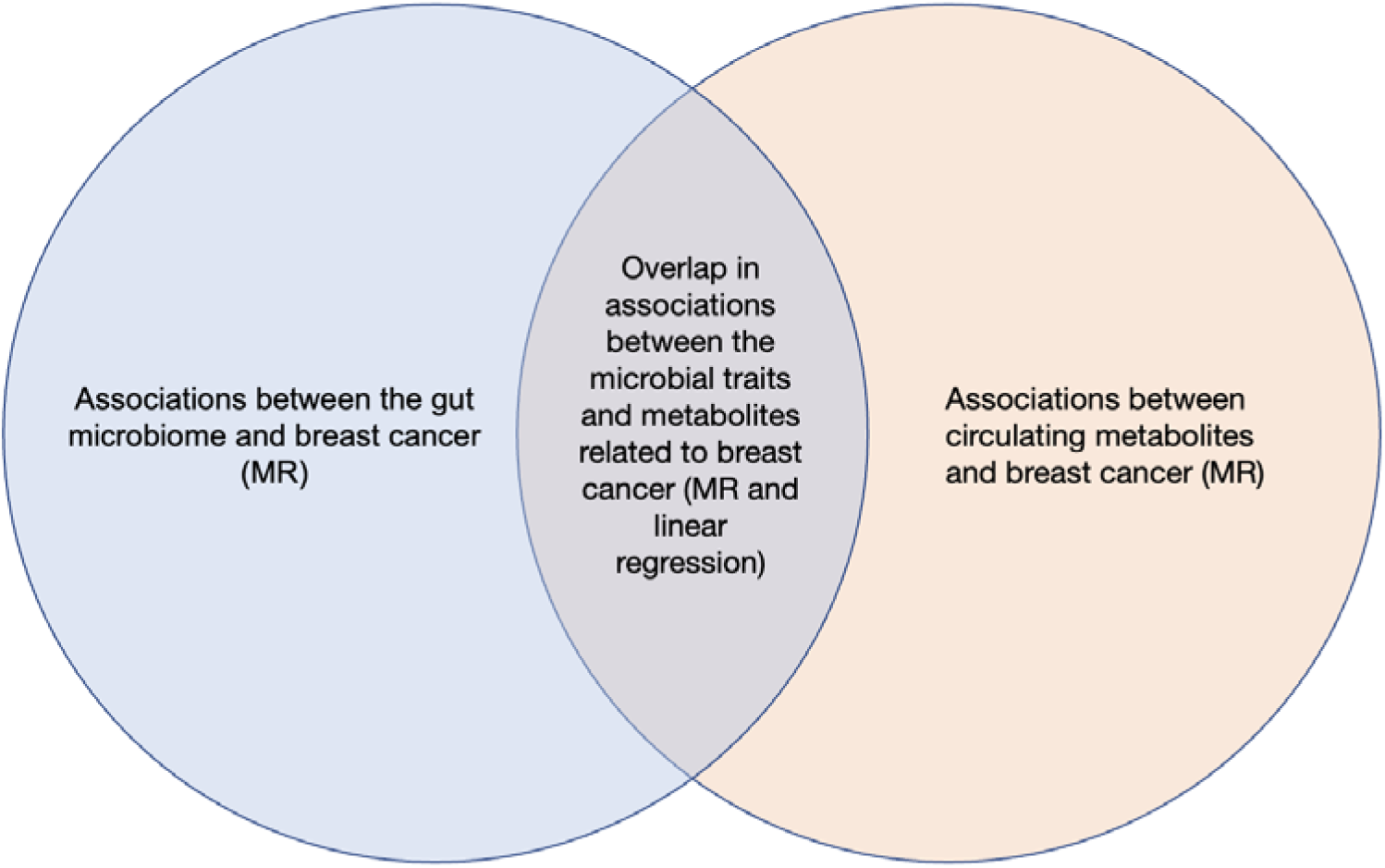
Proposed analysis structure. MR = Mendelian randomization. We tested for associations between the gut microbiome and breast cancer (BC) risk, and circulating metabolites with BC risk using Mendelian randomization (MR) (blue circle and orange circle, respectively). Then, MR and multivariable linear regression (middle section) were used to investigate associations between microbial traits and metabolites, where overlap in all analyses between the gut microbiome, metabolites and BC risk was then qualitatively investigated.

### Data Sources

All studies that contributed data to this analysis had relevant ethical approval and participants had provided informed consent. All data were accessed between June 2020 and August 2023. An overview of the analysis plan, data available and sample sizes of studies is provided in Figure 2.

**Figure 2.**
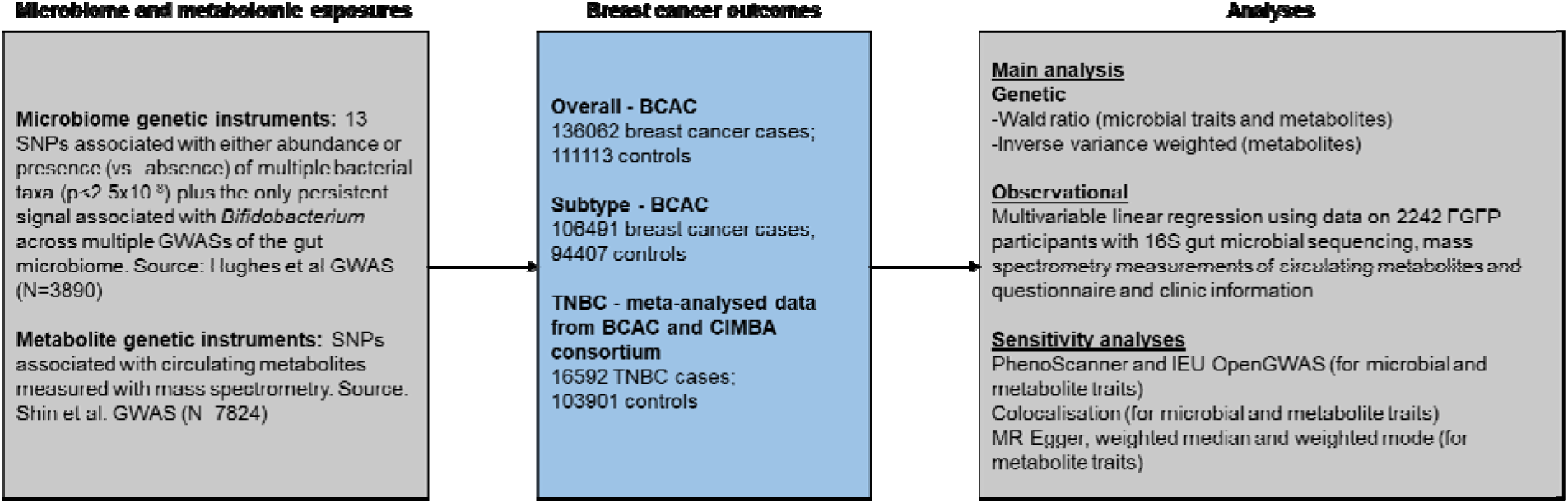
Genetic instruments, data sources used in the current analysis and analysis plan. BCAC = Breast Cancer Association Consortium; CIMBA = Consortium of Investigators of Modifiers of BRCA1/2; FGFP = Flemish Gut Flora Project; GWAS = genome-wide association study; HER2 = human epidermal growth factor 2 receptor; IVW = inverse variance weighted; MR = Mendelian randomization; SNP = single nucleotide polymorphism; TNBC = triple negative breast cancer. The current analysis used Mendelian randomization (MR) and multivariable linear regression to identify associations between the human gut microbiome, circulating metabolites, and breast cancer (BC) risk. Genetic instruments used to proxy variation in the human gut microbiome were generated from summary-level data from a genome-wide association study (GWAS) meta-analysis of the human gut microbiome from the Flemish Gut Flora Project (FGFP) and two German cohorts (N=3,890). Summary statistics from a GWAS of over 400 blood metabolite traits in a population of up to 7,824 adults of European descent were used in MR analyses to assess the effect of the gut microbiome on circulating metabolites and to assess the effect of circulating metabolites on BC risk. Individual-level data from FGFP were used for multivariable linear regression analyses to assess the association between gut microbial traits and circulating metabolites. Summary-level data were obtained from GWASs of overall and subtype-specific BC from the most recent Breast Cancer Association Consortium (BCAC) meta-analysis, where BC subtypes included Luminal A-like, Luminal B-like, human epidermal growth factor 2 receptor (HER2) negative and HER2 positive. Additional summary-level data were obtained from a triple negative BC (TNBC) GWAS meta-analysis and Breast Cancer Risk Allele (BRCA) 1 carriers from the Consortium of Investigators of Modifiers of BRCA1/2 (CIMBA) consortium, who were assumed to have TNBC. For exposures associated with a single associated SNP, MR estimates were generated per SNP using the Wald ratio method. For analyses using instruments with >1 SNP, the inverse variance weighted (IVW) method was used to estimate causal effects between each exposure and outcome. Individual multiple correction thresholds were applied to each analysis.

#### Breast cancer

Summary-level data were obtained from GWASs of overall and subtype-specific BC from the Breast Cancer Association Consortium (BCAC) meta-analysis (25). Women were included if they had a diagnosis of a primary invasive BC without known metastases. BC subtypes included Luminal A-like, Luminal B-like, human epidermal growth factor 2 (HER2) negative, HER2 positive, and triple negative BC (TNBC). Overall BC data from BCAC comprised a meta-analysis of several GWASs which used iCOGs genotyping array (38,349 cases and 37,818 controls), Oncoarray genotyping array (80,125 cases and 58,385 controls) and other studies which used bespoke arrays (17,588 cases and 14,910 controls), resulting in a total meta-analysis sample size of 136,062 cases and 111,113 controls (25). The GWAS of subtypes conducted by BCAC comprised 34,783 cases and 37,628 controls from iCOGs, and 71,708 cases and 56,779 controls from Oncoarray, for which tumour subtype data was available. The analysis then used a two-stage polytomous model to determine SNP-based associations with risk of each BC subtype across the total meta-analysis. This meant that the sample size for each subtype was not available; therefore, the total subtype GWAS case/control numbers were used in MR analyses as described previously(26) and according to personal communications with the authors (106,491 cases and 94407 controls)(25). Additional summary-level data were obtained from a TNBC meta-analysis of 16,592 cases and 103,901 controls, comprised of the BCAC TNBC cohort, and Breast Cancer Risk Allele (*BRCA*) 1 carriers from the Consortium of Investigators of Modifiers of BRCA1/2 (CIMBA) consortium, who were assumed to have TNBC(25). BCAC data were accessed in June 2023.

Participants in BCAC were enrolled from 1970 until 2020 and participants in CIMBA were enrolled between 1973 and 2014 with all individuals being of European ancestries. The median (interquartile range) ages of participants in the BCAC data contributing to both overall and BC subtype-specific analyses were 57 (49–65) years old in cases and 57 (48–66) years old in controls. Participants were enrolled from 28 countries (Australia, Austria, Belarus, Belgium, Canada, Czech Republic, Denmark, Finland, Germany, Greece, Hungary, Ireland, Israel, Italy, Latvia, Lithuania, Macedonia, Netherlands, New Zealand, Norway, Poland, Portugal, Russia, South Africa, Spain, Sweden, UK, USA). The *BRCA1* mutation carriers in the CIMBA cohort had a median age (interquartile range) of 40 (34–46) years in BC cases and 40 (32–49) years in controls. They came from 24 countries (Australia, Austria, Belgium, Canada, Czech Republic, Denmark, Finland, Germany, Greece, Hungary, Ireland, Israel, Italy, Latvia, Lithuania, Netherlands, Poland, Portugal, Russia, South Africa, Spain, Sweden, UK, USA). There was no overlap of participants between the BCAC and CIMBA data.

#### Gut microbiome

Summary-level data were obtained from a GWAS meta-analysis of the human gut microbiome from FGFP (N=2,223) and two German cohorts (FoCus, N=950 and PopGen, N=717)(27). The mean age in years of participants (range) was 52.3 (10–88), or 50.5 (10–88) for females and 54.9 (12–87) for males within FGFP. The mean age in years of participants (range) was 51.4 (18–83) within FoCus and 60.8 (25–83) within PopGen. The percentage of participants who were female in FGFP, FoCus and PopGen were 59.6%, 63.1% and 42.7%, respectively, with all participants of European ancestry. Faecal 16S ribosomal RNA (rRNA) sequencing was used to provide taxonomical classifications of all 16S sequences in each sample down to the genus level. Some 16S sequences could not be confidently assigned into all taxonomic levels and, beyond the level of confident assignment, they are assigned to an “unclassified” group. Taxa that had substantial zero-inflated distributions were modelled in a two-step process within the GWAS, which included a binary (presence vs. absence) analysis and a zero-truncated rank normal transformed abundance analysis. All other taxa were treated as abundance phenotypes and rank normal transformed.

GWAS analyses of host genotypic data with these microbial traits provided evidence for associations between 13 SNPs and either abundance or presence (vs. absence) of 13 microbial traits, meeting a genome-wide meta-analysis p-value threshold (P<2.5×10^-08^). For the current MR analyses, we used all 13 SNPs associated with microbial traits (each SNP being associated with one microbial trait), along with the only persistent signal across all previous GWASs of the gut microbiome – the rs4988235 SNP in the *MCM6*/*LCT* locus associated with *Bifidobacterium* (which reached a p-value of 1.34×10^-06^ in the Hughes *et al*. GWAS) (Supplementary Table S1)(23,28–32). For abundance phenotypes, the GWAS effect estimates represent normalized standard deviation (SD) units of change in microbial trait relative abundance with each effect allele and, for binary phenotypes, the effect estimates represent the log odds ratio (OR) of presence vs. absence of each microbial trait with each effect allele.

#### Circulating metabolites

Two separate data sources were used for the MR and multivariable regression analyses comprising metabolite data. Firstly, summary-level data were obtained from a GWAS (published by Shin et al.) of over 400 blood metabolite traits, measured using mass spectrometry (MS), in a population of up to 7,824 adults of European descent (33), and were used in MR analyses to assess the effect of circulating metabolites on BC risk and the effect of the gut microbiome on circulating metabolites. Data for the 299 SNPs that were independently and strongly associated (P<5×10^-08^) with MS-derived metabolites in this GWAS were obtained (i.e., those presented in their Supplementary Table 2). Metabolite values were transformed such that GWAS effect estimates reflected the log_10_ unit change in the concentration in each metabolite per effect allele. Secondly, individual-level metabolite data (N=2,906) for the 2,228 FGFP participants, who also had 16S microbiome data (N=2,248) and epidemiological data (N=2,959) available, were used for multivariable linear regression analyses to assess the association between gut microbial traits and circulating metabolites. Experimentally, blood serum samples from 3,005 FGFP participants were processed by MS on the Metabolon (Durham, NC, USA) platform, following Metabolon’s standard data quality control measures(34). For certain metabolites, multiple metabolites may share identical backbones and biochemical names whilst being unique metabolites (as indicated by compid). In total, there were 1,049 metabolites available for analysis with each of the 14 microbial traits.

For these multivariable regression analyses, we included seven covariables: sex, age, body mass, smoking status, average alcohol consumption in the week prior to sampling, vegetarian status, and monthly household income. Sex (binary; N=2,880), age (years; N=2,948) and body mass index (derived from weight divided by the square of height, kg/m^2^; N=2,945) were self-reported as opposed to taken from medical records. Smoking was a continuous trait (coded as 0 for a never smoker, 1 for a former smoker, and 2 for a current smoker; N=2,635), alcohol consumption was a continuous trait (coded from 0 = no alcohol, to 7 = more than 10 units; N=2,635), vegetarian status was a self-described binary trait (N=2,629), and monthly household income was a continuous trait (coded from 1 = less than 750 Euros, to 7 = greater than 3500 Euros; where the 960 individuals who did not know or preferred not to answer were coded as NA; N=2,224). Individuals with a mismatch in self-reported and health record sex and age were removed from analyses (N=75), and all individuals less than 18 years of age were removed from analyses (N=39).

### Statistical Analyses

#### Mendelian randomization

The MR framework makes three assumptions: firstly, that genetic instruments are strongly associated with the exposure (the “relevance” assumption); secondly, that there is no confounding between the genetic instruments and the outcome (the “independence” or “exchangeability” assumption); and lastly, that genetic instruments have no effect on the outcome except by acting through the exposure (the “exclusion restriction” assumption). To assess the causal effect of each microbial trait and metabolite on BC risk, SNPs used to proxy the exposure of interest were extracted from the relevant GWAS and harmonized to ensure that all GWAS beta-coefficients were in reference to the effect allele that increased the exposure. If any exposure-related SNP was not available in the outcome GWAS, we identified proxies using LD-Link with data of European ancestries (35) that were in high (R^2^>0.9) linkage disequilibrium (LD) with the primary SNP and, if available, extracted data for these proxy SNPs from the outcome GWAS instead. Where 1 SNP was available, estimates were generated per SNP using the Wald ratio, where *β_IV =_ β_ZY/_β_ZX_*, where *β_ZY_* is the effect estimate of association between the SNP and the outcome and *β_ZX_* is the effect estimate of association between the SNP and the exposure. For analyses using instruments with >1 SNP, the inverse variance weighted (IVW) method with multiplicative random effects was used to estimate causal effect between each exposure and outcome, which combines Wald ratio estimates for each SNP, weighting each estimate by the inverse of the variance of the SNP-outcome association.

For all analyses, the strength of SNP-exposure association was assessed using an F-statistic for instrument strength and an R^2^ value was used to calculate the variance in each exposure explained by associated SNP(s). For continuous traits, R^2^ was calculated using the formula:

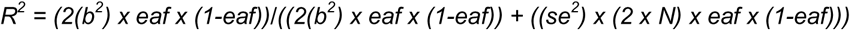

where b is beta of the SNP-exposure association, eaf is effect allele frequency of the SNP, se is standard error of the SNP-exposure association and N is sample size of the SNP-exposure GWAS. F-statistics were calculated using the formula:

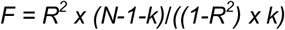

where k is the number of SNPs in the instrument and N is sample size of the SNP-exposure GWAS.

For binary traits, R was calculated using the ‘get_r_from_lor’ function in the TwoSampleMR package (the result of which was then squared to obtain the R^2^), and F-statistics were then calculated as above. For binary microbial traits specifically, the R^2^ calculation used the prevalence estimates of microbial trait presence derived from FGFP(27). For multiple SNP instruments, R^2^ was calculated by taking the sum of the R^2^ value for each SNP being used as an instrument, and the F-statistic was the mean of the F-statistics across all SNPs being used as an instrument.

For MR analyses of microbial traits on BC risk, the Wald ratio estimate was exponentially transformed to give the odds ratio (OR) per normalized SD change in each continuous microbial trait abundance or with an approximate doubling of the genetic liability of presence (vs. absence) of each binary microbial trait. Similarly, for MR analyses of metabolites on BC risk, estimates were exponentially transformed to represent the OR in BC risk per log_10_ higher concentration of each metabolite. Finally, Wald ratio estimates of the association between microbial traits and metabolites represented the log_10_ unit change in each metabolite with the same interpreted changes in each continuous (normalized SD) abundance and binary (approximate doubling in the genetic liability) presence vs. absence microbial trait, as described above.

For all analyses, P-values were interpreted as continuous indicators of evidence strength and conclusions were drawn based on effect sizes and their precision. Given the much greater number of metabolites being assessed in this study, a Bonferroni-corrected p-value threshold accounting for the approximate number of independent metabolites was applied to MR and linear regression analyses (see "*Multivariable linear regression*" section below) assessing possible causal relationships between metabolites and BC risk. In the MR analyses, as we were using summary-level data (and given that full summary statistics were not available for all metabolites at the time of analyses), we were unable to accurately determine the number of truly independent metabolites analysed in the Shin *et al*. GWAS; therefore, we used a Bonferroni-corrected p-value for the analyses of metabolites with overall and subtype-specific BC depending on the total number of metabolites that were available for MR analyses (i.e., those that had associated SNPs).

#### Multivariable linear regression

Within FGFP, we used multivariable linear regression to estimate associations between microbial traits and circulating metabolites. Our analyses were adjusted for sex, age, body mass index, smoking status (categorised as “never”, “ever” and “current”), alcohol consumption in the week prior to sampling (defined as the weekly average), vegetarian status (as a proxy for overall dietary behaviour, which was a binary trait defined as “vegetarian” vs. “other”), and income status (defined as monthly household income) as possible confounders of this relationship. Given the correlated nature of the metabolite data, a tree-cutting algorithm on a dendrogram derived from a Spearman’s correlation distance (1-absolute(rho)) matrix (https://github.com/hughesevoanth/iPVs; cut-height = 0.4) was used to estimate the number of effectively independent variables (N=542 metabolites) within the data. Having determined the number of independent observations using individual-level data, we therefore defined a Bonferroni p-value correction for the multivariable linear regression analyses accounting for the number of microbial traits for which there was evidence for an effect on BC risk (n) and 542 independent metabolites as 0.05/(n*542). Both microbial traits and metabolites were rank normal transformed, randomly splitting tied values, prior to linear modelling in R with the function lm() from the stats package. After accounting for complete data across all linear model variables, the largest sample size in any linear model between the microbial traits and metabolites was 1,590. Results from these analyses represent the rank normalized SD unit change in each metabolite with each normalized SD unit change in abundance in each continuous gut microbial trait or with presence (vs. absence) of each binary microbial trait. Although multiple testing thresholds were set for each analysis (described above), the magnitude and precision of effect estimates, together with consistency in the direction across multiple tests (and sensitivity analyses), were considered the main basis of scientific conclusions.

#### Identifying overlap between the gut microbiome, metabolites and breast cancer risk

To provide qualitative evidence of a possible biologically plausible mediating role of a circulating metabolite in the relationship between a gut microbial trait and BC risk, we compared the consistency of direction between (i) MR-derived estimates of the effect of that gut microbial trait on BC risk, (ii) MR-derived estimates of the effect of a circulating metabolite concentration on BC risk and (iii) the estimates linking the gut microbial trait with the circulating metabolite concentration using multivariable linear regression and MR analyses. If there was evidence that a higher abundance (or presence vs. absence) of a microbial trait increased BC risk (i.e., a positive association), we looked for either consistently positive or consistently negative associations across both analyses assessing the associations between (i) that microbial trait and circulating metabolite and (ii) that circulating metabolite and BC risk. If there was evidence that a higher abundance (or presence vs. absence) of a microbial trait reduced the risk of BC risk (i.e., a negative association), we looked for the following two possible conditions to be satisfied: (i) a positive association between that microbial trait and circulating metabolite and a negative association between that circulating metabolite and BC risk or (ii) a negative association between that microbial trait and a circulating metabolite and a positive association between that metabolite and BC risk.

### Sensitivity analyses

To examine the robustness of causal estimates to violations of MR assumptions (predominantly to assess the presence of horizontal pleiotropy), the following sensitivity analyses were undertaken. For single-SNP instruments, sensitivity analyses were based on manually searching both PhenoScanner (36) and the IEU OpenGWAS (37) (i.e., due to the unique GWAS summary statistics across these resources, despite a high level of overlap) to determine if SNPs used as instruments were associated with other traits that could also cause changes in the outcome independently of the exposure of interest. P-value thresholds indicating evidence for an association between exposure-related SNPs and traits were defined to account for the number of traits present within each database, where a 10% Bonferroni correction was applied due to the likelihood that traits could be correlated within those databases. For PhenoScanner, the p-value threshold was therefore set at 0.0001 (i.e., accounting for the largest number of results that the online tool provides; N=1,000) and, for the IEU OpenGWAS, a p-value threshold was set at 2.53×10^-06^ to correct for the 39,603 GWAS datasets within the database at the time of running analyses (2^nd^ December 2022). For these analyses, we present all results in the Supplementary tables that reached a p-value of 0.001 in the IEU OpenGWAS and across the five databases in PhenoScanner (i.e., diseases and traits, gene expression, proteins, metabolites and epigenetic markers), allowing proxies from European ancestries data (defined by an R^2^ of 0.8 on human genome build 37), highlighting those that reached our ≥ defined p-value thresholds accounting for multiple testing. If results are not presented for any of the five databases within PhenoScanner, there were no results that reached a p-value of 0.001 in that search.

Given the limited number of SNPs associated with each microbial trait and some metabolites, we also used colocalisation to examine whether the microbial traits or metabolites and BC risk were likely to share a causal variant or whether both traits were influenced by distinct causal variants which may be correlated due to LD (38). While not sufficient, shared causal variants between two traits are necessary for them to be causally related. The coloc R package (version 2.0) was used, which uses approximate Bayes factor computation to generate posterior probabilities that associations between two traits represent each of the following configurations: (H_0_) neither trait has a genetic association in the region, (H_1_) only the first trait (i.e., a microbial trait or metabolite) has a genetic association in the region, (H_2_) only the second trait (i.e., BC) has a genetic association in the region, (H_3_) both traits are associated but have different causal variants and (H_4_) both traits are associated and share a single causal variant. This colocalisation method assumes that there is a single causal variant in the region (39). Prior probabilities were set as the default (i.e., at H_1_ = 10^-4^, H_2_ = 10^-4^, and H_3_ = 10^-5^) and the posterior probability used was also the default (i.e., >0.80 providing strong evidence for a particular hypothesis), with additional comparison between hypotheses. Where MR analyses provided evidence of a causal effect of any microbial trait or metabolite on BC risk, which was instrumented by a single SNP, colocalisation analysis was either performed using FGFP data alone (i.e., where we had access to genome-wide summary-level data for each microbial trait, as this was not available across both FoCus and PopGen due to the methodology of the GWAS published by Hughes *et al.* (27)) or the Shin *et al.*(33) summary statistics (i.e., for metabolites). This was undertaken by generating windows ±1Mb from the top SNP used to instrument the microbial trait or metabolite exposure and extracting the summary statistics from both the exposure and outcome GWAS for those SNPs. Regional association plots were generated to visualise genetic colocalisation using the LocusCompareR package (40).

Lastly, for analyses with more than 3 associated SNPs, heterogeneity of estimates between these SNPs was calculated using a Cochran’s Q statistic and the likelihood of horizontal pleiotropy was assessed through the estimation of the MR-Egger test intercept and the comparison of the causal effect estimates obtained from the weighted median, weighted mode and MR-Egger estimators. In all instances, the directional consistency between these analyses and the IVW method was assessed (41–43). Leave-one-out analyses were also used to determine whether any individual SNP from the multi-SNP instrument was driving the MR estimate. In all MR analyses, the sources of exposure and outcome data were independent and homogenous (i.e., in terms of ancestry).

All analyses were conducted using R Studio version 4.0.2 and the TwoSampleMR (version 0.5.6) packages. Code used to execute analyses is available on Github (https://github.com/ge8793/Biome_Public).

## RESULTS

### Gut microbial traits and breast cancer risk

Of the 14 microbial traits that had associated genetic variants in the Hughes *et al*. GWAS (27), 13 SNPs individually associated with 13 microbial traits were available in the GWAS of BC risk published by the Breast Cancer Association Consortium (BCAC). In our analysis of overall BC risk, rs561177583 (associated with G. *Coprococcus*) or any related proxy SNPs (as identified using LD-Link (35)), was unavailable in the BCAC GWAS. Characteristics of the SNPs used to proxy either the abundance or presence (vs. absence) of microbial traits are shown in Supplementary Table S1. F-statistics used to examine the first “relevance” MR assumption ranged from 12.73 to 38.46 for the respective genetic instruments, suggesting that weak instrument bias was unlikely to contribute to these analyses.

When examining the relationships between microbial traits and subtype-specific BC, the number of available microbial trait-associated SNPs varied according to molecular subtype of BC. For analyses of Luminal A-Like, Luminal B-like, HER2 positive and HER2 negative in BCAC, 11 microbial trait-associated SNPs were available (rs561177583 associated with G. *Coprococcus*, rs4494297 associated with F. *Sutterellaceae* and rs150018970 associated with G. *Ruminococcus* and any possible proxies for these SNPs were unavailable). Thirteen microbial traits were available for the MR analysis of TNBC in the combined BCAC and Consortium of Investigators of Modifiers of BRCA1/2 (CIMBA) consortia, with rs561177583 (associated with G. *Coprococcus*) and any proxies for this SNP being unavailable in the GWAS meta-analysis.

In MR analyses, the strongest evidence (in terms of precision) for a possible causal effect was for the presence of bacteria within the *Ruminococcus* genera (G. *Ruminococcus*) on overall BC risk (odds ratio (OR) per approximate doubling of the genetic liability of presence vs. absence: 1.03; 95% CI: 1.00, 1.06; P=0.04; Figure 3, Supplementary Table S3). For subtypes, there was also evidence suggesting that a higher abundance of bacteria within the *Parabacteroides* order (G. *Parabacteroides*) decreased the risk of TNBC (OR per normalized SD higher abundance: 0.84; 95% CI: 0.71, 0.90; P=0.03) and the presence of an unclassified group of bacteria within the *Bacteroidales* order (G. unclassified, O. *Bacteroidales*) increased the risk of TNBC (OR per approximate doubling of the genetic liability of presence: 1.12; 95% CI: 1.03, 1.22; P=0.01) (Figure 4, Supplementary Table S3). Whilst some estimates of the associations between other microbial traits and BC risk were larger in magnitude than those presented here (e.g., G. *Veillonella* decreasing and G. *Butyricicoccus* increasing overall BC risk, and G. *Bifidobacterium* decreasing Luminal A and HER2 negative BC risk), the confidence intervals surrounding these larger estimates crossed the null. Therefore, these estimates were considered insufficiently precise to draw firm conclusions.

**Figure 3.**
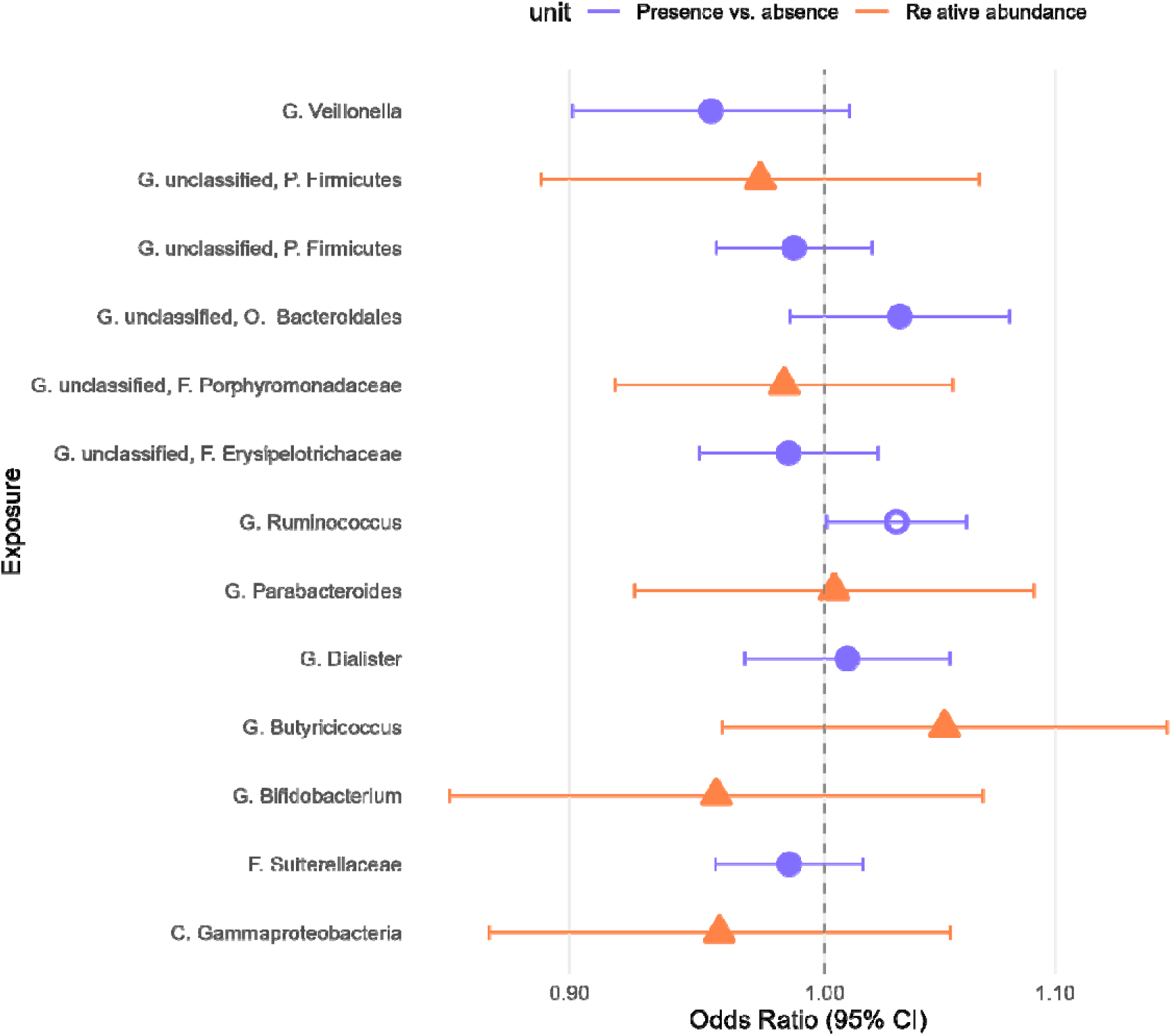
Association between microbial traits and overall breast cancer risk. Letters in microbial trait name reflect the taxonomical level: C = class; F = family; G = genus; O = order; P = phylum; unclassified = bacterial genus within the stated higher taxon could not accurately be classified. Presence vs. absence and relative abundance traits demonstrated by purple circles or orange triangles, respectively, as indicated by the legend. Open shapes represent results that met the pre-defined p-value threshold. Odds ratios between microbial traits and overall breast cancer (BC) risk, where units represent the change in risk of overall BC per normalized standard deviation (SD) in the relative abundance of each continuous microbial trait or with an approximate doubling of the genetic liability to presence (vs. absence) of each binary microbial trait.

**Figure 4.**
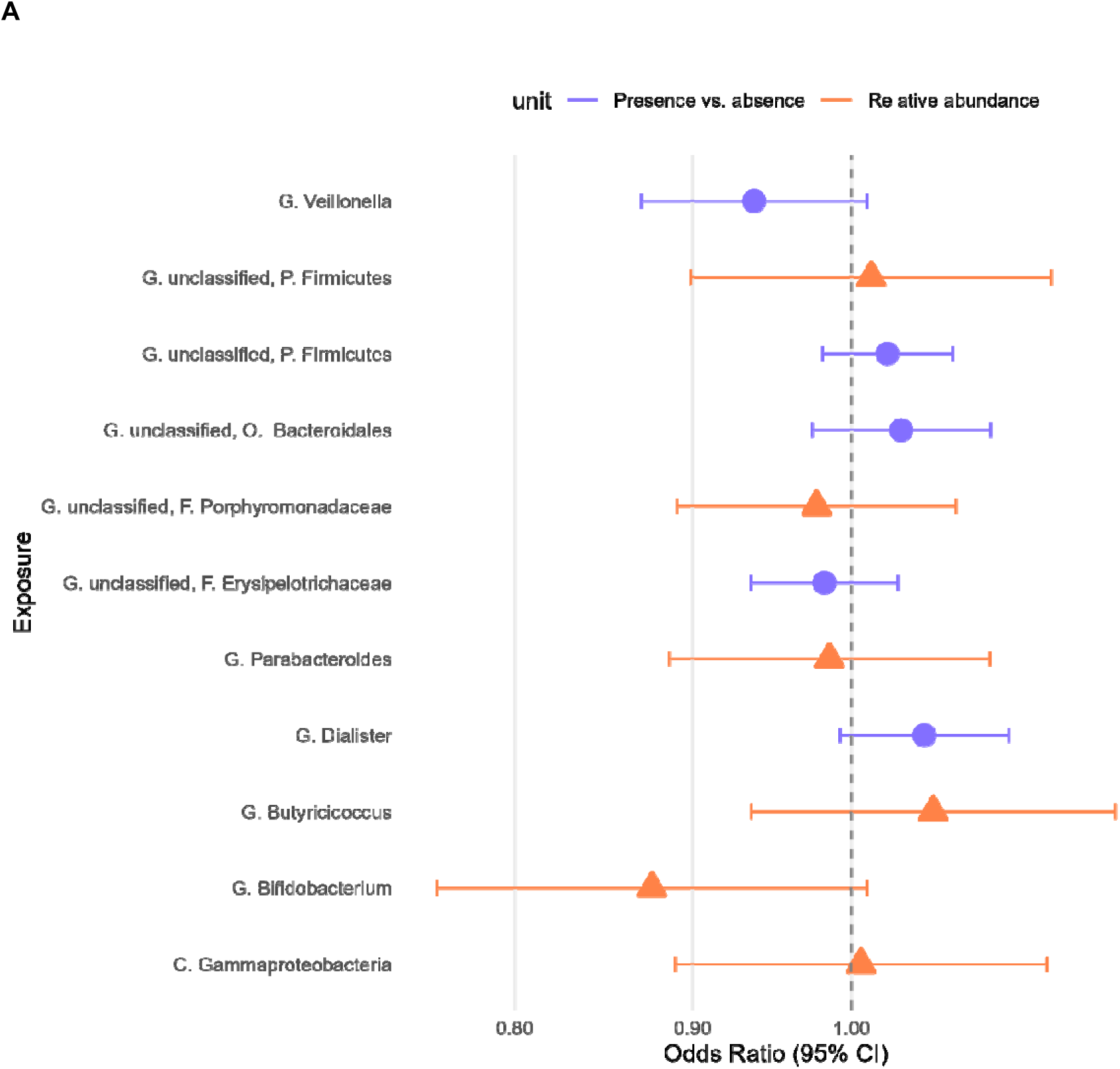
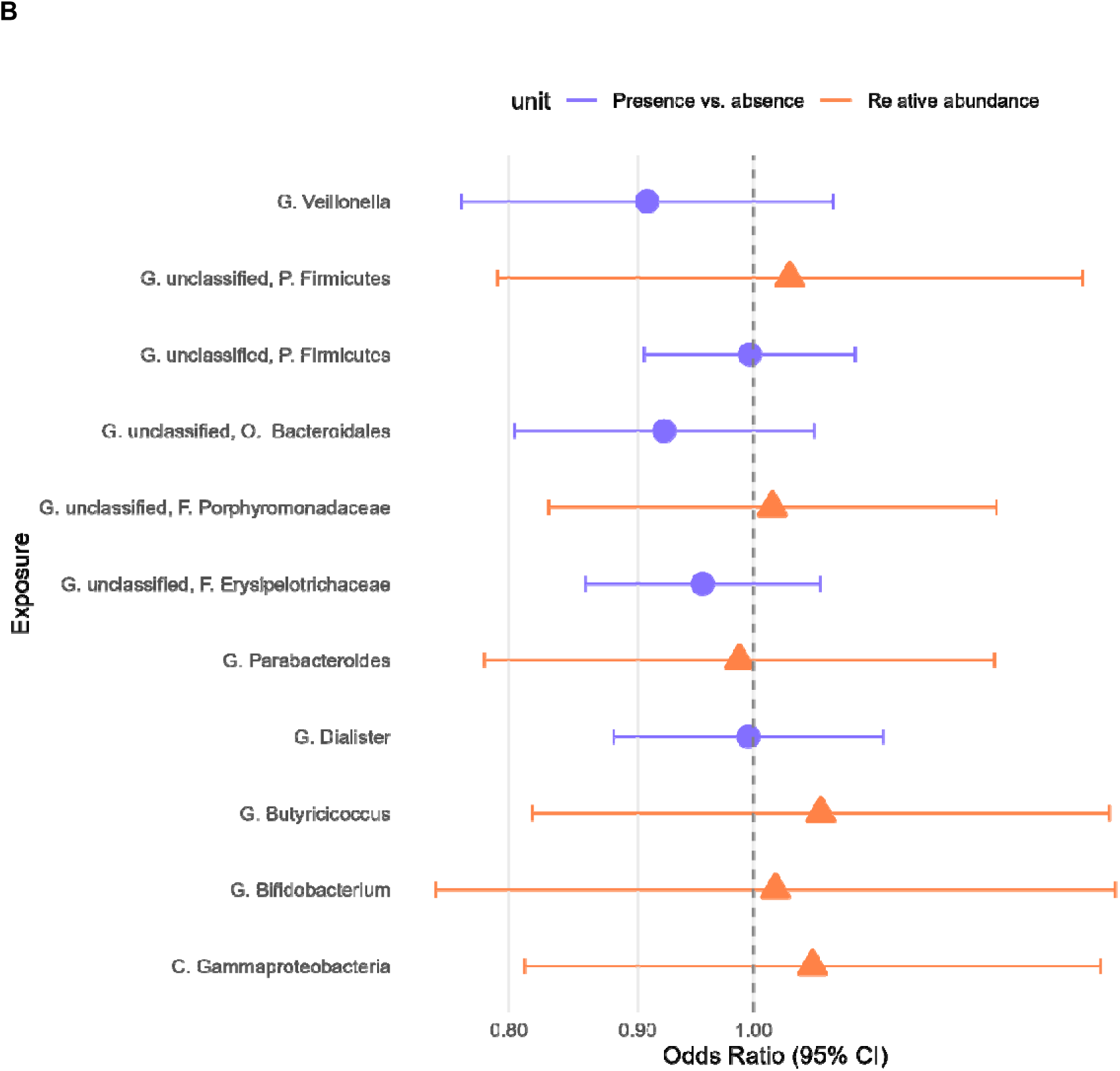
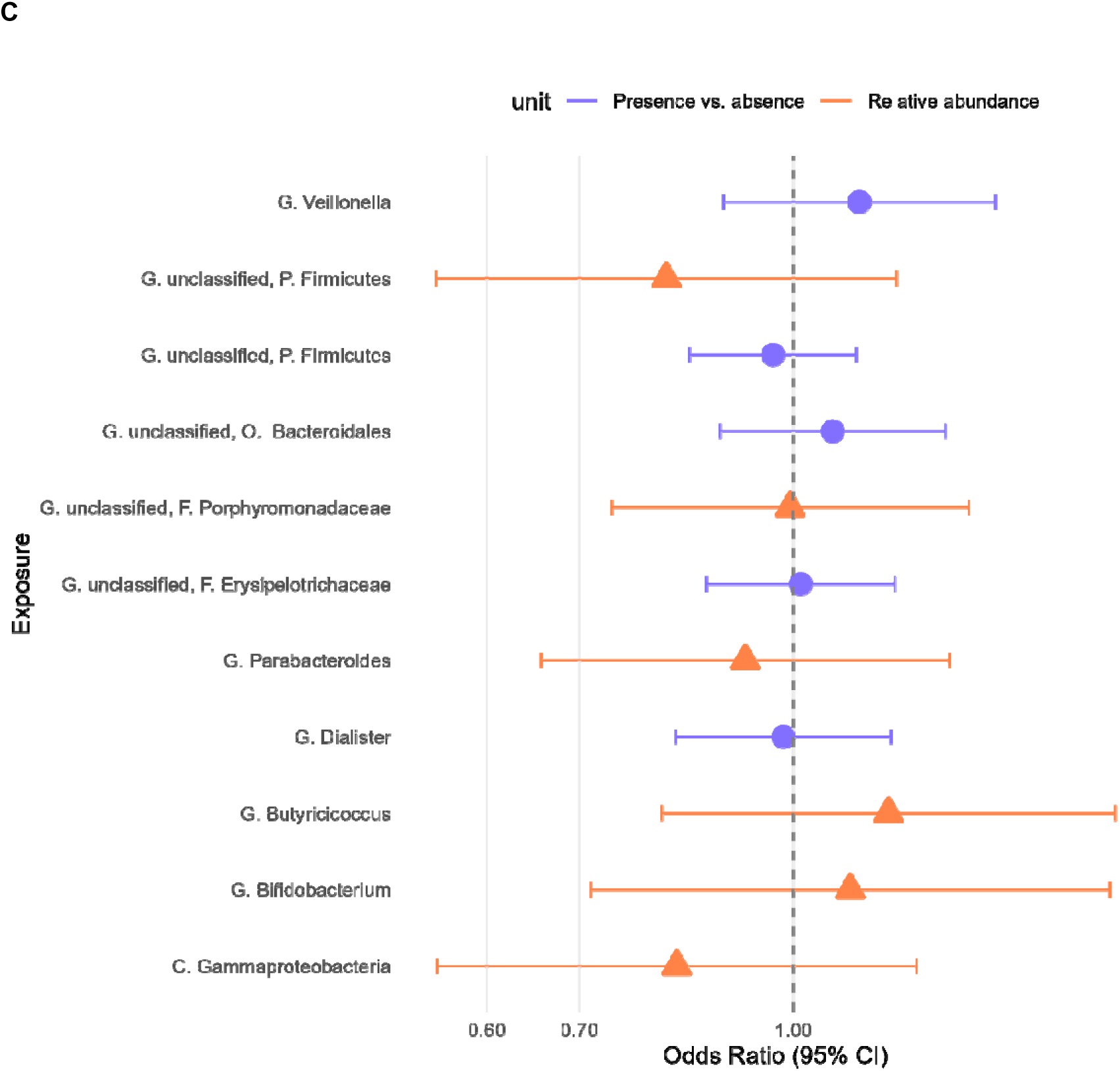
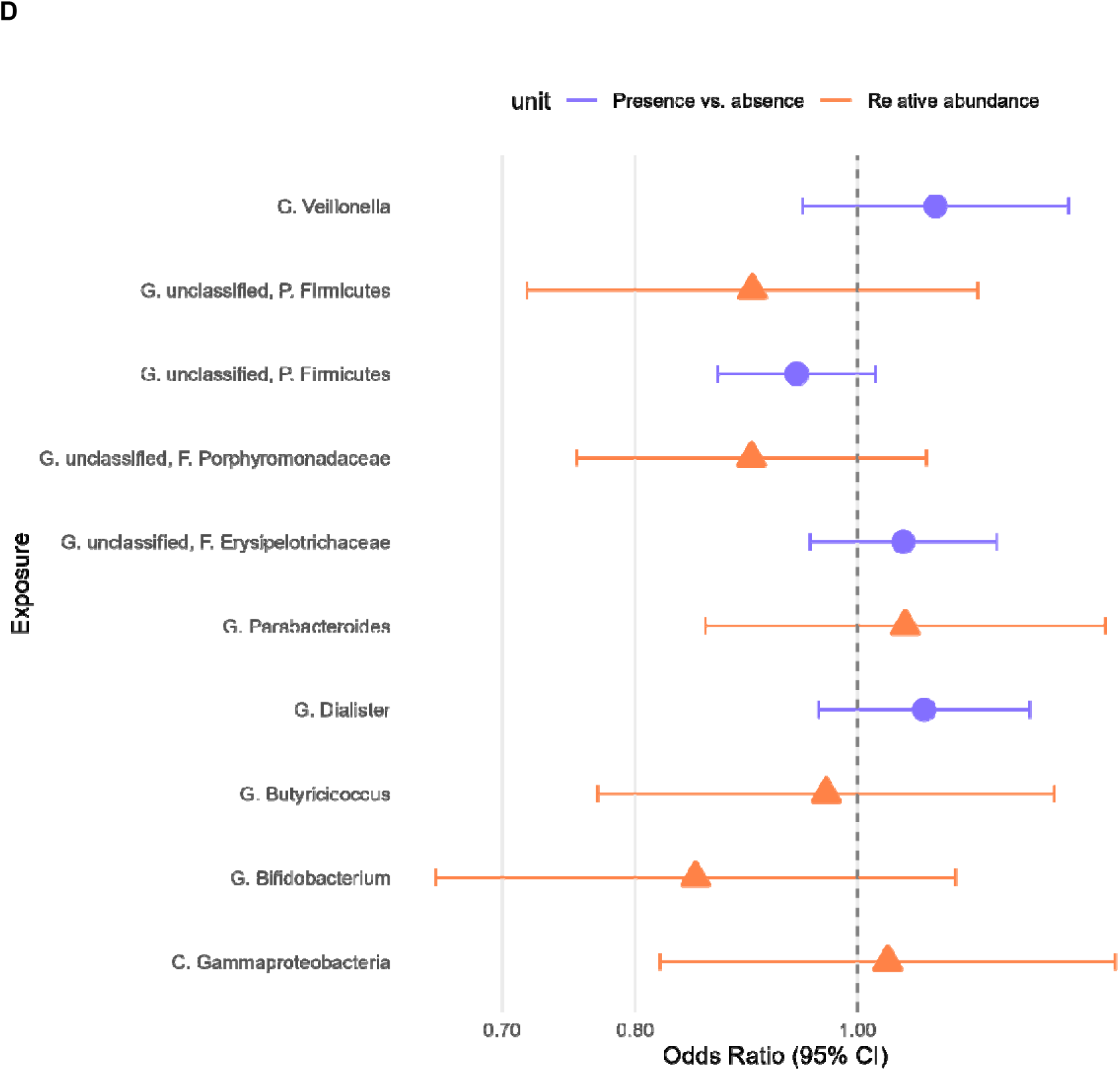
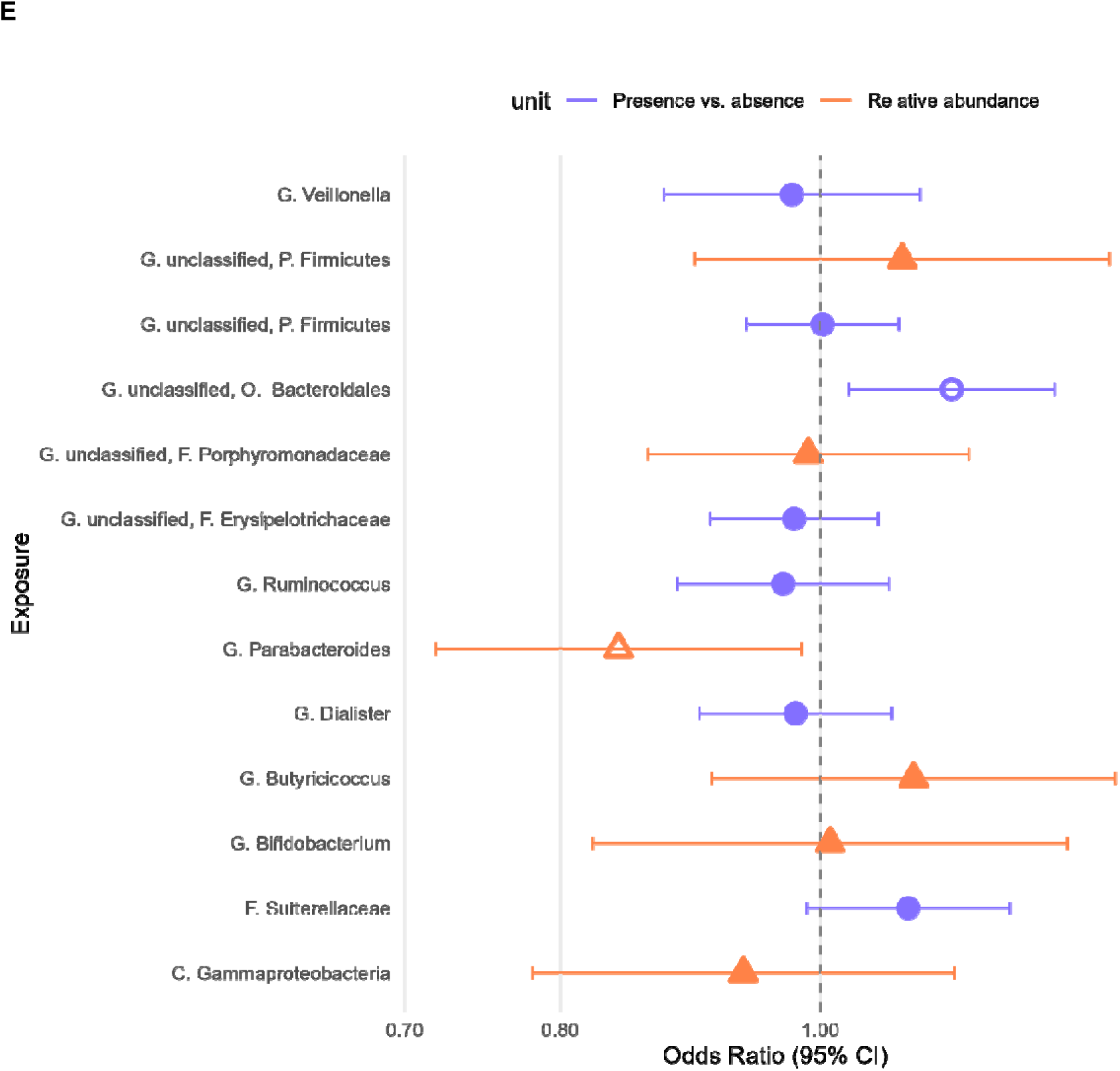
Associations between microbial traits and subtype-specific breast cancer risk. HER2 = Human Epidermal Growth Factor 2; TNBC = triple negative breast cancer. Letters in microbial trait name reflect the taxonomical level: C = class; F = family; G = genus; O = order; P = phylum; unclassified = bacterial genus within the stated higher taxon could not accurately be classified. Presence vs. absence and relative abundance traits demonstrated by purple circles or orange triangles, respectively, as indicated by the legend. Open shapes represent results that met the pre-defined p-value threshold. Odds ratios shown for microbial traits with A: Luminal A Breast Cancer, B: Luminal B Breast Cancer, C: HER2 positive breast cancer, D: HER2 negative breast cancer and E: Meta-analysed TNBC, where units represent the change in risk of each breast cancer (BC) subtype per normalized standard deviation (SD) in each continuous microbial trait abundance or with an approximate doubling of the genetic liability to presence (vs. absence) of each binary microbial trait.

### Circulating metabolites and breast cancer risk

Two-sample MR was then undertaken to identify associations between circulating metabolites measured via mass spectrometry (MS) and BC risk. Within the Shin *et al.* (33) GWAS, there were 299 SNP-metabolite associations, reflecting 218 unique SNPs associated with 219 circulating metabolites. Of these, up to 215 genetic variants associated with up to 216 unique circulating metabolites were available in the three GWAS of BC risk (Supplementary Table S2). Instruments for 208 metabolites were available in BCAC for the MR analysis of metabolites and overall, Luminal A-Like, Luminal B-like, HER2 positive and HER2 negative BC and instruments for 216 metabolites were available in the combined BCAC and CIMBA meta-analysis for MR analysis of metabolites and TNBC. SNPs associated with the other metabolites in the Shin *et al.* GWAS (or proxies of these) were not available in either the BCAC or combined BCAC and CIMBA datasets. F-statistics ranged from 30.24 to 3180.06 for the respective genetic instruments, suggesting that weak instrument bias was unlikely to impact the analyses (Supplementary Table S2).

In MR analyses, there was evidence that five metabolites altered the risk of overall BC at the p-value threshold corrected for multiple testing (i.e., 0.05/208 = 2.40×10^-04^). Specifically, homostachydrine (OR per log_10_ unit change: 0.37; 95% CI: 0.25, 0.55; P=7.28×10^-07^), 3-dehydrocarnitine (OR: 0.45; 95% CI: 0.30, 0.67; P=1.01×10^-04^) and the ratio between DSGEGDFXAEGGGVR and ADpSGEGDFXAEGGGVR (OR: 0.69; 95% CI: 0.60, 0.79; P=1.32×10^-07^) reduced the risk of overall BC. Conversely, ADpSGEGDFXAEGGGVR (OR per log_10_ unit change: 1.59; 95% CI: 1.33, 1.89; P=2.01×10^-07^) and the ratio between ADpSGEGDFXAEGGGVR and X-14304-leucylalanine (OR: 1.32; 95% CI:1.17, 1.48; P=3.07×10^-06^) increased the risk of overall BC (Figure 5, Supplementary Table S4). There was also evidence that four metabolites, several of which were the same metabolites that altered overall BC, altered subtype-specific BC risk at a p-value threshold corrected for multiple testing (i.e., 0.05/215 = 2.31×10^-04^ for TNBC and 0.05/208 = 2.40×10^-04^ for other subtypes). Specifically, homostachydrine (OR: 0.33; 95% CI: 0.20, 0.56; P=2.94×10^-05^) reduced the risk of Luminal A BC and the ratio between ADpSGEGDFXAEGGGVR and X-14304--leucylalanine (OR: 1.40; 95% CI: 1.20, 1.63; P=1.25×10^-05^) increased the risk of Luminal A BC. Tyrosine also increased the risk of Luminal A BC (OR: 15.50; 95% CI: 3.78, 63.61; P=1.42×10^-04^). There was also evidence that betaine reduced the risk of TNBC (OR: 0.16; 95% CI: 0.07, 0.35; P=3.52×10^-06^) (Figure 5, Supplementary Table S4). No other findings met the corrected p-value threshold in subtype analysis.

**Figure 5.**
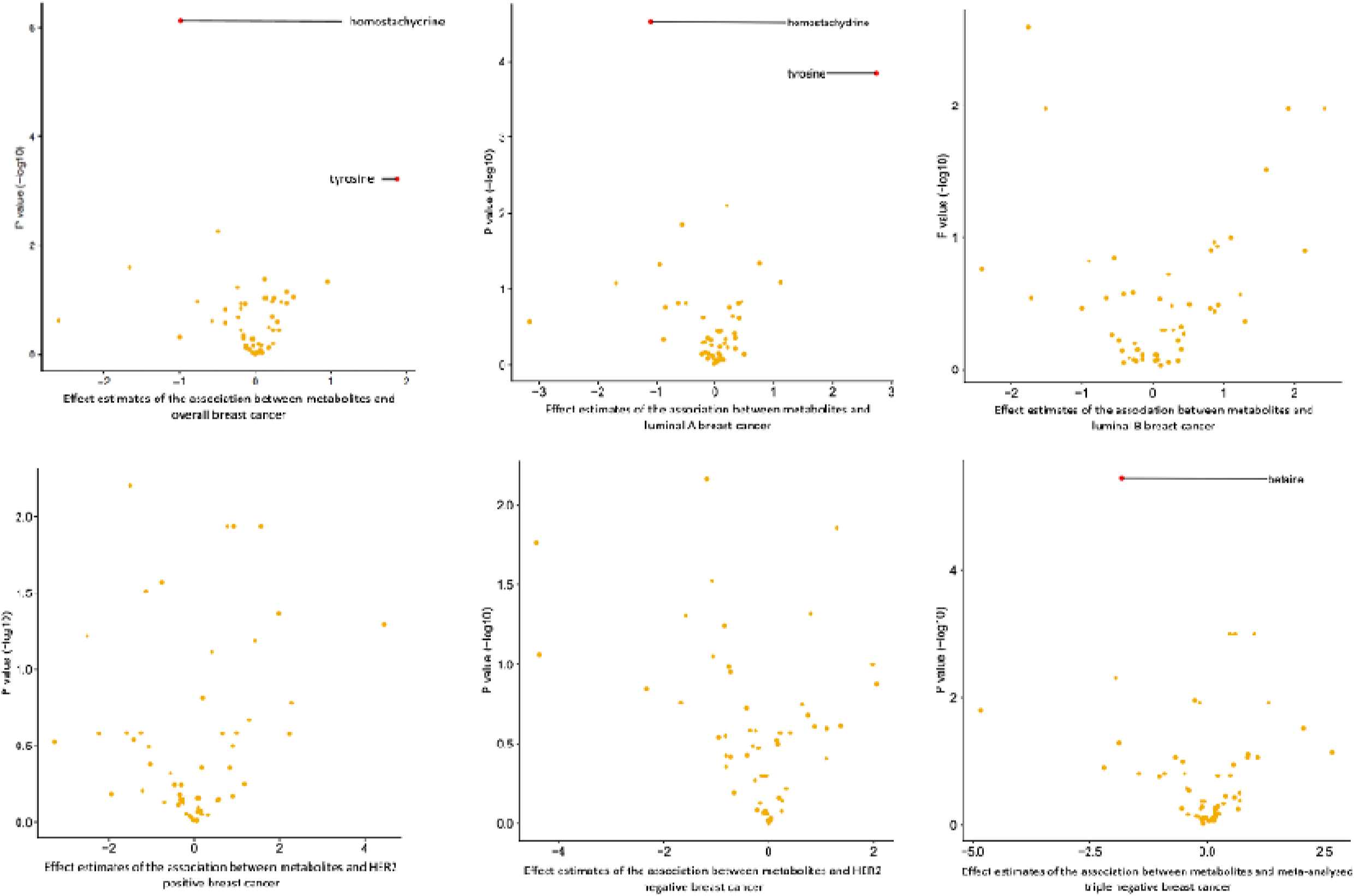
Volcano plots of Mendelian randomization analysis of circulating metabolites on breast cancer risk. HER2 = Human Epidermal Growth Factor 2. Volcano plot of results from the Mendelian randomization analysis of circulating metabolites and subtype-specific breast cancer (BC) risk. Estimates reflect the log odds of the risk of each cancer subtype (indicated in the x-axis legend) per log10 unit change in each circulating metabolite. The red dots reflect results which met the Bonferroni-corrected P value threshold (P<8.06×10^-04^ for triple-negative BC and P<8.47×10^-04^ for overall BC and all other BC subtypes) and metabolites are labelled if they pass the corrected P-value threshold.

### Identifying gut microbiome-associated circulating metabolites

As circulating metabolites may be a potential mechanism by which the gut microbiome could influence BC risk, we then investigated the relationship between the three microbial traits for which there was the strongest evidence for a causal effect on either overall BC (G. *Ruminococcus*) or TNBC (G. *Parabacteroides and* G. unclassified, O. *Bacteroidales*) and circulating metabolites. This was undertaken in two ways: firstly, using multivariable linear regression within FGFP (where we had measurements of both microbial composition and circulating metabolites from the same individuals) and secondly, using two-sample MR.

Multivariable linear regression analyses identified multiple circulating metabolites that were associated with these microbial traits within FGFP (Figure 6, Supplementary Table S5). Specifically, in multivariable linear regression analyses, at a Bonferroni-corrected p-value threshold of 3.08×10^-05^ (i.e., 0.05/(3*542), where 542 was the estimated number of independent metabolites in FGFP, see Methods) 42 metabolites were associated with G. *Ruminococcus.* The strongest of these associations (in terms of precision and magnitude) included 3-phenylpropionate (hydrocinnamate) (rank normalized SD change with presence vs. absence of *G. Ruminococcus* (beta): 1.15; 95% CI: 0.94, 1.36; P=1.02×10^-25^), isoursodeoxycholate (beta: -1.06; 95% CI: -1.27, -0.86; P=8.93×10^-24^) and X-24410 (beta: -1.00; 95% CI: -1.20, -0.80; P=2.38×10^-22^) (Supplementary Table S5). Four metabolites were associated with G. *Parabacteroides* at the same Bonferroni-corrected p-value threshold and with the largest effect sizes (rank normalized SD change per rank normalized SD change in *G. Parabacteroides* (beta) for in p-cresol sulfate: 0.13; 95% CI: 0.08, 0.18; P=2.00×10^-07^; beta for glycodeoxycholate sulfate: 0.12; 95% CI: 0.07, 0.17; P=2.75×10^-06^; beta for X-12851: 0.13; 95% CI: 0.07, 0.18; P=5.25×10^-06^ and beta for X-21286: 0.11; 95% CI: 0.06, 0.15; P=1.93×10^-05^) (Supplementary Table S5). Eleven metabolites were associated with G. *unclassified,* O. *Bacteroidales* at the Bonferroni-corrected p-value threshold (Supplementary Table S5). Of these metabolites X-11850 (rank normalized SD change with presence vs. absence of G. *unclassified,* O. *Bacteroidales* (beta): 0.51; 95% CI: 0.39, 0.64; P=2.17×10^-16^), X-11843 (beta: 0.45; 95% CI: 0.32, 0.58; P=4.93×10^-12^ ), cinnamoylglycine (beta: 0.41; 95% CI: 0.29, 0.52; P=1.74×10^-11^) and 3-phenylpropionate (hydrocinnamate) (beta: 0.42; 95% CI: 0.29, 0.54; P=4.18×10^-11^) were associated with the greatest degree of precision and were also featured amongst those metabolites with the largest magnitude of effect estimate.

**Figure 6.**
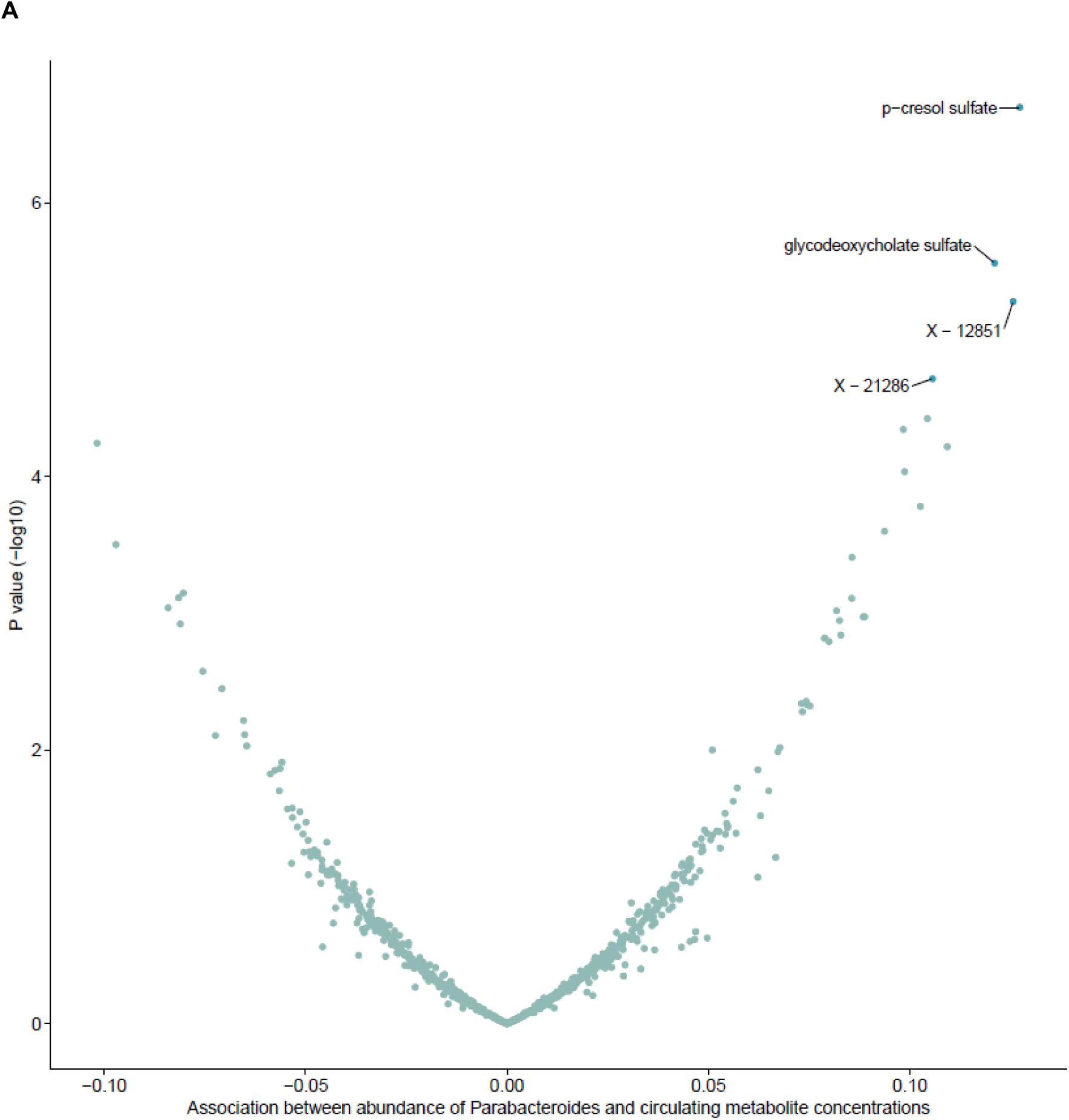
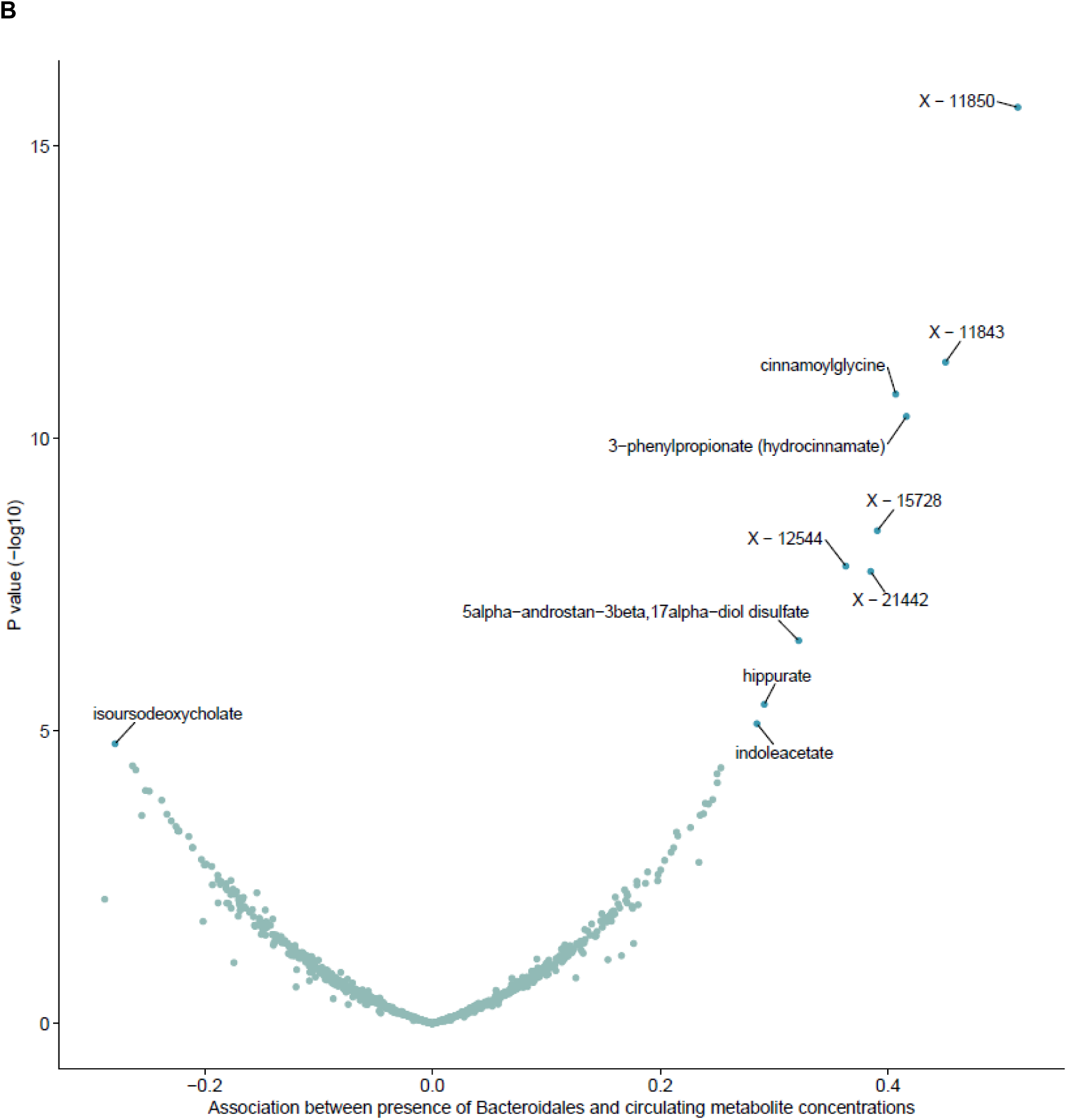
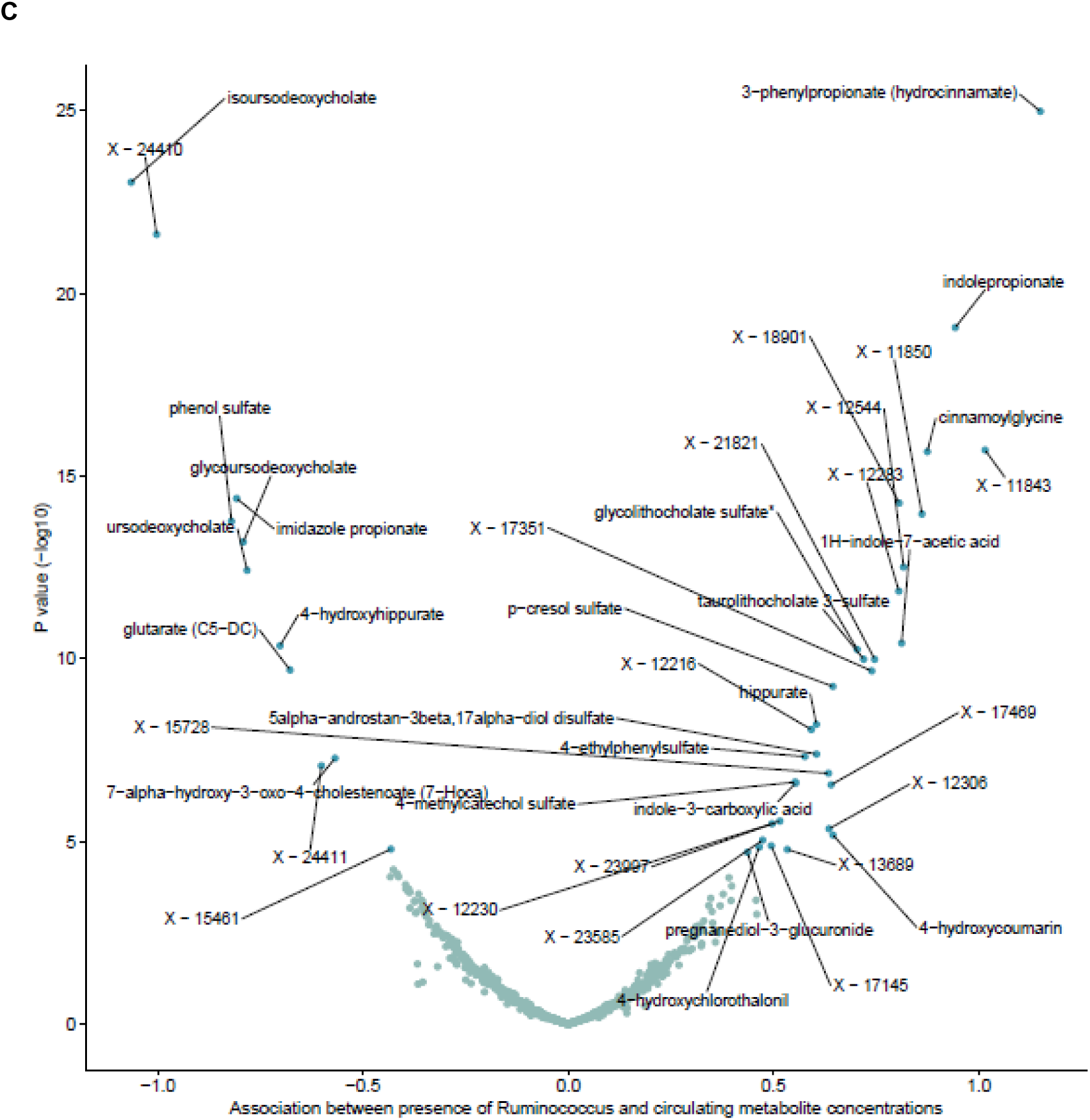
Volcano plots of linear regression analysis investigating associations between gut microbial traits and circulating metabolites within the Flemish Gut Flora Project for (A) *G. Parabacteroides*, (B) *G. unclassified, O. Bacteroidales* and (C) *G. Ruminococcus*. Letters in microbial trait name reflect the taxonomical level: G = genus; O = order; unclassified = bacterial genus within the stated higher taxon could not accurately be classified. Volcano plot of results from the linear regression analysis of gut microbial traits and circulating metabolite concentrations. The dots reflect the unit change in each metabolite with each normalized standard deviation (SD) change in each continuous microbial trait abundance or with presence (vs. absence) of each binary microbial trait (indicated by the x-axis label). The red dots reflect results which met the Bonferroni-corrected P value threshold (P<3.08×10^-05^) and metabolites are labelled if they pass the corrected p-value threshold.

Two-sample MR analyses were then undertaken using genetic variants associated with G. *Ruminococcus*, G. *Parabacteroides,* and G. unclassified, O. *Bacteroidales* from the Hughes *et al.* GWAS (27) as instruments for the three microbial traits and summary-level data from the Shin *et al.* GWAS of circulating metabolites (33). We were not able to extract SNPs (or their proxies) associated with either *G. Parabacteroides* or *G. unclassified, O. Bacteroidales* from the metabolite GWAS data; therefore, MR analyses were only conducted with G. *Ruminococcus*, where the single SNP associated with G. *Ruminococcus* was available for 17 metabolites from the Shin *et al.* GWAS summary-level data. There was little evidence that *G. Ruminococcus* was associated with alterations in the concentrations of any characterised metabolites, with no associations meeting the p-value threshold corrected for multiple testing (i.e., 0.05/17; P=2.94×10^-03^) (Supplementary Table S6). However, of the six metabolites that were analysed in both in the multivariable linear regression and two-sample MR, all effect estimates were directionally consistent (i.e., *G. Ruminococcus* was positively associated with ergothioneine and X-12729 and negatively associated with taurochenodeoxycholate, taurocholate, X-11478 and X-12847) across both analyses. Given the limited ability to run two-sample MR analyses of many metabolites for which multivariable linear regression results indicated an association with microbial traits, coupled with the directional consistency between multivariable regression and two-sample MR analyses for those that could be compared, we based our follow-up analyses (i.e., investigating the overlap in results between the gut microbiome, metabolites and BC risk) on the multivariable linear regression results alone. Therefore, overlap analyses included all three gut microbial traits (i.e., *G. Ruminococcus*, *G. Parabacteroides*, and *G. unclassified, O. Bacteroidales*) and all associated metabolites analysed across multivariable linear regression and two-sample MR (i.e., 79 for *G. Ruminococcus*, 83 for *G. Parabacteroides* and 84 for *G. unclassified, O. Bacteroidales*).

### Identifying overlap between the gut microbiome, metabolites and breast cancer risk

Integrating our analyses, we qualitatively compared the consistency of direction of association between (i) MR-derived estimates of the effect of gut microbial traits on BC risk, (ii) MR-derived estimates of the effect of circulating metabolite concentrations on BC risk and (iii) the multivariable linear regression estimates linking gut microbial traits with circulating metabolite concentrations. There was consistency in the directions of association between *G. Ruminococcus*, 23 circulating metabolites, and overall BC risk; between *G. Parabacteroides*, 35 circulating metabolites and TNBC; and between *G. unclassified, O. Bacteroidales*, 38 circulating metabolites and TNBC (Supplementary Tables S7-S9).

Whilst, in some instances, the results for these combinations of phenotypes reached the p-value thresholds specific for each analysis, there were no cases where the directionally consistent results met our pre-defined p-value thresholds across all analyses. Homostachydrine was consistently negatively associated with both *G. Ruminococcus* and overall BC risk (Supplementary Table S7); however, the strength of the evidence in multivariable linear regression linking homostachydrine and *G. Ruminococcus* was relatively weak (SD change per normalized SD higher abundance: -0.005; 95% CI: -0.20, 0.20; P=0.96). Similarly, 3-phenylpropionate (hydrocinnamate) was consistently associated with both *G. unclassified, O. Bacteroidales* and TNBC; however, the evidence from two-sample MR analyses of these metabolites on TNBC risk was relatively weak and did not meet our specified p-value thresholds for this analysis (OR per log_10_ unit change in 3-phenylpropionate (hydrocinnamate): 1.19; 95% CI: 0.49, 2.87; P=0.70). These comparisons, however, did provide some evidence for biologically plausible directionally consistent relationships between *G. Ruminococcus*, homostachydrine and overall BC, alongside *G. unclassified, O. Bacteroidales*, 3-phenylpropionate (hydrocinnamate) and TNBC worth following up further.

### Sensitivity analyses

#### Gut microbial traits and breast cancer risk

PhenoScanner and the IEU OpenGWAS databases were used to test for the presence of horizontal pleiotropy in the MR-derived effects observed between G. *Ruminococcus*, G. *Parabacteroides* and G. *unclassified,* O. *Bacteroidales* on BC risk. Within the IEU OpenGWAS database, there was little evidence for any associations between the SNPs associated with either *G. Ruminococcus* or *G. unclassified, O. Bacteroidales* at the pre-defined p-value threshold (P<2.53×10^-06^ to correct for the 39,603 GWAS datasets within the database at the time of running analyses). However, there was evidence for associations between the SNP (or proxies) associated with *G. Parabactoides* (rs13207588) and three traits relating to admission to and discharge from hospital (Supplementary Table S10). As the purpose of hospital admission was not provided, it is unclear whether admission was related to cancer; therefore, it is difficult to determine whether this observed association could explain any mechanism by which the SNP impacts BC independent from the gut microbiome (i.e., horizontal pleiotropy).

Within PhenoScanner, there was evidence for an association between the SNP (or proxies) related to *G. Ruminococcus* (rs150018970) and 20 diseases and traits (including noninflammatory disorders of the vulva and perineum, self-reported non-Hodgkin lymphoma and traits labelled as “other conduction disorders”), four unspecified gene expression loci within the adrenal gland and specific expression of *UCK1* in the pancreas at the pre-defined p-value threshold (P<0.0001, accounting for the largest number of results that the online tool provides; N=1,000; Supplementary Tables S11-S13). There was also evidence that the SNP (or proxies) associated with *G. unclassified, O. Bacteroidales* (rs116135844) was associated with one disease and trait phenotype (i.e., disorders of the patella) and two gene expression loci within whole blood (one of which was unspecified, the other was the *UBE2J2* gene) (Supplementary Tables S14-S15). The SNP (or proxies) associated with *G. Parabacteroides* was associated with one disease and trait phenotype indicative of population density within the home area, expression of *FOXP4* in whole blood and 11 DNA methylation epigenetic markers including nine identified in whole blood, one in cord blood and one specifically in T cells (Supplementary Tables S16-S19). Whilst it is difficult to discern whether some of these phenotypes may act as a horizontal pleiotropic pathway between these various SNPs and BC, it is biologically plausible that expression of these aforementioned genes could have a downstream impact on BC (e.g., the *UBE2J2*, *FOXP4* and *UCK1* genes have all been related to cardiometabolic traits, autoimmune diseases and several forms of carcinoma).

Despite some evidence for a causal effect of three microbial traits on either overall or subtype-specific BC risk, colocalisation analyses suggested that these microbial traits and BC outcomes were unlikely to share a single causal variant (Supplementary Table S20, Supplementary Figure 1). For G. *Ruminococcus* and overall BC risk, there was a 57% posterior probability that neither trait had a causal variant within the region examined, though the posterior probability for there being a causal variant associated with *G. Ruminococcus* (28%) was larger than probability for their being a causal variant associated with TNBC (9%), with a 1% posterior probability of shared causal variants across traits. For G. *Parabacteroides* and TNBC risk, there was a 52% posterior probability that only G. *Parabacteroides* had a causal variant with the region examined (compared to a 1% posterior probability for there being a causal variant associated with TNBC), with a 4% posterior probability of a shared causal variant (compared to a 41% posterior probability for there being a different causal variant in the region). For G. *unclassified,* O. *Bacteroidales* and TNBC risk, there was a 40% posterior probability that only G. *unclassified,* O. *Bacteroidales* had a causal variant (compared to the 9% probability that there was a causal variant associated with TNBC), and a 41% posterior probability that neither trait had a causal variant within the region examined, with a 2% posterior probability of a shared causal variant).

#### Circulating metabolites and breast cancer risk

For metabolites associated with BC in MR analyses that were associated with single SNPs (i.e., ADpSGEGDFXAEGGGVR/X-14304-leucylalanine, homostachydrine and tyrosine), potential evidence of horizontal pleiotropy for the associated instruments was explored as above using PhenoScanner and the IEU OpenGWAS (Supplementary Tables S10, S21-S35). There were no associations available between traits within the IEU OpenGWAS and the SNPs associated with any metabolite (see Methods; Supplementary Table S10). Within PhenoScanner, there was evidence that the SNP (or proxies) associated with homastachydrine (rs11950562) was associated with over 2500 diseases and traits, over 6800 gene expression loci, 45 proteins, 477 metabolites and over 8100 epigenetic markers (Supplementary Tables S21-S25). The SNP (or proxies) associated with tyrosine (rs9400467) was associated with 18 diseases and traits, 41 gene expression loci, 42 metabolites and 33 epigenetic markers (Supplementary Tables S26-S30). The SNP (or proxies) associated with ADpSGEGDFXAEGGGVR/X-14304--leucylalanine (rs579459) was associated with over 1000 diseases and traits, over 800 gene expression loci, over 1100 proteins, 405 metabolites and over 1000 epigenetic markers (Supplementary Tables S31-S35). Considering the number of traits identified, and the relevance of these traits to cancer aetiology and cancer-related traits (e.g., body composition, circulating blood components and glycaemic traits), the assumption of “no horizontal pleiotropy” in MR analyses may plausibly have been violated when using these single SNP instruments to proxy the effect of the desired metabolites on BC risk.

Colocalisation analyses of the metabolites with single SNP instruments provided strong evidence for a shared causal variant with each of the BC outcomes with ADpSGEGDFXAEGGGVR/X-14304-leucylalanine (Supplementary Table S36). Specifically, the posterior probabilities of colocalisation (i.e., H_4_) were 0.99 for ADpSGEGDFXAEGGGVR/X-14304-leucylalanine and overall BC risk and 0.98 for ADpSGEGDFXAEGGGVR/X-14304-leucylalanine and Luminal A BC risk. The posterior probability reflecting evidence for colocalisation was 0.82 for tyrosine and Luminal A BC risk; however, there was some LD within the genomic region around the lead SNP associated with tyrosine, which may indicate an impact of genetic confounding on the MR result. For homostachydrine, there was evidence that the homostachydrine-related SNP (rs11950562) was more strongly associated with the outcomes – both overall BC (H_2_ = 0.30 vs. H_1_ = 0.002) and Luminal A BC (H_2_ = 0.28 vs. H_1_ = 0.04) – and that there was more likely a different causal variant in the genomic region associated with both overall BC (H_3_ = 0.59) and Luminal A BC (H_3_ = 0.56) than colocalisation of the same causal variant. It is worth noting, however, that the homostachydrine-related SNP (rs11950562) reported by the Shin *et al*. GWAS (33), and therefore used as the instrument in MR analyses, did not seem to be the SNP most strongly related to homostachydrine in the GWAS data downloadable from the IEU OpenGWAS used in colocalisation analyses.

Heterogeneity across individual SNP effects for those metabolites with multiple associated SNPs for which there was evidence of a causal effect on BC risk (i.e., betaine with TNBC and 3-dehydrocarnitine, ADpSGEGDFXAEGGGVR and the ratio between DSGEGDFXAEGGGVR and ADpSGEGDFXAEGGGVR with overall BC risk) was assessed using Cochran’s Q statistic. Whilst there was little evidence for heterogeneity in the IVW analyses (e.g., P=0.87 for betaine and TNBC; P=0.64 for 3-dehydrocarnitine and overall BC; P=0.39 for ADpSGEGDFXAEGGGVR and overall BC; and P=0.67 for the ratio between DSGEGDFXAEGGGVR and ADpSGEGDFXAEGGGVR), such a test statistic is usually limited by power and interpretability with so few SNPs, as is the case here (Supplementary Table S37). Importantly, for those metabolites with three associated SNPs (i.e., betaine), there was directional consistency in the effect estimates obtained from the main IVW analysis compared with the weighted median, weighted mode and MR-Egger sensitivity analyses (Supplementary Table S4), all of which indicated a consistent negative association between betaine and TNBC. Leave-one-out analysis for betaine and TNBC did not provide evidence that the effect estimate based on the multi-allelic instrument was being driven by any single betaine-related SNP (Supplementary Figure S3). However, these analyses are possibly underpowered given the small number of SNPs (n=3) and may, in fact, be biased if the SNPs share the same horizontal pleiotropic pathways.

## DISCUSSION

In this study, we sought to utilise recently available large-scale epidemiological and genetic datasets to investigate the causal relationship between the human gut microbiome, circulating metabolites and the risk of developing BC. Our initial MR analyses suggested that bacterial members of the *Ruminococcus* genus*, Parabacteroides* genus and *Bacteroidales* order may influence either overall or subtype-specific BC. We also found evidence that several circulating metabolites that were previously implicated in the literature including amino-acids or their precursors or derivatives (specifically, homostachydrine, 3-dehydrocarnitine, the ratio between DSGEGDFXAEGGGVR and ADpSGEGDFXAEGGGVR, ADpSGEGDFXAEGGGVR, the ratio between ADpSGEGDFXAEGGGVR and X-14304-leucylalanine, tyrosine and betaine) may also influence overall and subtype-specific BC risk.

Using multivariable linear regression within FGFP, we sought to identify metabolites associated with microbial traits, which may act as possible mediators between the gut microbiome and BC aetiology. We found evidence for an association between the three microbial traits and circulating metabolites including some which have been shown to be produced by various bacterial species in multiple studies, including multi-omic studies and gut microbiome simulation (e.g., p-cresol sulfate (44)). To corroborate the findings from conventional observational epidemiological analyses, we then undertook a separate MR analysis assessing the role of these microbial traits on all metabolites. Whilst there was a limited ability to perform these MR analyses within the current study, due to the availability of microbiome-related genetic variants in the GWAS summary statistics published by Shin *et al.*(33), all estimates were directionally consistent across the two methodologies.

In qualitatively assessing for overlap between all analyses of the gut microbiome, metabolites, and BC risk, the directions of association were consistent for multiple combinations of phenotypes. Specifically, the strongest and most consistent evidence was observed for a relationship between bacteria within the *Ruminococcus* genus, homostachydrine and overall BC risk and for bacteria within the *Bacteroidales* order, 3-phenylpropionate (hydrocinnamate) and TNBC. However, there were no analyses for which results reached the pre-defined p-value thresholds across all analyses. Nevertheless, this analysis did identify trends of association between gut microbial traits, metabolites and BC risk that could be important to follow up with further causal inference analyses in larger scale cohort studies.

Importantly, sensitivity analyses demonstrated that the relationships observed between the microbial traits, metabolites and BC risk using MR were unlikely to reflect causality. Specifically, there was some evidence that the observed possible causal effects of *Ruminococcus, Parabacteroides* and *Bacteroidales* on either overall or subtype-specific BC risk were explained by horizontal pleiotropy due to the high number of associations with other traits that could also independently influence cancer risk. Colocalisation analyses also suggested that these microbial traits were unlikely to share causal variants with BC risk. As colocalisation is necessary, but not sufficient, for causality (39,45), and given the lack of knowledge of the mechanisms by which host genetic variation influences these microbial traits, we cannot confidently conclude that these microbial traits have a causal role in BC aetiology from these analyses.

With the circulating metabolites that had a single associated SNP, colocalisation analyses provided evidence for shared causal variants across the ratio between ADpSGEGDFXAEGGGVR and X-14304-leucylalanine and tyrosine with overall and Luminal A BC risk. In colocalisation analyses with homostachydrine, despite being reflective of the same GWAS summary statistics, the SNP used in MR analyses (due to being reported as the SNP most strongly associated with homostachydrine in the original metabolite GWAS) did not seem to be the strongest associated SNP in the data downloaded from the IEU OpenGWAS. Regardless, these analyses suggested that the SNP used as an instrument in MR analyses was more strongly associated with overall and Luminal A BC, providing evidence against colocalisation and evidence against causality, and the figure suggested there was a high level of LD in the region with SNPs associated with the BC outcomes. Additionally, for all metabolites with a single associated SNP, results indicated a high likelihood that SNPs were associated with numerous phenotypes that could indicate that causal effects were driven by pleiotropy. Though, it is impossible to decipher whether this is reflective of bias-inducing horizontal or mechanistic vertical pleiotropy. For metabolites with multiple associated SNPs, there was limited heterogeneity across SNPs associated with 3-dehydrocarnitine, ADpSGEGDFXAEGGGVR, the ratio between DSGEGDFXAEGGGVR and ADpSGEGDFXAEGGGVR and betaine with BC risk, with constant effects observed across all pleiotropy-robust MR methods with betaine and TNBC. With the limited number of associated SNPs, we were unable to provide conclusive evidence with MR analyses that either gut microbial traits or circulating metabolites had causal effects on BC risk. However, further work is required to investigate and assess the causal role of many of these metabolites, for which there was evidence for colocalisation and consistent effects over several MR methods, with overall and subtype-specific BC risk.

Recently, two MR studies provided evidence for a causal relationship between several gut microbiota and the risk of several cancers (including BC). Comparing the estimates for those microbial traits common across our paper and those published previously, Long *et al*. reported evidence that bacteria within the *Ruminococcaceae* family decreased BC risk (46), while in our study we found an increased risk of BC. Wei *et al.* provided evidence that bacteria within the *Parabacteroides* genus decreased BC risk (47); however, this was inconsistent with our findings, where there was very little evidence for an effect of *Parabacteroides* bacteria on overall BC risk but evidence for a reducing effect on TNBC risk. However, it is not clear if the results for the other microbial traits we analysed are consistent with those previously published, as authors do not present the results for all microbial traits.

There are several key explanations that are likely to explain these differences in findings across papers. Firstly, authors used a different dataset (i.e., the MiBioGen consortium (48)) as the source of their microbiome-related genetic variants. Whilst the sample size of the MiBioGen consortium is larger than the GWAS utilised in this current analysis (n=18,340), there are samples of non-European individuals within the MiBioGen GWAS, which could bias estimates if the effects of genetic variants on the gut microbial traits differ across those population samples. Unlike the Hughes *et al*. GWAS, the way in which the microbial traits were transformed and analysed differ across these individual cohorts, the heterogeneity of which may bias the MR-derived effect estimates or decrease their precision.

Secondly, there are several methodological differences in the generation of microbiome-related instruments that would likely drive differences in results, even if we had used the MiBioGen consortium as our source of microbiome-related instruments (i.e., Long *et al.* excluded SNPs that were deemed to be horizontally pleiotropic before the MR analyses were conducted, rather than assessing the impact of these SNPs on the main results, both Long *et al.* and Wei *et al.* removed palindromic SNPs despite the effect allele frequency being available for harmonization across the exposure and outcome datasets and Wei *et al*. also removed SNPs that had an F-statistic lower than 10). Thirdly, whilst Long *et al*. use the traditional genome-wide p-value threshold to select microbiome-related instruments (which is best practice for MR analyses), they also used a lenient p-value threshold (P<1×10^-06^) and Wei *et al.* use a similarly lenient p-value (P<1×10^-05^) as their main analyses, which could induce bias, through weak instrumentation, horizontal pleiotropy, genetic confounding and reverse causation. The presence of horizontal pleiotropy in the paper published by Long *et al.* was also not clear given they do not present the results of their sensitivity analyses. Lastly, the GWASs of BC risk used in both previously published papers were much smaller than the data used in the current analysis and all results showing evidence for a causal role of microbial traits on BC in the Long *et al.* paper were those using UK Biobank. Whilst authors also conducted analyses using the BCAC GWAS data, it is unclear whether these results were consistent with those using UK Biobank (and those that we present in the current study), as the results are not presented due to not reaching their defined p-value thresholds. Importantly, whilst both previous papers used the comparable datasets of summary-level data for both the microbiome and BC risk, their results are not consistent.

Various assumptions must be satisfied in MR analyses, and we utilised a combination of statistical tests to demonstrate adequate instrument strength, and to reduce our suspicion that associations of microbial traits, metabolites and BC were affected by horizontal pleiotropy. Regarding the first core MR assumption (i.e., the “relevance” assumption), the current two-sample MR analyses utilized genetic variants that were strongly associated with each trait (and thus unlikely to be susceptible to weak instrument bias); however, most of these genetic instruments contained only a single SNP, meaning that our results lacked precision. This also meant that we were largely unable to undertake sensitivity analyses to examine for evidence of horizonal pleiotropy (i.e., the third MR assumption), such as MR-Egger (41), weighted median (42), weighted mode (43) and leave-one-out analyses. Even where these methods were possible to undertake, most analyses included few SNPs meaning that these analysis results were particularly difficult to interpret and, in fact, likely biased (which is indeed why these methods should be interpreted with caution when applying in scenarios with few SNPs).

Regarding the second MR assumption (i.e., the “independence” assumption), our analyses were restricted to participants of European ancestries, and the GWASs used in the current study were adjusted for principal components to remove variation likely due to differences in ancestry or population structure, reducing the likelihood that the assumption was violated. Other strengths of this analysis include use of colocalisation to interrogate the robustness and potential causal nature of any associations identified. It is worth noting that, whilst other colocalisation methods, which do not assume the existence of singular causal variants in a genomic region, could have been used here, it is likely that the conclusions would not have changed considering the power and signal strength for the gut microbiome given the available data. Furthermore, the sample size of the summary-level data for BC outcomes were the largest to date (e.g. 136,062 cases and 111,113 controls for overall BC). However, the current study has several limitations.

Whilst multivariable linear regression analyses identified associations between bacterial members of the *Ruminococcus* genus, *Parabacteroides* genus and *Bacteroidales* order with circulating metabolite concentrations within FGFP, there was limited availability of microbiome-related SNPs in the metabolite GWAS summary-level data to corroborate these findings using MR. Therefore, we took only those metabolites with which these three microbial traits were associated into follow-up analyses. The fact that we were unable to undertake MR analyses, to which we could have compared our results from linear regression analyses, generates uncertainty about the reliability of the observed relationships, and whether they could be explained by external (e.g., confounding factors). Given the difference in sample size across microbial traits and across circulating metabolomic features, there was a varying level of power (and therefore precision in derived estimates) when comparing across these traits within individual analyses. Although we considered the magnitude and precision of effect estimates, together with consistency in the direction across multiple tests, as the main basis of scientific conclusions, greater sample sizes will ultimately improve the power and ability to draw more conclusive causal inference in future analyses. The composition of the gut microbiome varies over the lifecourse in response to environmental factors and, in women, after menopause(49). Similarly, the circulating metabolome varies with age, sex, diet, lifestyle and the composition of the gut microbiome(50).

Although the genetic variants associated with gut microbiome composition and metabolites utilised in the current study were derived from a large and meta-analysed GWASs, which aim to identify genetic variants associated with microbial traits and the metabolome in a way that minimises environmental influences (e.g., population structure, which could bias MR estimates), such influences cannot be completely eliminated. In general, little is known about the mechanisms that link host genetic variation with the gut microbiome; therefore, it is likely that the signals from GWASs of microbial traits are reflective of host-driven effects upstream of the gut microbiome (which could bias MR analyses if such effects are independently associated with either circulating metabolites or BC, in this instance). This also means that it is currently difficult to confidently discern whether the high number of associations between microbiome-related SNPs and other traits that could also influence cancer risk found within our sensitivity analyses represent horizontal pleiotropy (and thus an invalidation of MR assumptions and biased estimates) or vertical pleiotropy (i.e., a mechanism by which the SNP influences cancer risk through both the microbial and other traits). Until we have a greater understanding of the host genetic contributions to the gut microbiome and the biological mechanisms by which they occur, this will remain to be a major limitation of the current applications of MR to understand the causal role of the gut microbiome in human health and disease.

The microbiome and metabolite GWASs used in the current study were also derived from sex-combined samples and combined with a sex-specific BC dataset. If, for example, the effect of genetic variation on either gut microbial traits or metabolites differed between men and women, the causal effects derived from MR analyses using samples not representative of the same underlying population of the outcome sample (in this case, women) may bias results away from or towards the null depending on the SNP effects in males. Similarly, these analyses were undertaken using population samples of European ancestry; therefore, it is not clear how generalizable these results are to other populations, especially those with differing gut microbial compositions (e.g., relating to dietary and environmental characteristics).

An additional limitation of GWASs of microbial traits is that these traits were only characterizable at the genus or higher levels of bacterial taxonomy due to the nature of 16S rRNA sequencing. Therefore, it is possible that there are conflicting relationships between different bacterial species within these clades, and BC risk. Relating to this characterisation of phenotypes, various metabolites analysed here were ratios between two existing metabolites. Whilst the use of covariate-adjusted exposures in MR analyses can induce bias in cases where the covariate likely influences the outcome as well as the originally unadjusted exposure, analyses of metabolite ratios are more likely to influence the interpretation of effect estimates rather than induce considerable bias.

Lastly, colocalisation analyses were reliant on genome-wide summary-level statistics from the GWAS of the microbiome, metabolome and BC risk. Whilst we had access to full summary-level data for the gut microbiome and BC risk, data for the former were only available in the FGFP cohort alone and not for the meta-analysis of FGFP, FoCus and PopGen (i.e., due to the GWAS pipeline conducted by Hughes *et al*. (27)). Therefore, our colocalisation analyses assessing the robustness of MR analyses assessing the role of the gut microbiome and BC risk were restricted and likely underpowered given that the strength of association between SNPs and microbial traits was weaker than that presented in the full meta-analysis. Furthermore, it is worth noting that the current colocalisation method assumes the presence of a single causal variant in the genomic region, which may not be appropriate in some cases; therefore, other colocalisation methods that do not carry this assumption should be explored.

In the future, the availability of more microbiome- and metabolite-related SNPs from larger GWASs, importantly coupled with greater knowledge of the mechanisms by which genetic variation influences these traits, may allow greater confidence and precision in identifying associations with disease outcomes. Additional further investigation of the relationships between individual bacterial species and the risk of BC, and potential underlying mediating mechanisms, will need further and larger laboratory, epidemiological and genetic studies (including larger and more specific GWASs).

In conclusion, whilst our initial analyses suggested that the gut microbiome and certain circulating metabolites may be associated with BC risk, importantly, sensitivity analyses demonstrated that these relationships are complex and, certainly for the case of the gut microbiome, unlikely to reflect causality. To improve causal inference when interrogating the role played by the gut microbiome or metabolites in BC (and other health outcomes) within an MR framework, larger GWASs delivering a greater numbers of SNPs may provide the ability to apply appropriate sensitivity analyses. However, greater knowledge of how genetic variation influences the gut microbiome coupled with appropriate implementation of sensitivity analyses assessing the robustness of findings, cautious interpretation of results (as we have demonstrated here) and a triangulation approach comparing results from differently designed studies, are required when making claims of causality. It is crucial that future studies examining the relationship between the gut microbiome (and the metabolome) and human disease cautiously and considerately interrogate their findings, as claims of causality generated using MR can be spurious or damaging if not supported by additional methodology and appropriate sensitivity analyses.

## Supporting information

STROBE-MR Reporting

## Data Availability

Summary statistics for the microbiome-associated SNPs were obtained from the paper published by Hughes et al. (27) comprising three cohort studies (i.e., FGFP, FoCus and PopGen; N=3,890). Summary-level data from this GWAS is also available online at the University of Bristol data repository, at https://data.bris.ac.uk/data/dataset/22bqn399f9i432q56gt3wfhzlc. Individual-level data from FGFP are not open access, but are available by application to the FGFP data access committee, subject to a data use agreement with the FGFP and organized via email to. Please see the study website http://www.raeslab.org/companion/fgfp-gwas/ for further details. Summary-level data from the BCAC and BCAC/CIMBA GWAS are publicly available at https://bcac.ccge.medschl.cam.ac.uk/bcacdata/ (accessed in 2020). Summary-level data from the GWAS of metabolites published by Shin et al. and are publicly available through the IEU OpenGWAS project https://gwas.mrcieu.ac.uk/.

http://www.raeslab.org/companion/fgfp-gwas/

https://bcac.ccge.medschl.cam.ac.uk/bcacdata/

https://gwas.mrcieu.ac.uk/

## ACKNOWLEDGEMENTS

GE is funded by the Elizabeth Blackwell Institute for Health (Bristol, UK) through a Wellcome Trust ISSF (grant ID 204813/Z/16/Z), and Bristol Veterinary School. TR is supported by a National Institute for Health Research (NIHR) Development and Skills Enhancement Award (NIHR302363). The funding bodies acknowledged here did not have any role in the design of the study, the collection, design analysis, and interpretation of data or in the writing of the current manuscript. KHW is supported by Cancer Research UK [grant number RCCPDF\100007] and the University of Bristol. JR is funded by KU Leuven, VIB and the Rega Institute. NJT is a Wellcome Trust Investigator (202802/Z/16/Z), is the PI of the Avon Longitudinal Study of Parents and Children (MRC & WT 217065/Z/19/Z), is supported by the University of Bristol NIHR Biomedical Research Centre (BRC-1215-2001), the MRC Integrative Epidemiology Unit (MC_UU_00011/1) and works within the CRUK Integrative Cancer Epidemiology Programme (C18281/A29019). DH is supported by the Wellcome Trust Investigator Award (202802/Z/16/Z; PI: NJT). JY is supported by a Cancer UK Population Postdoctoral Fellowship (C68933/A28534). Where authors are identified as personnel of the International Agency for Research on Cancer/World Health Organization, the authors alone are responsible for the views expressed in this article and they do not necessarily represent the decisions, policy or views of the International Agency for Research on Cancer/World Health Organization.

## AUTHOR CONTRIBUTIONS

Conceptualization GE, TR, KHW; Data curation GE, TR, DH, KHW; Formal Analysis GE, TR, DH, JY, KHW; Funding acquisition, GE, TR, KHW; Investigation GE, TR, DH, BH, JY, KHW; Methodology GE, TR, DH, JY, KHW; Project administration GE, TR, KHW; Resources JY, DH, LG, JB, JR, NT; Supervision KHW; Validation GE, TR, DH, JY, NJT, KHW; Visualization GE, TR, ML; Writing – original draft GE, TR; Writing – review & editing GE, TR, KHW. All authors approved the final draft of the manuscript.

## DATA AVAILABILITY STATEMENT

Summary statistics for the microbiome-associated SNPs were obtained from the paper published by Hughes *et al.* (27) comprising three cohort studies (i.e., FGFP, FoCus and PopGen; N=3,890). Summary-level data from this GWAS is also available online at the University of Bristol data repository, at https://data.bris.ac.uk/data/dataset/22bqn399f9i432q56gt3wfhzlc. Individual-level data from FGFP are not open access, but are available by application to the FGFP data access committee, subject to a data use agreement with the FGFP and organized via email to. Please see the study website http://www.raeslab.org/companion/fgfp-gwas/ for further details. Summary-level data from the BCAC and BCAC/CIMBA GWAS are publicly available at https://bcac.ccge.medschl.cam.ac.uk/bcacdata/ <u>(accessed in 2020)</u>. Summary-level data from the GWAS of metabolites published by Shin *et al.* and are publicly available through the IEU OpenGWAS project https://gwas.mrcieu.ac.uk/.

## ADDITIONAL INFORMATION

Tim Robinson has received grants from Merck-Serono Dohme, Daiichi-Sankyo Company Limited and Amgen Inc. to attend conferences and educational workshops. No other authors declare any conflicts of interest.

