## Supplementary Figure 1 for "Exploring the causal role of the human gut microbiome in breast cancer risk"

**Supplementary Figure 1: Regional association plots demonstrating genetic colocalisation results for microbial traits and breast cancer risk**

A)

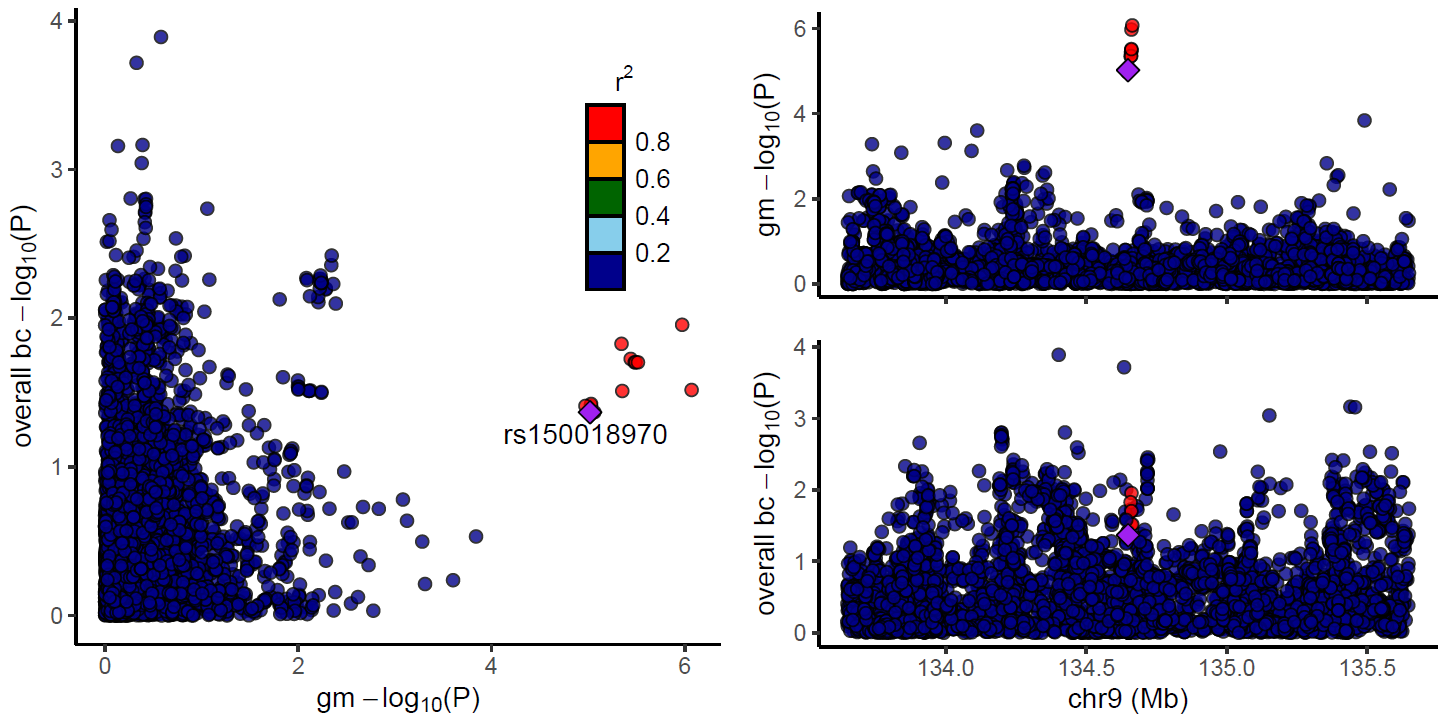

B)

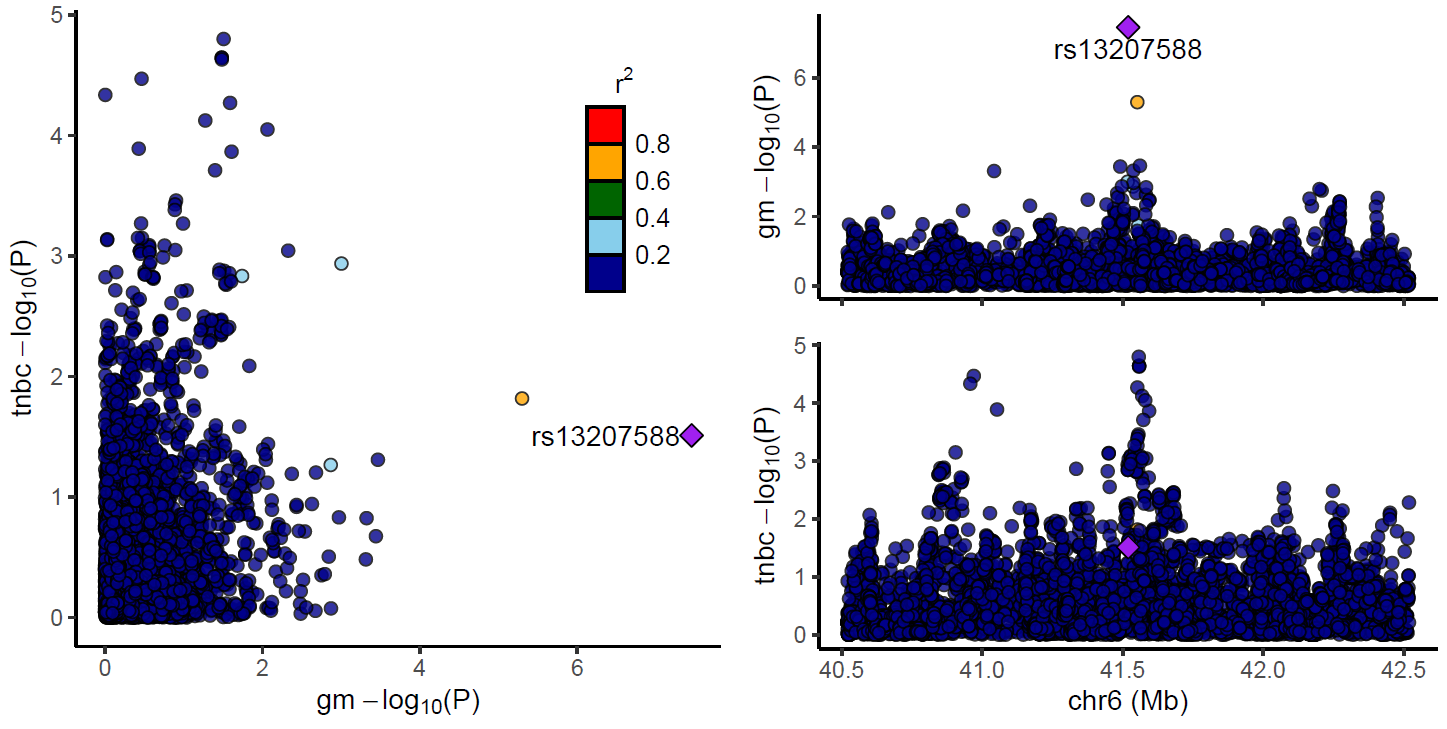

C)

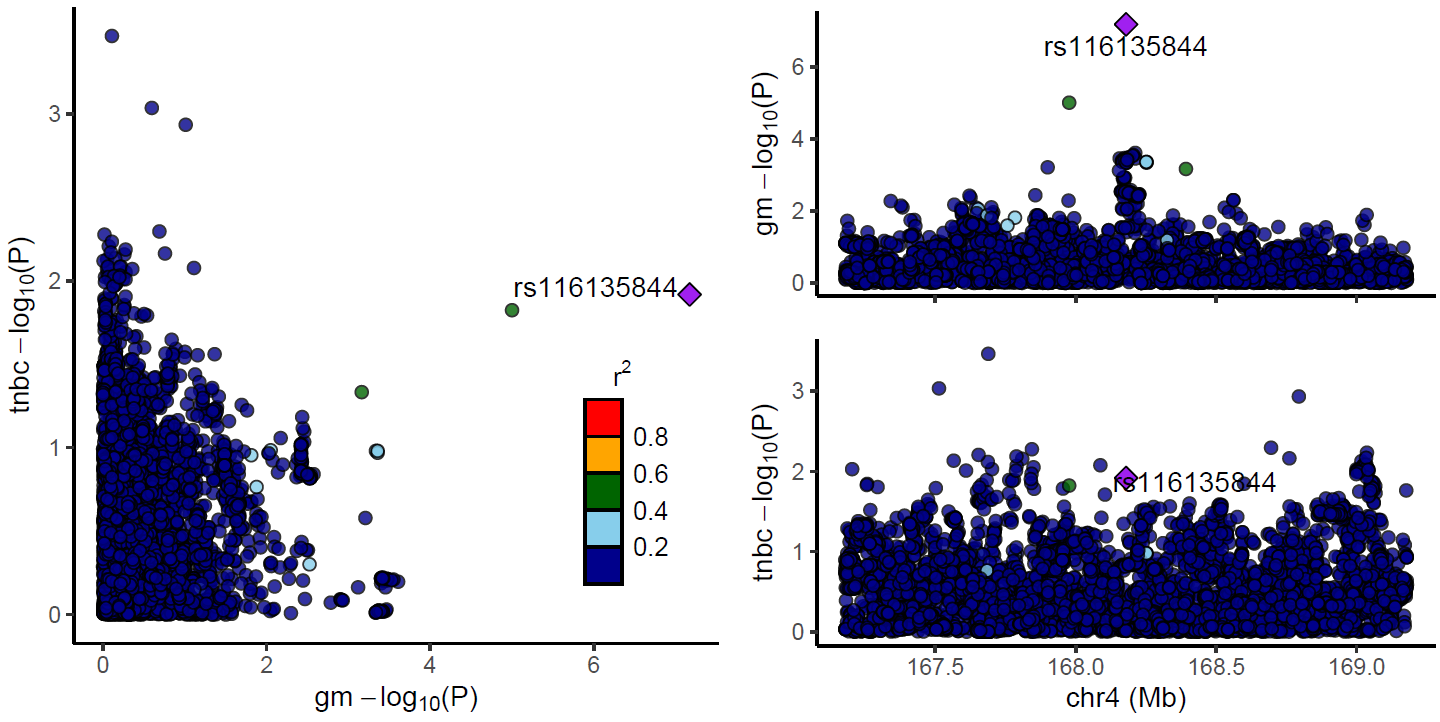

*bc = breast cancer; gm = gut microbiome; tnbc = triple negative breast cancer. Regional association plots were created using summary-level data from the Flemish Gut Flora project and Breast Cancer Association Consortium or the Consortium of Investigation of Modifiers of BRCA1/2 with the LocusCompareR package. The -log10 p-value of each lead SNP is shown by a mauve diamond. Figures show colocalisation results for the (A) lead SNP, rs150018970, associated with presence/absence of bacteria within the Ruminococcus genus and overall breast cancer risk; (B) lead SNP, rs13207588, associated with abundance of bacteria within the Parabacteroides genus and meta-analysed triple negative breast cancer risk; and (C) lead SNP, rs116135844, associated with presence/absence of unclassified bacteria within the Bacteroidales order and meta-analysed triple negative breast cancer risk.*
