## Supplementary Figure 2 for "Exploring the causal role of the human gut microbiome in breast cancer risk"

**Supplementary Figure 2: Regional association plots demonstrating genetic colocalisation results for metabolites and breast cancer risk**

A)

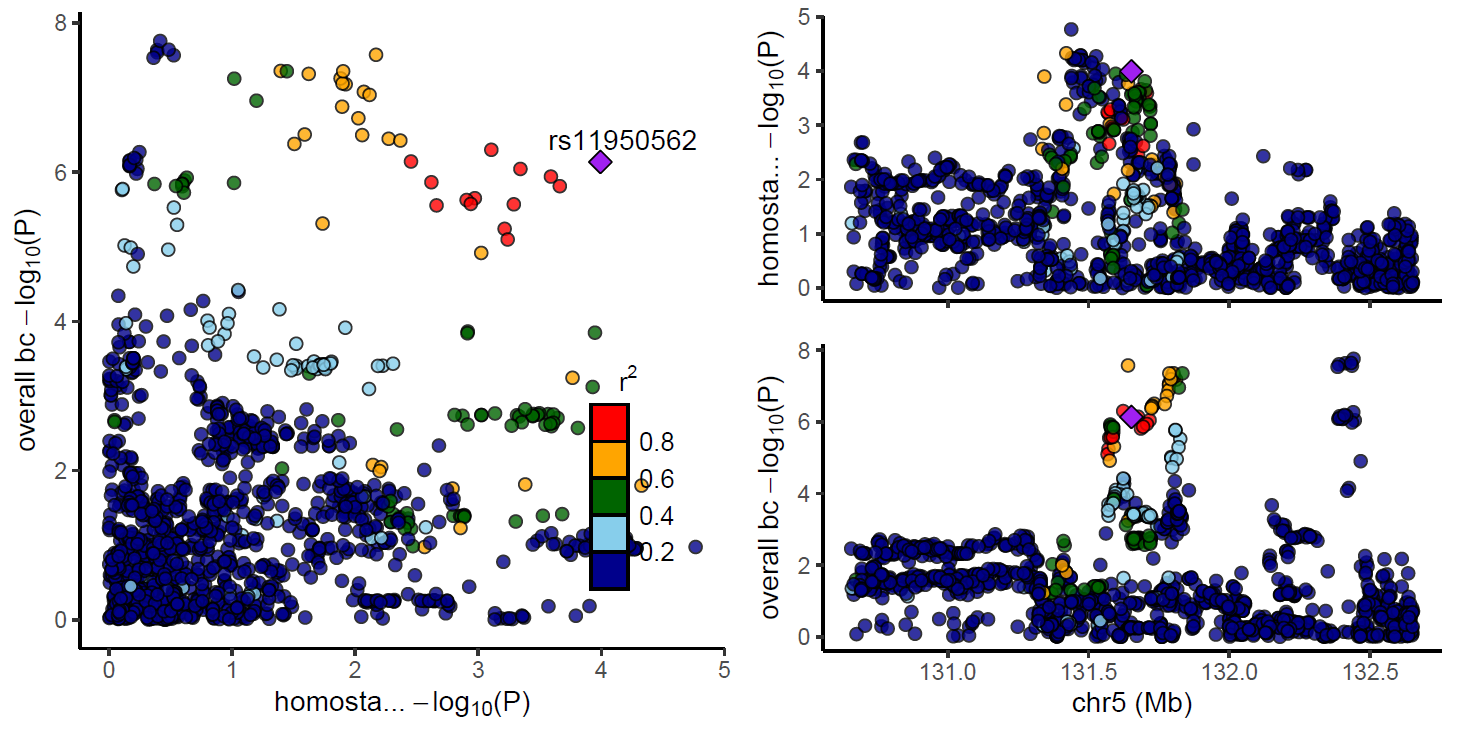

B)

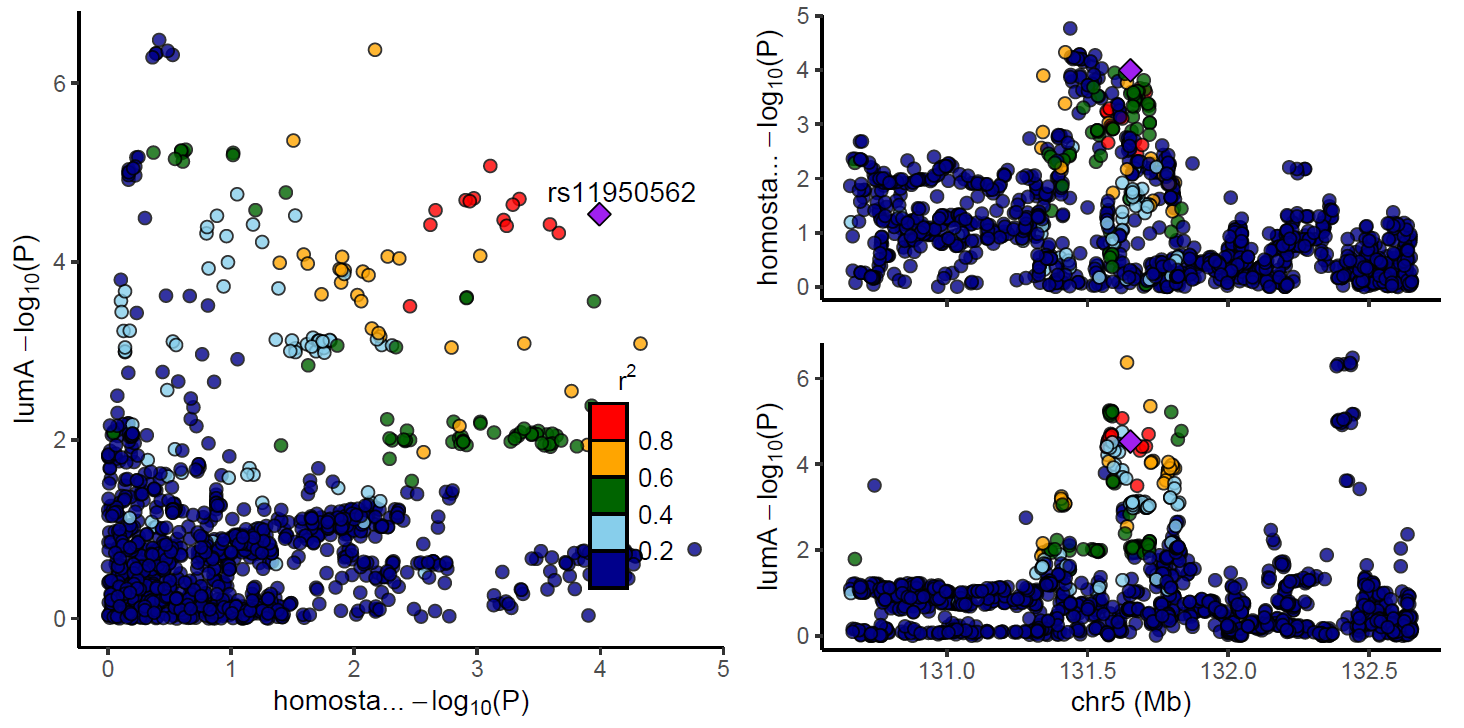

C)

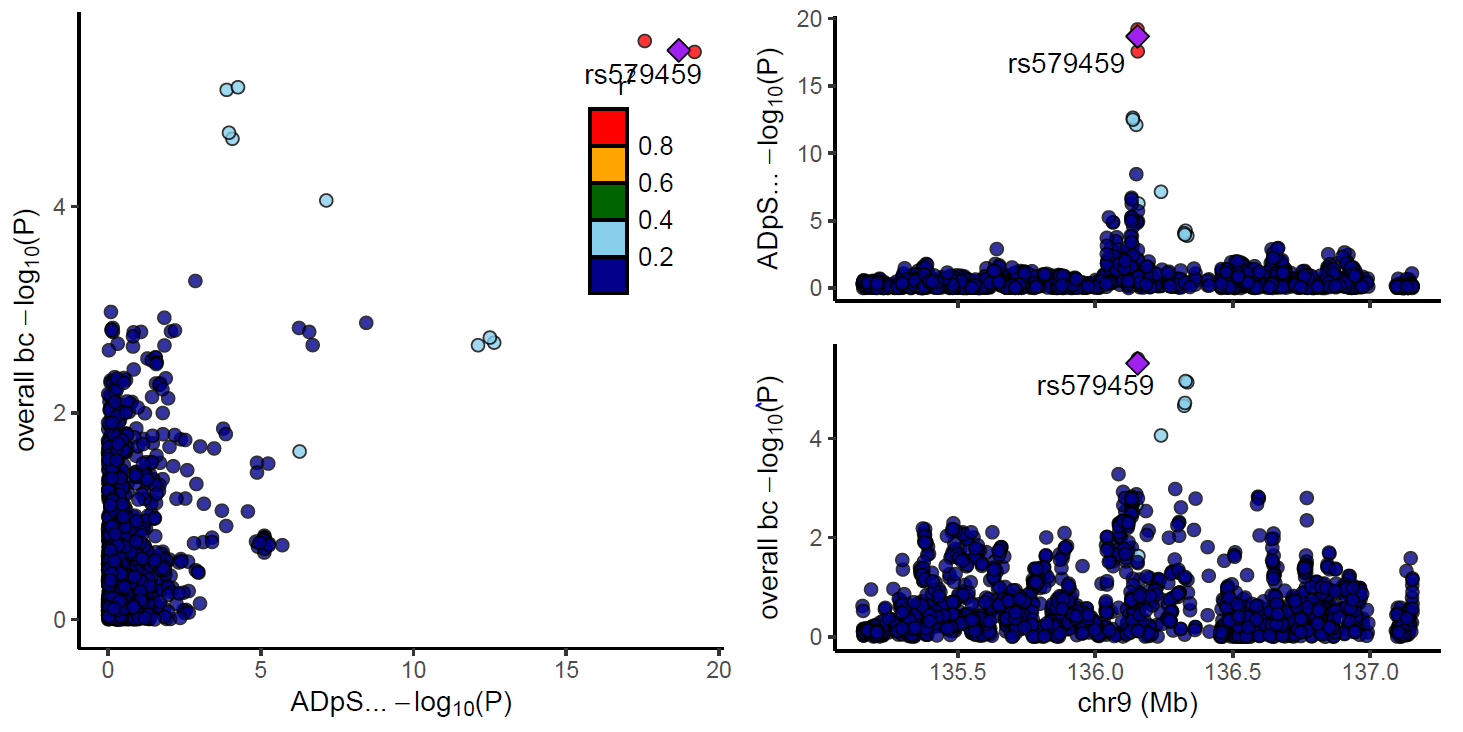

D)

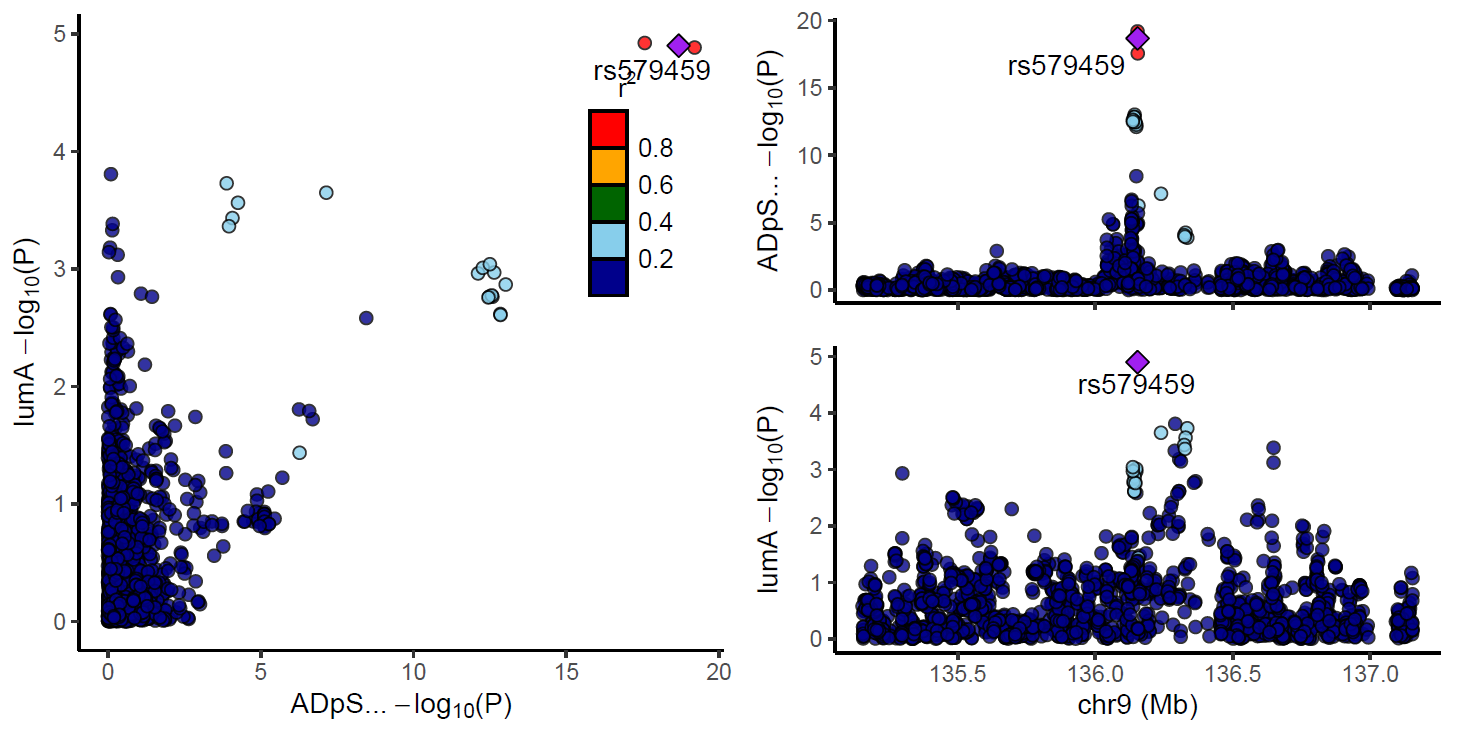

E)

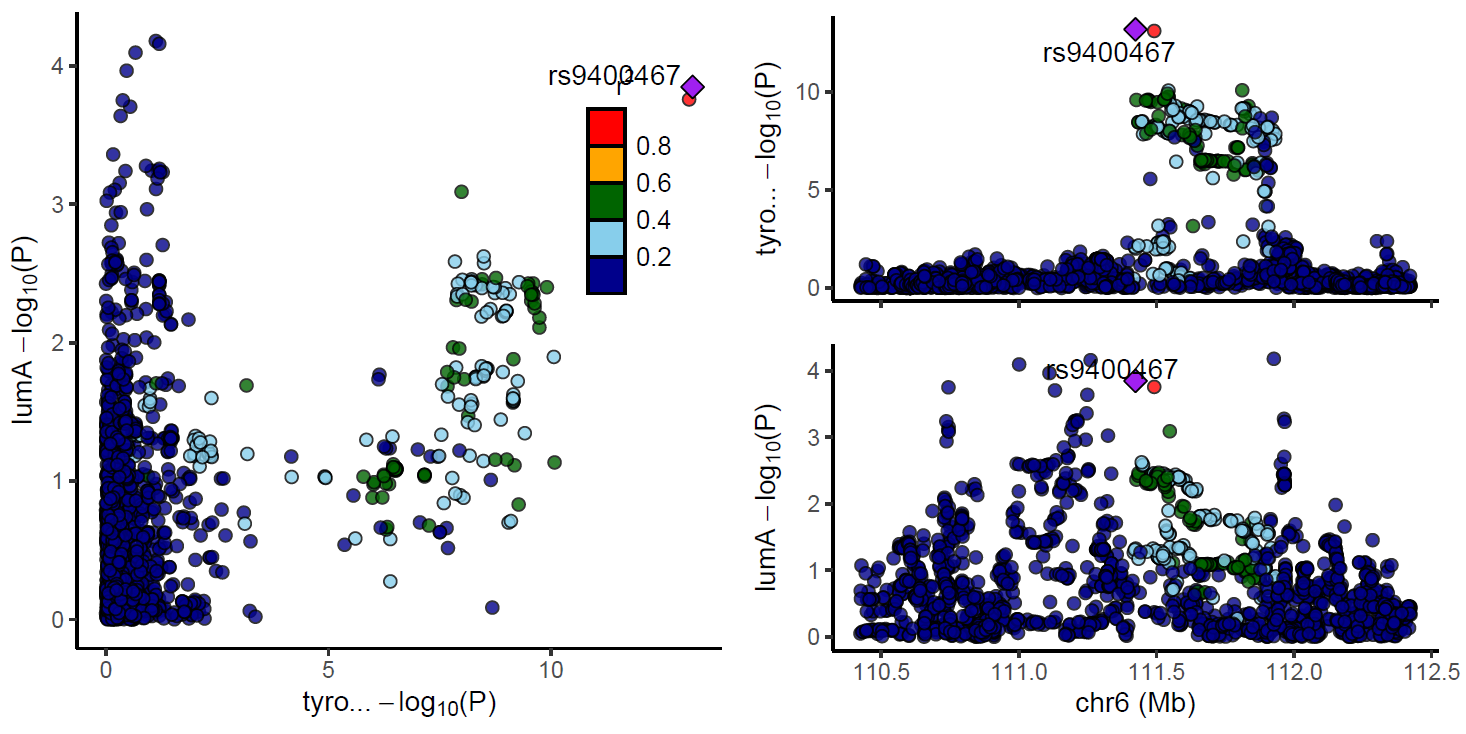

*bc = breast cancer; lumA = Luminal A; metab = metabolite. Regional association plots were created using summary-level data from the Shin et al. genome-wide association study and Breast Cancer Association Consortium or the Consortium of Investigation of Modifiers of BRCA1/2 with the LocusCompareR package. The -log10 p-value of each lead SNP is shown by a mauve diamond. Figures show colocalisation results for the (A) lead SNP, rs11950562, associated with homostachydrine and overall breast cancer risk; (B) same lead SNP, rs11950562, associated with homostachydrine and Luminal A breast cancer risk; (C) lead SNP, rs579459, associated with the ratio between ADpSGEGDFXAEGGGVR and X-14304—leucylalanine and overall breast cancer risk; (D) same lead SNP, rs579459, associated with the ratio between ADpSGEGDFXAEGGGVR and X-14304—leucylalanine and Luminal A breast cancer risk; and (E) lead SNP, rs9400467, associated with tyrosine and Luminal A breast cancer.*
