## Supplementary Figure 3 for "Exploring the causal role of the human gut microbiome in breast cancer risk"

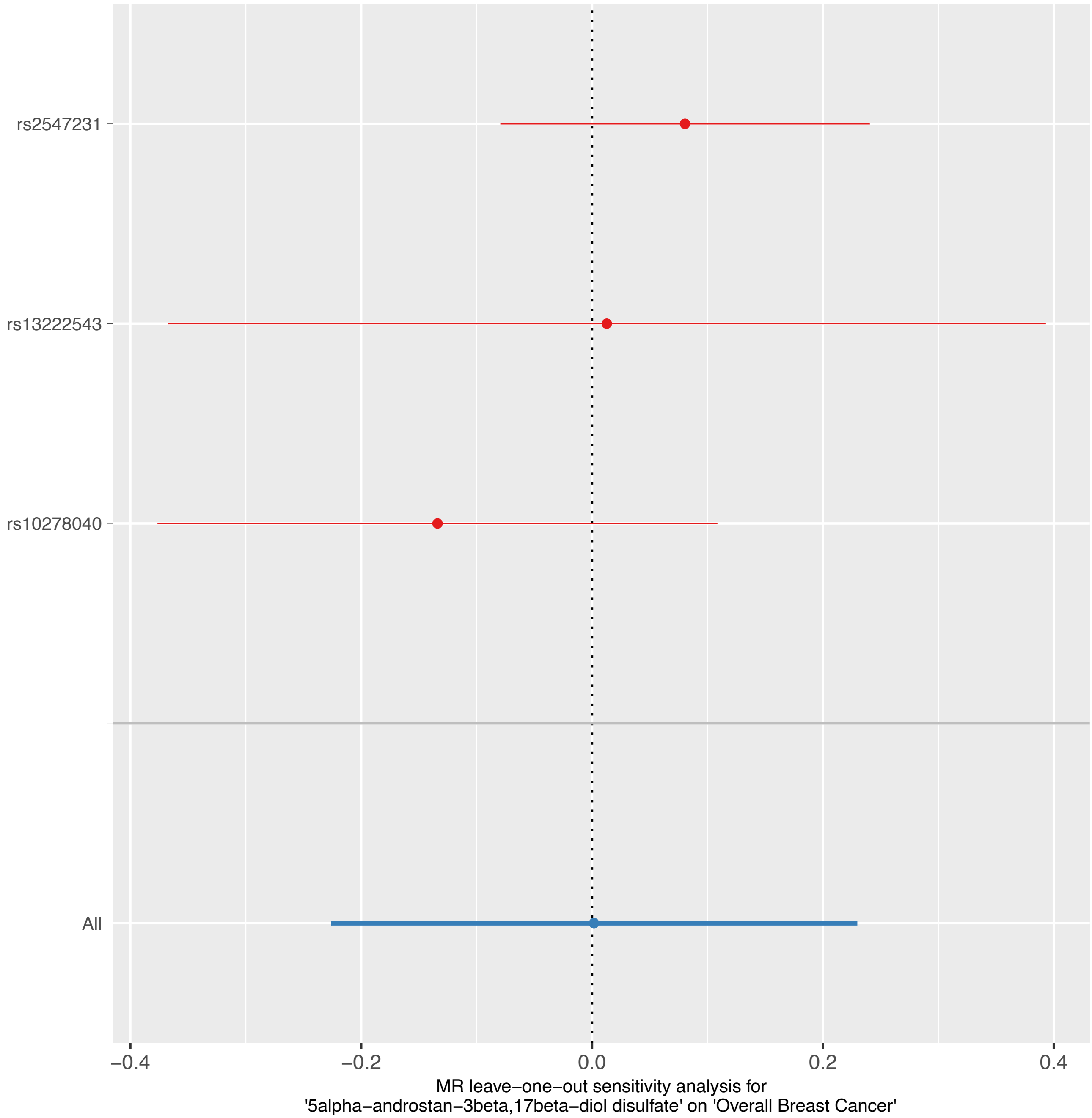

rs2547231
rs13222543
rs10278040
All
−0.4
−0.2
0.0
0.2
0.4
MR leave−one−out sensitivity analysis for
'5alpha−androstan−3beta,17beta−diol disulfate' on 'Overall Breast Cancer'

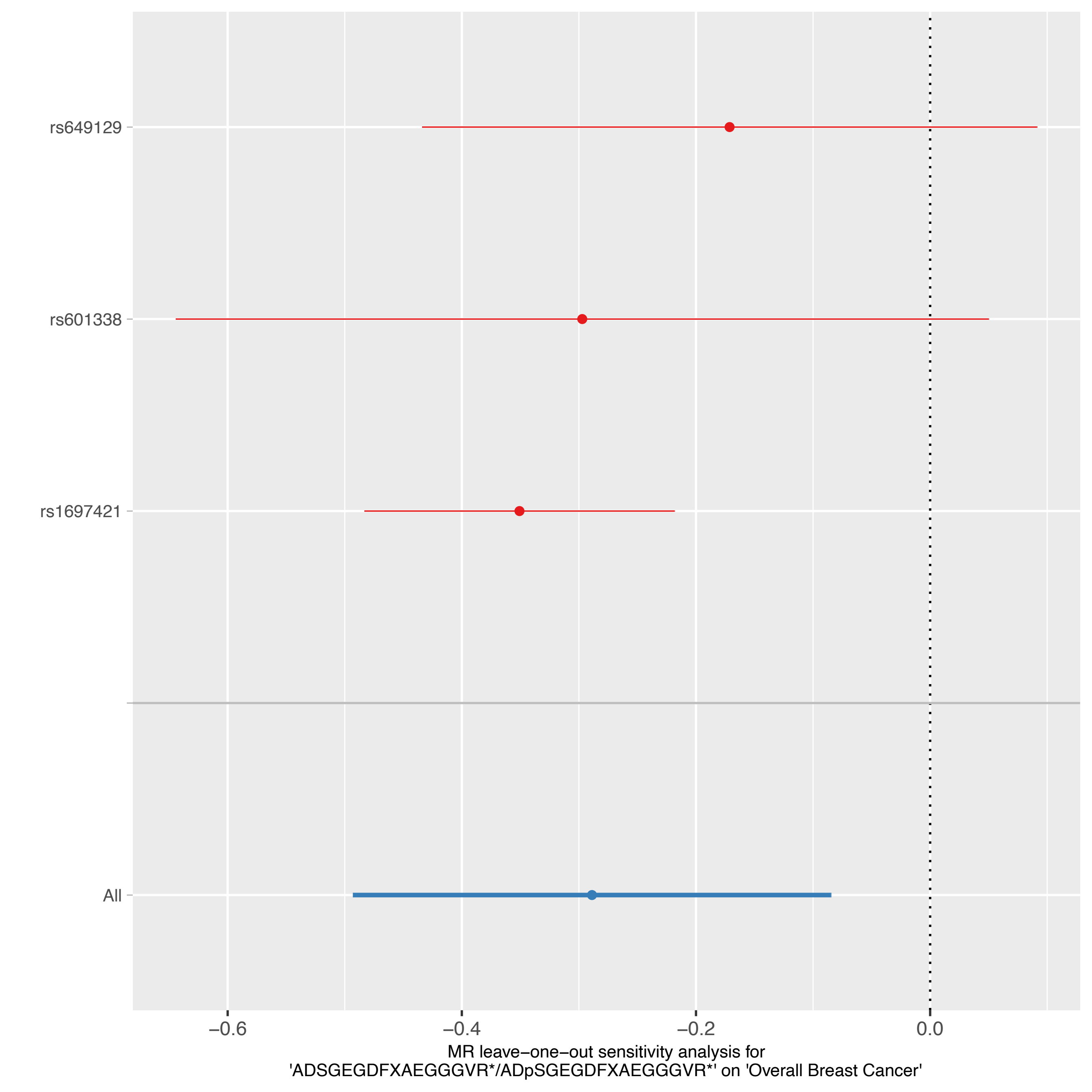

rs649129
rs601338
rs1697421
All
−0.6
−0.4
−0.2
0.0
MR leave−one−out sensitivity analysis for
'ADSGEGDFXAEGGGVR*/ADpSGEGDFXAEGGGVR*' on 'Overall Breast Cancer'

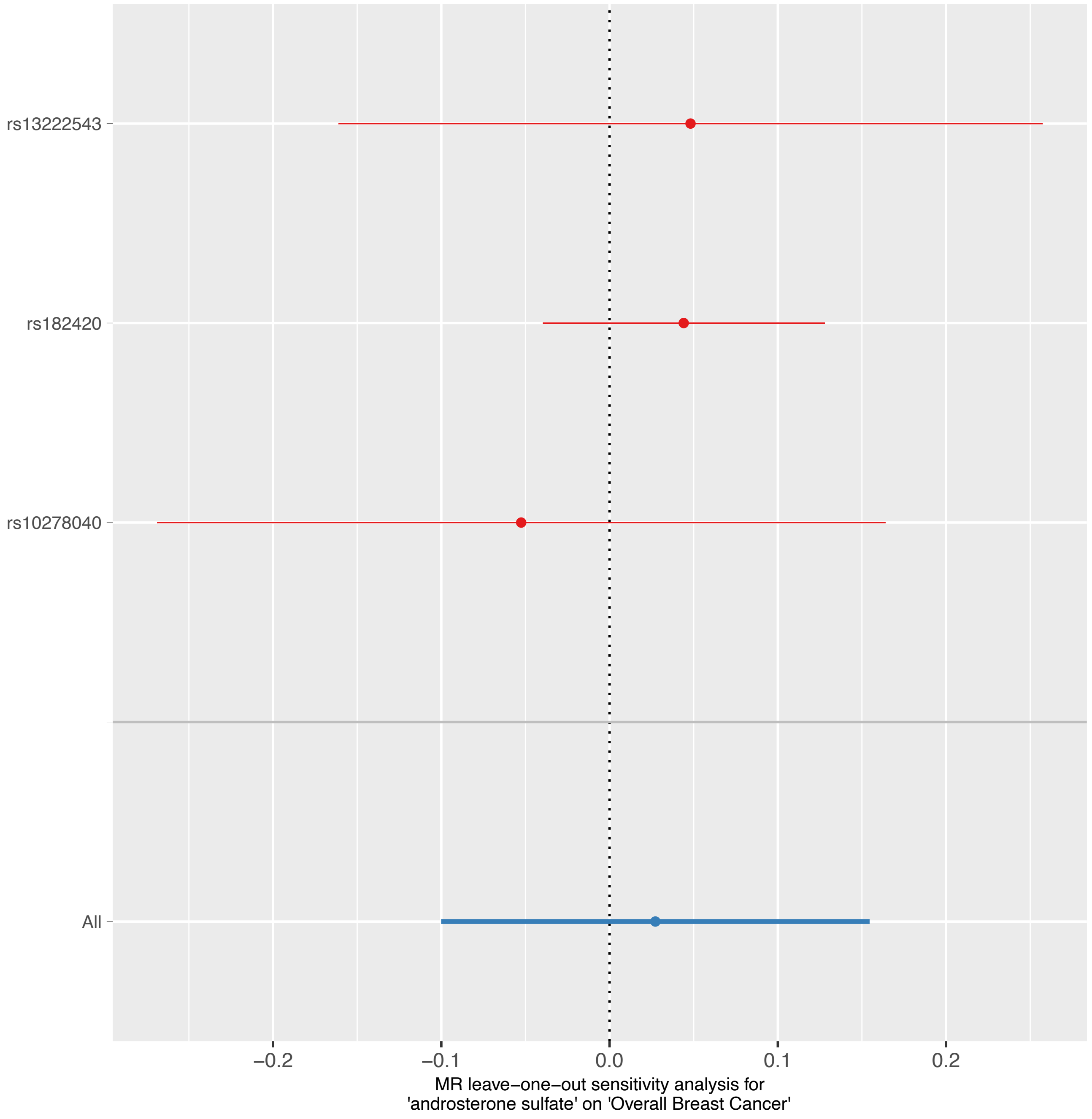

rs13222543
rs182420
rs10278040
All
−0.2
−0.1
0.0
0.1
0.2
MR leave−one−out sensitivity analysis for
'androsterone sulfate' on 'Overall Breast Cancer'

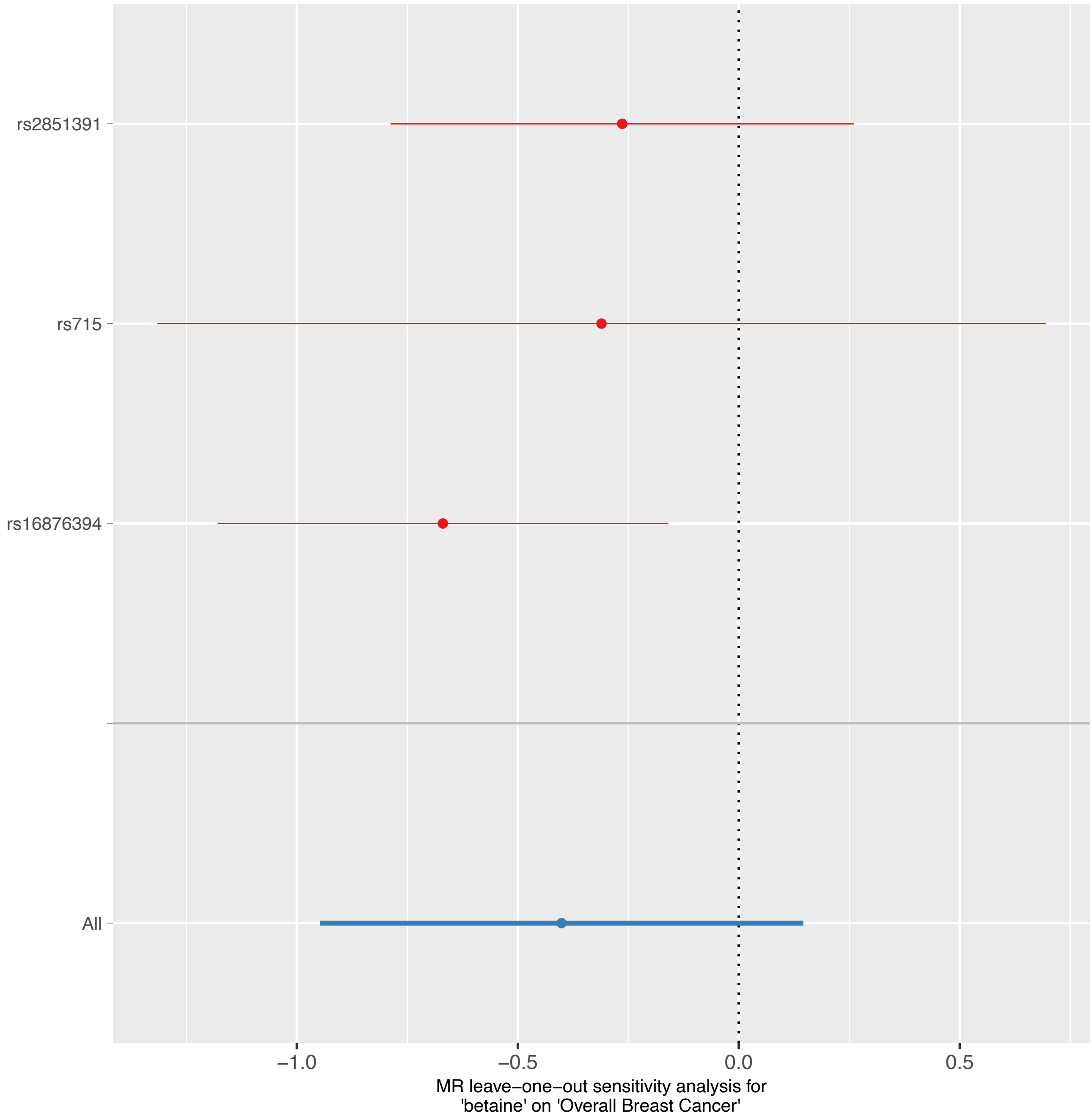
rs2851391
rs715
rs16876394
All
−1.0
−0.5
0.0
0.5
MR leave−one−out sensitivity analysis for
'betaine' on 'Overall Breast Cancer'

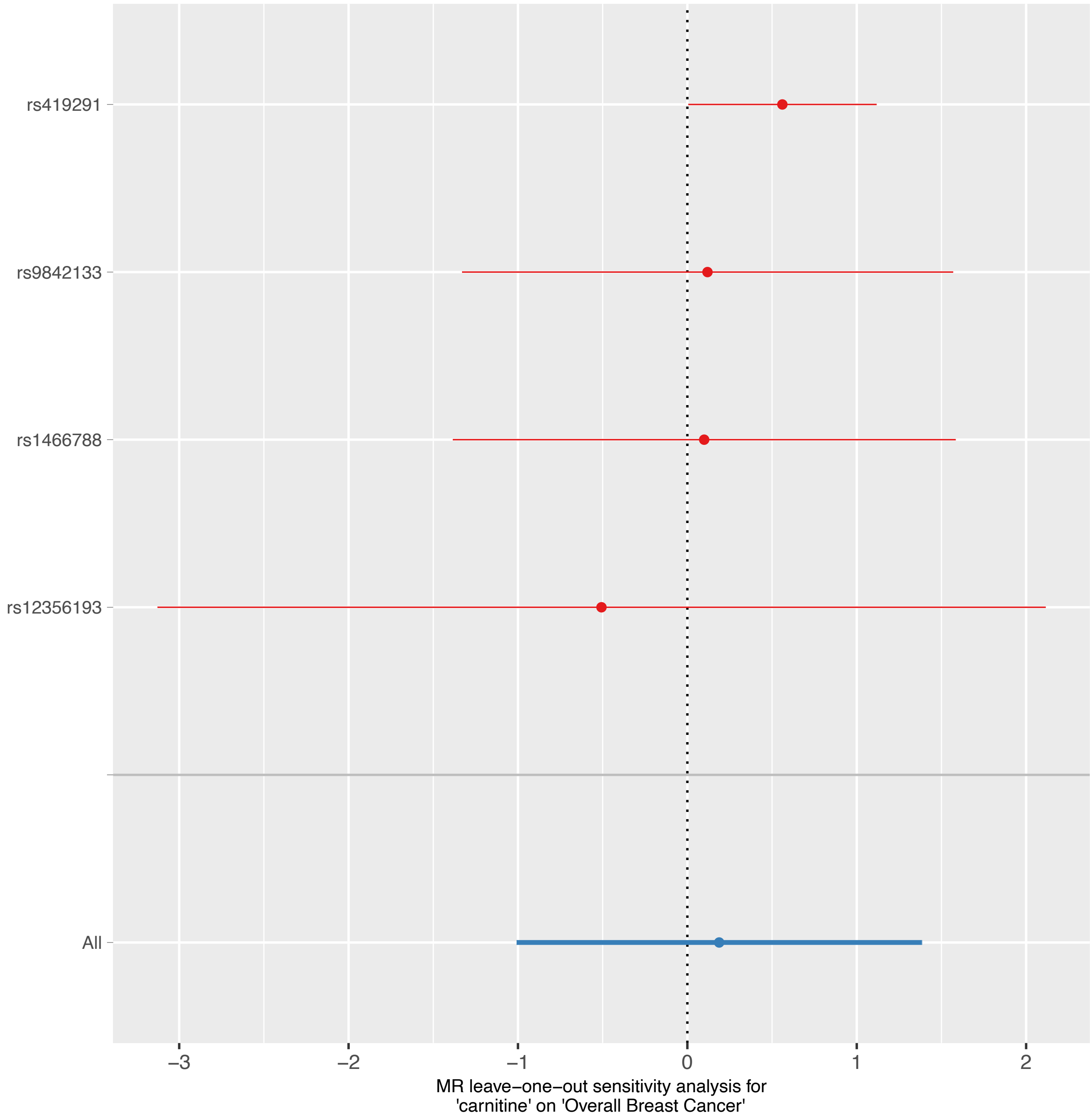

rs419291
rs9842133
rs1466788
rs12356193
All
−3
−2
−1
0
1
2
MR leave−one−out sensitivity analysis for
'carnitine' on 'Overall Breast Cancer'

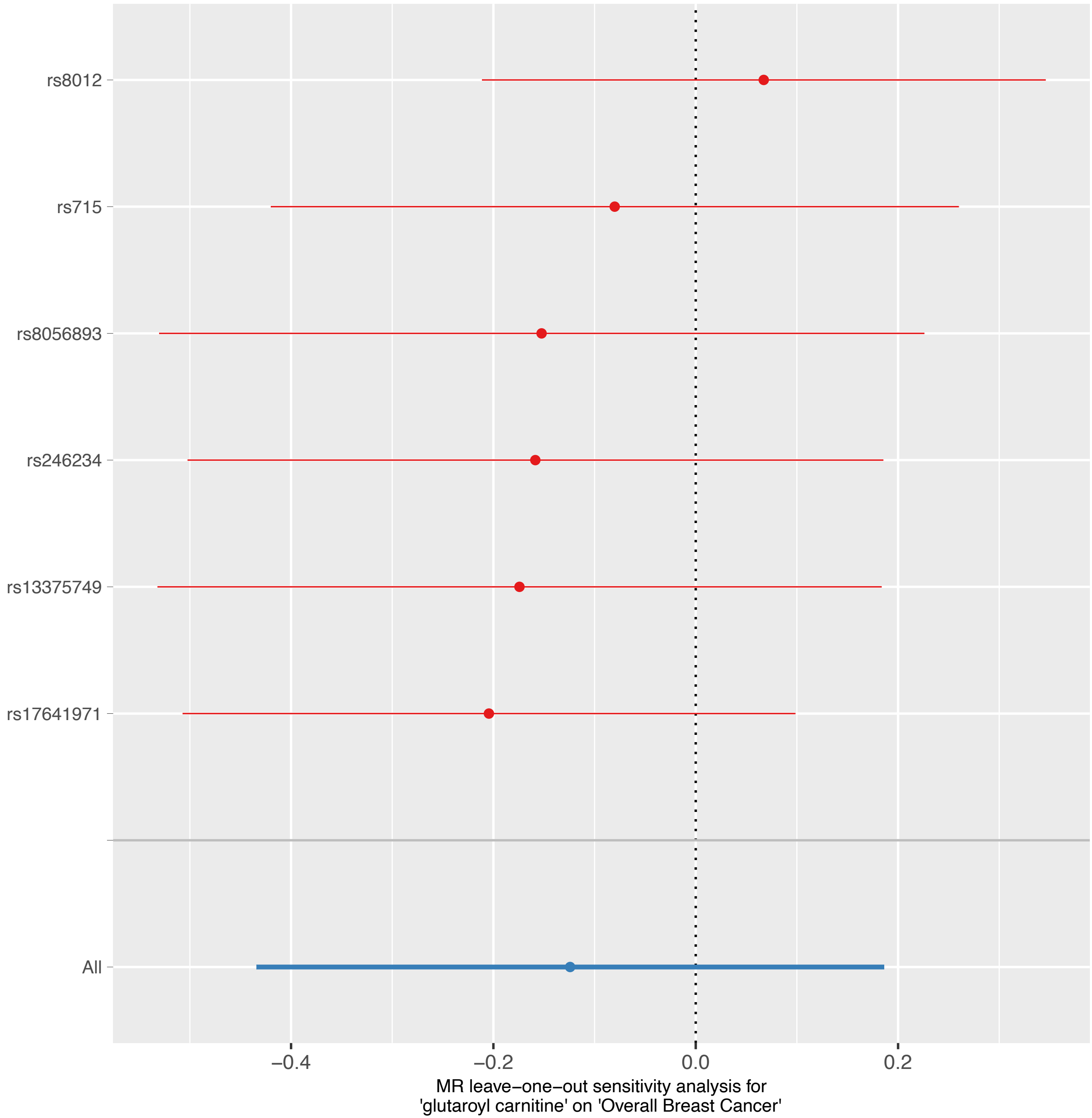
rs8012
rs715
rs8056893
rs246234
rs13375749
rs17641971
All
−0.4
−0.2
0.0
0.2
MR leave−one−out sensitivity analysis for
'glutaroyl carnitine' on 'Overall Breast Cancer'

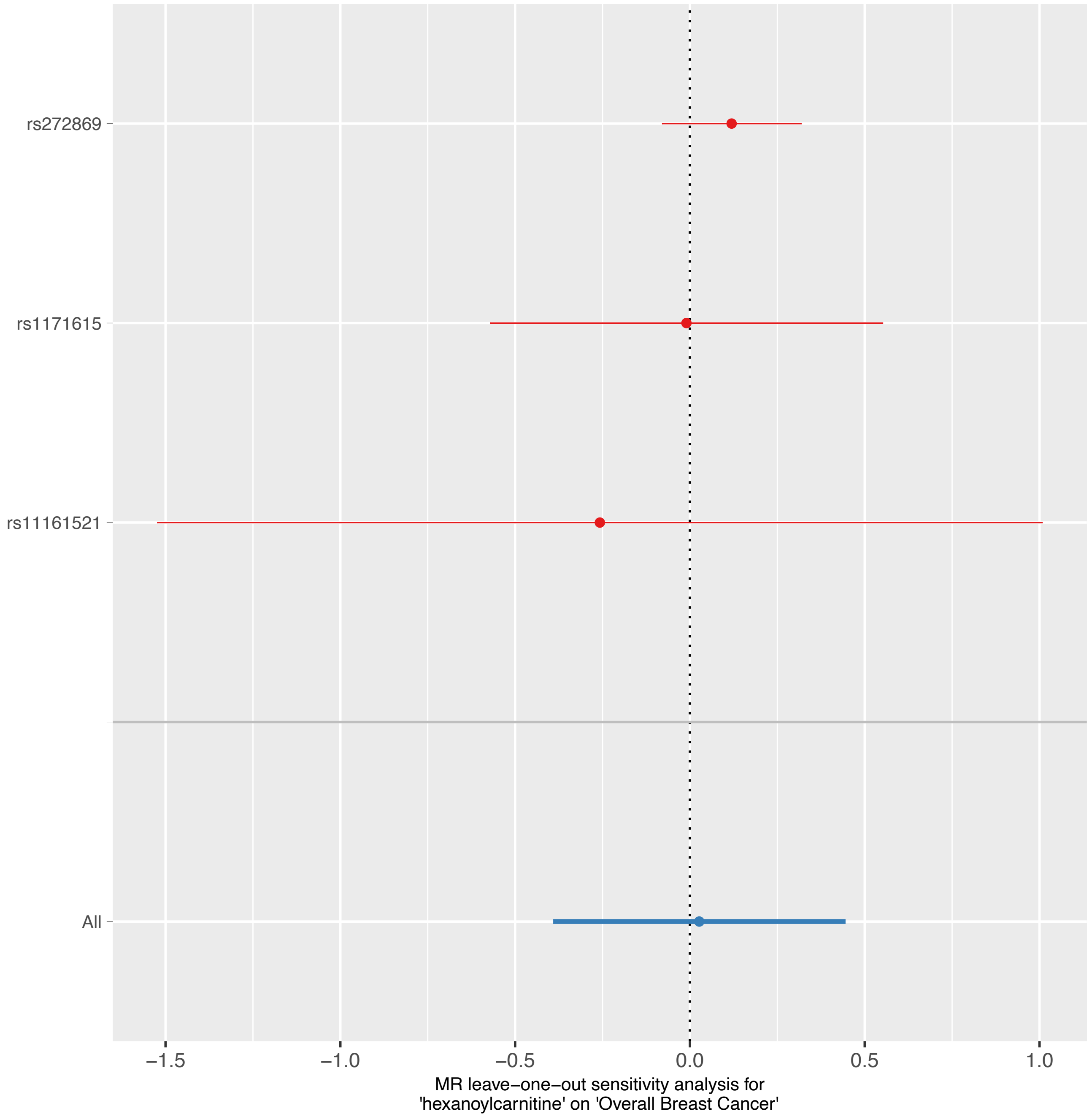
rs272869
rs1171615
rs11161521
All
−1.5
−1.0
−0.5
0.0
0.5
1.0
MR leave−one−out sensitivity analysis for
'hexanoylcarnitine' on 'Overall Breast Cancer'

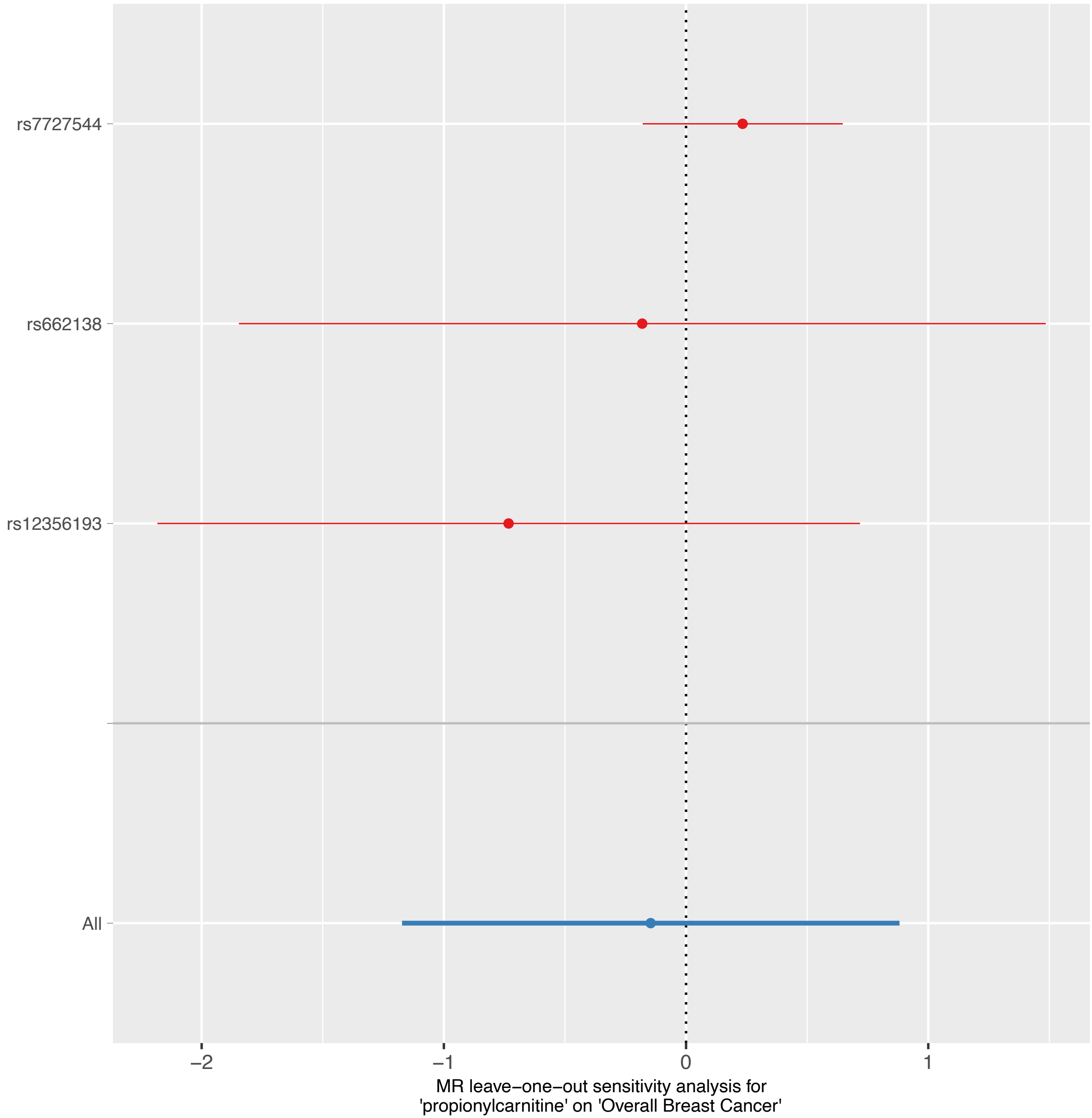

rs7727544
rs662138
rs12356193
All
−2
−1
0
1
MR leave−one−out sensitivity analysis for
'propionylcarnitine' on 'Overall Breast Cancer'

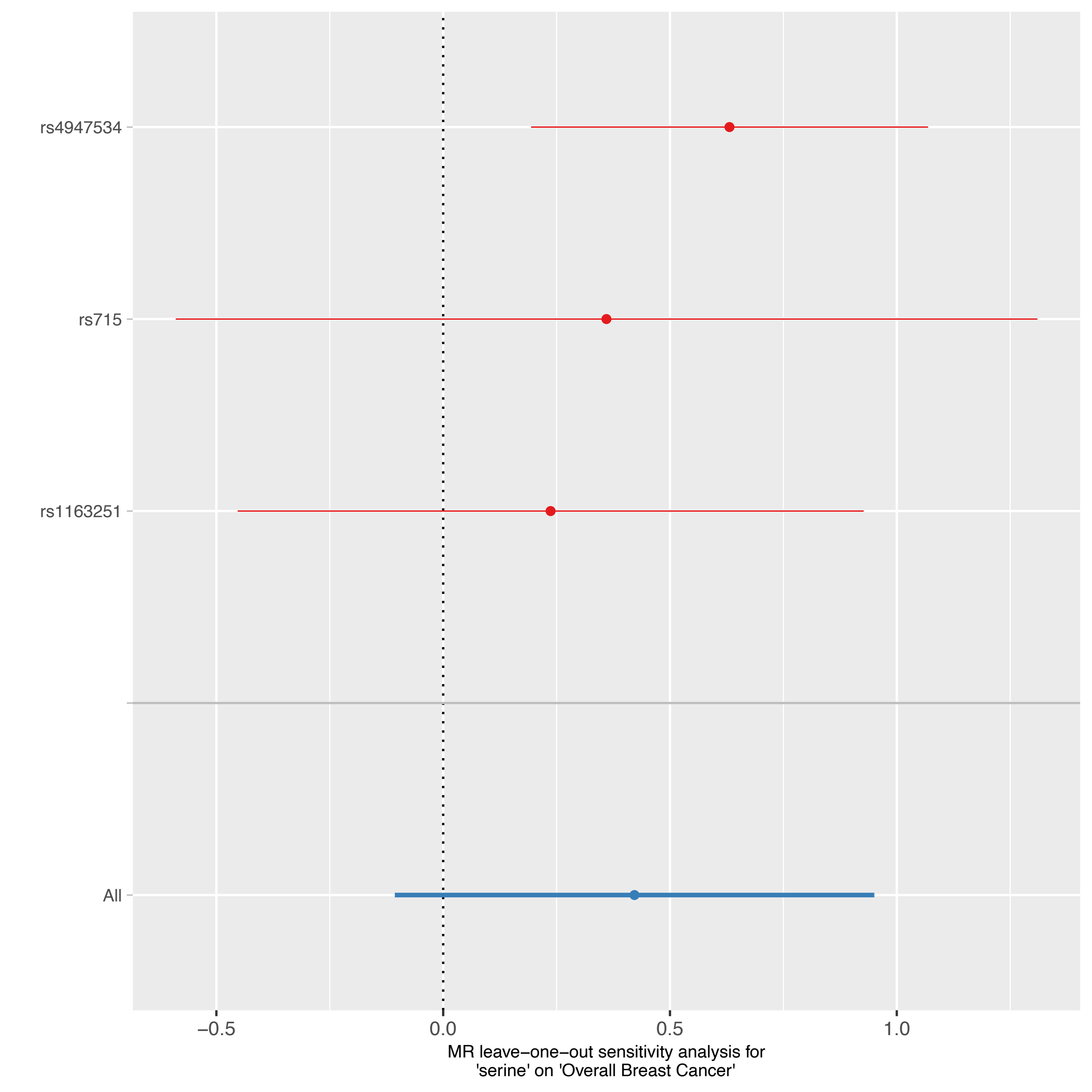

rs4947534
rs715
rs1163251
All
−0.5
0.0
0.5
1.0
MR leave−one−out sensitivity analysis for
'serine' on 'Overall Breast Cancer'

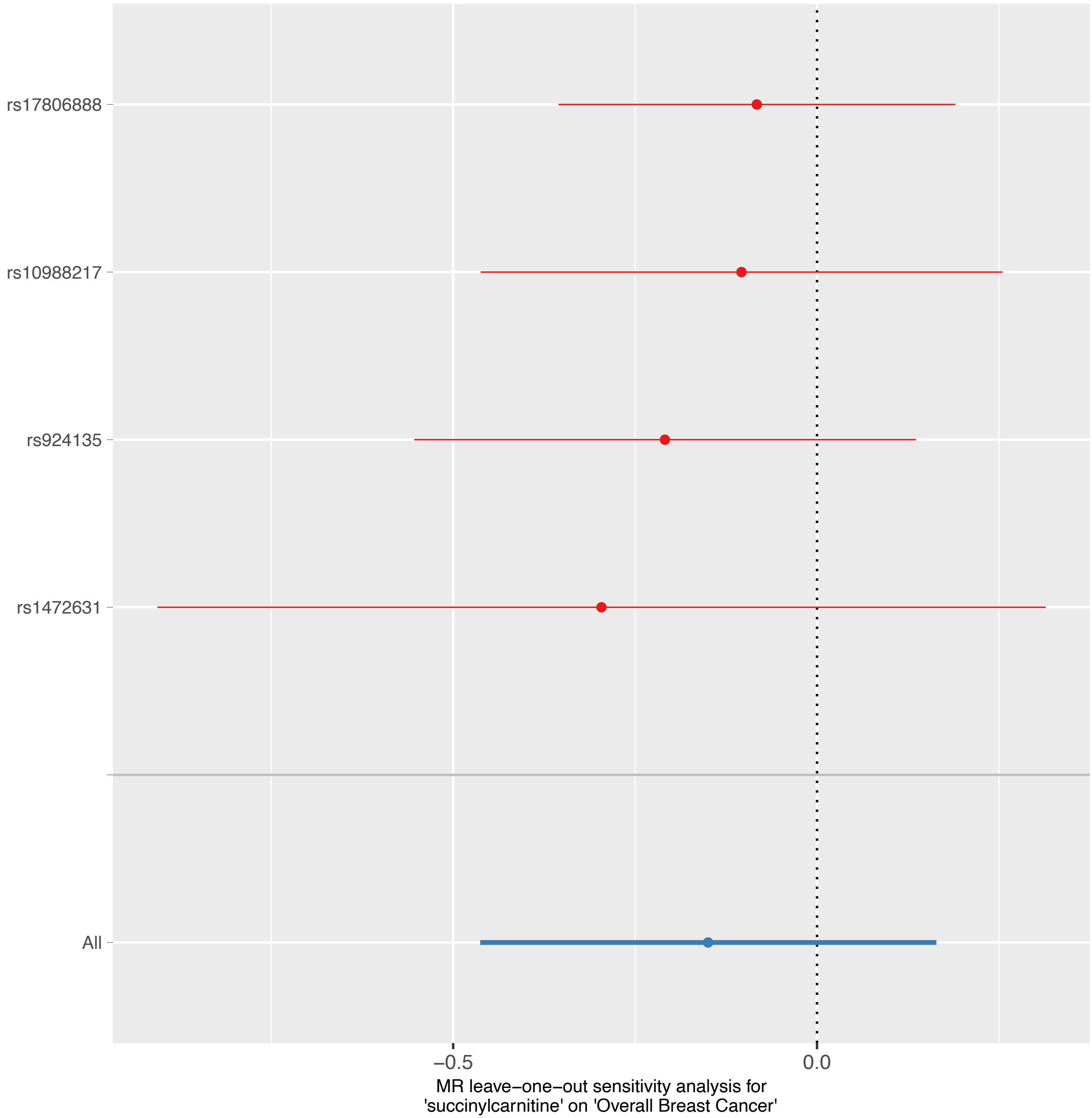
rs17806888
rs10988217
rs924135
rs1472631
All
−0.5
0.0
MR leave−one−out sensitivity analysis for
'succinylcarnitine' on 'Overall Breast Cancer'

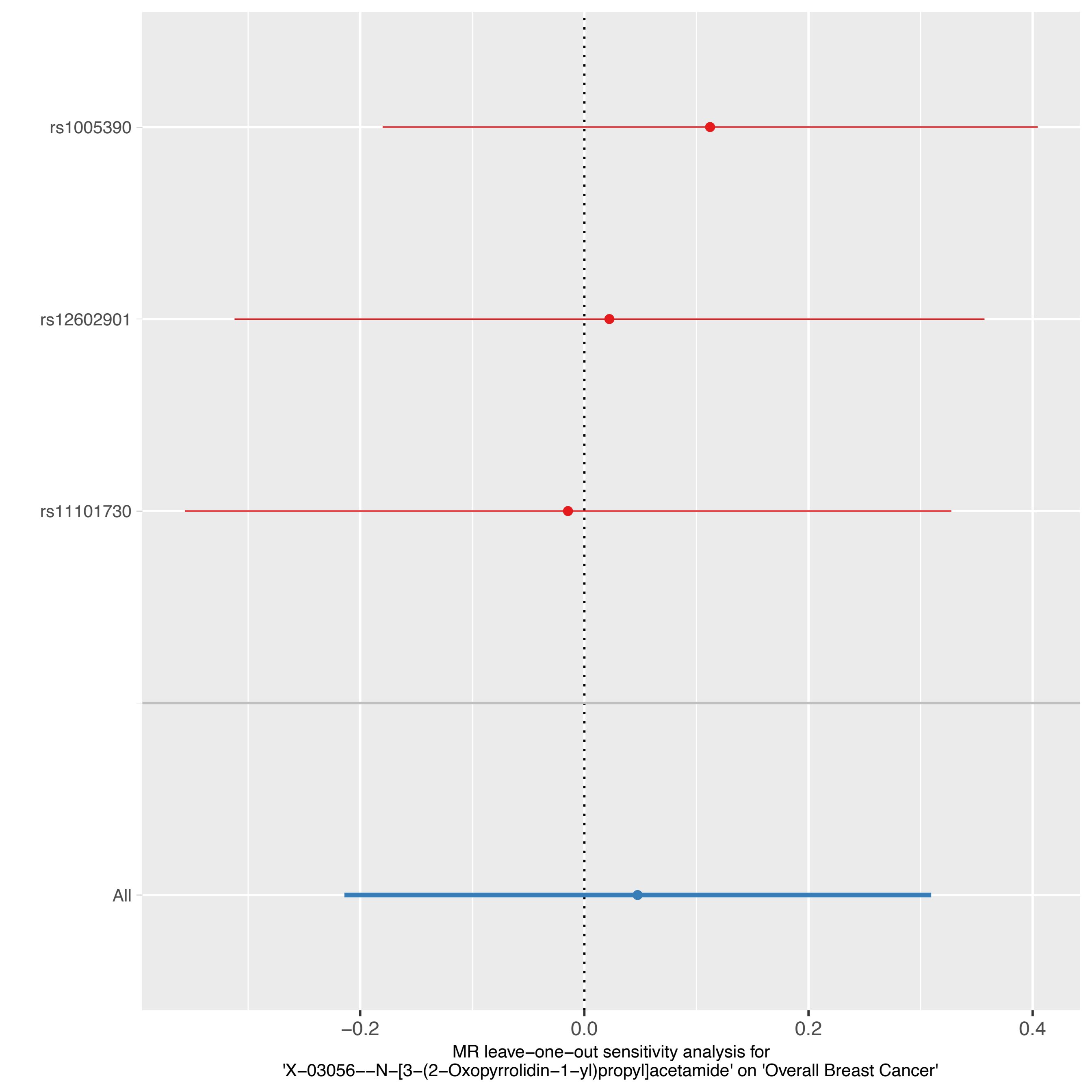

rs1005390
rs12602901
rs11101730
All
−0.2
0.0
0.2
0.4
MR leave−one−out sensitivity analysis for
'X−03056−−N−[3−(2−Oxopyrrolidin−1−yl)propyl]acetamide' on 'Overall Breast Cancer'

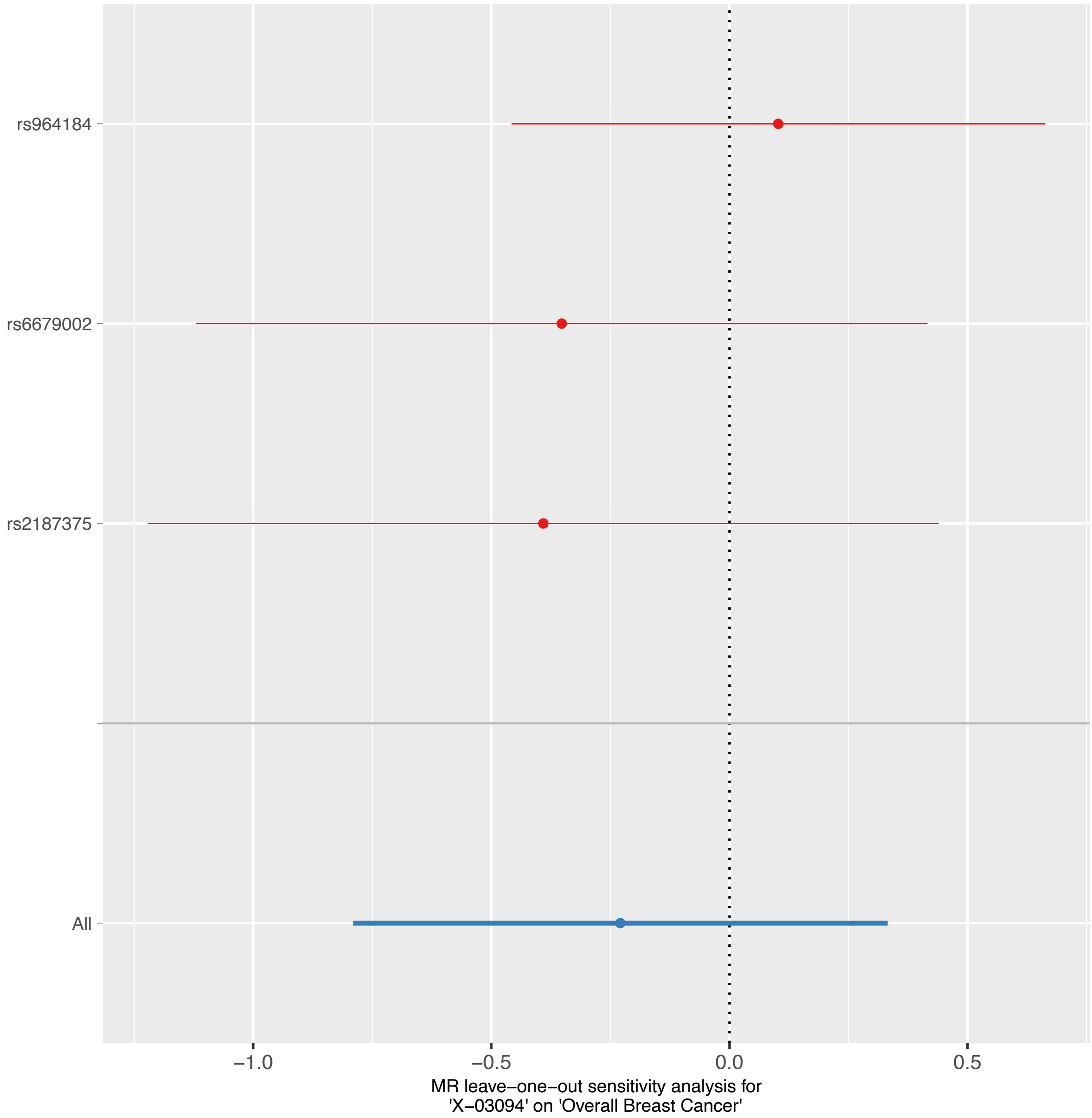

rs964184
rs6679002
rs2187375
All
−1.0
−0.5
0.0
0.5
MR leave−one−out sensitivity analysis for
'X−03094' on 'Overall Breast Cancer'

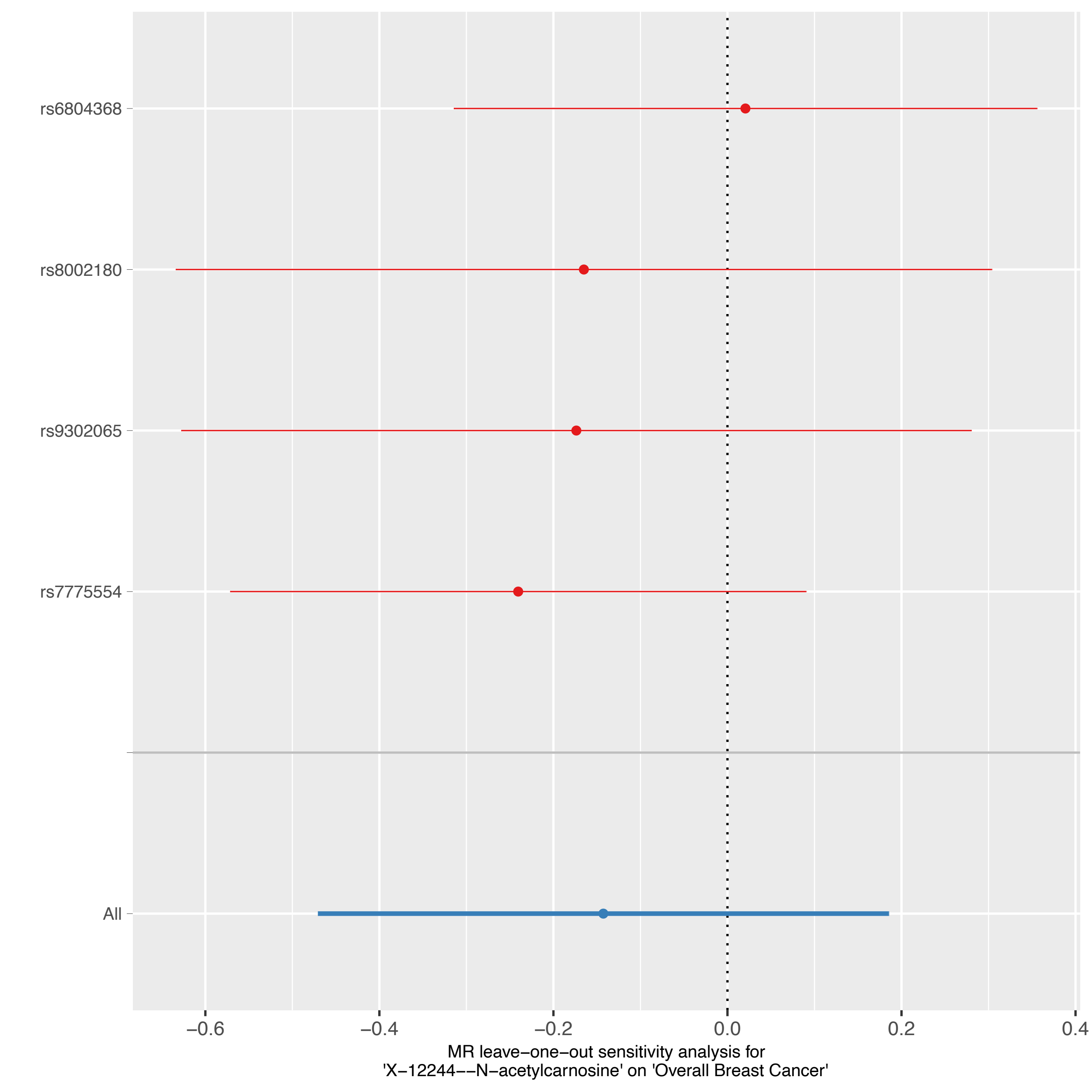
rs6804368
rs8002180
rs9302065
rs7775554
All
−0.6
−0.4
−0.2
0.0
0.2
0.4
MR leave−one−out sensitivity analysis for
'X−12244−−N−acetylcarnosine' on 'Overall Breast Cancer'

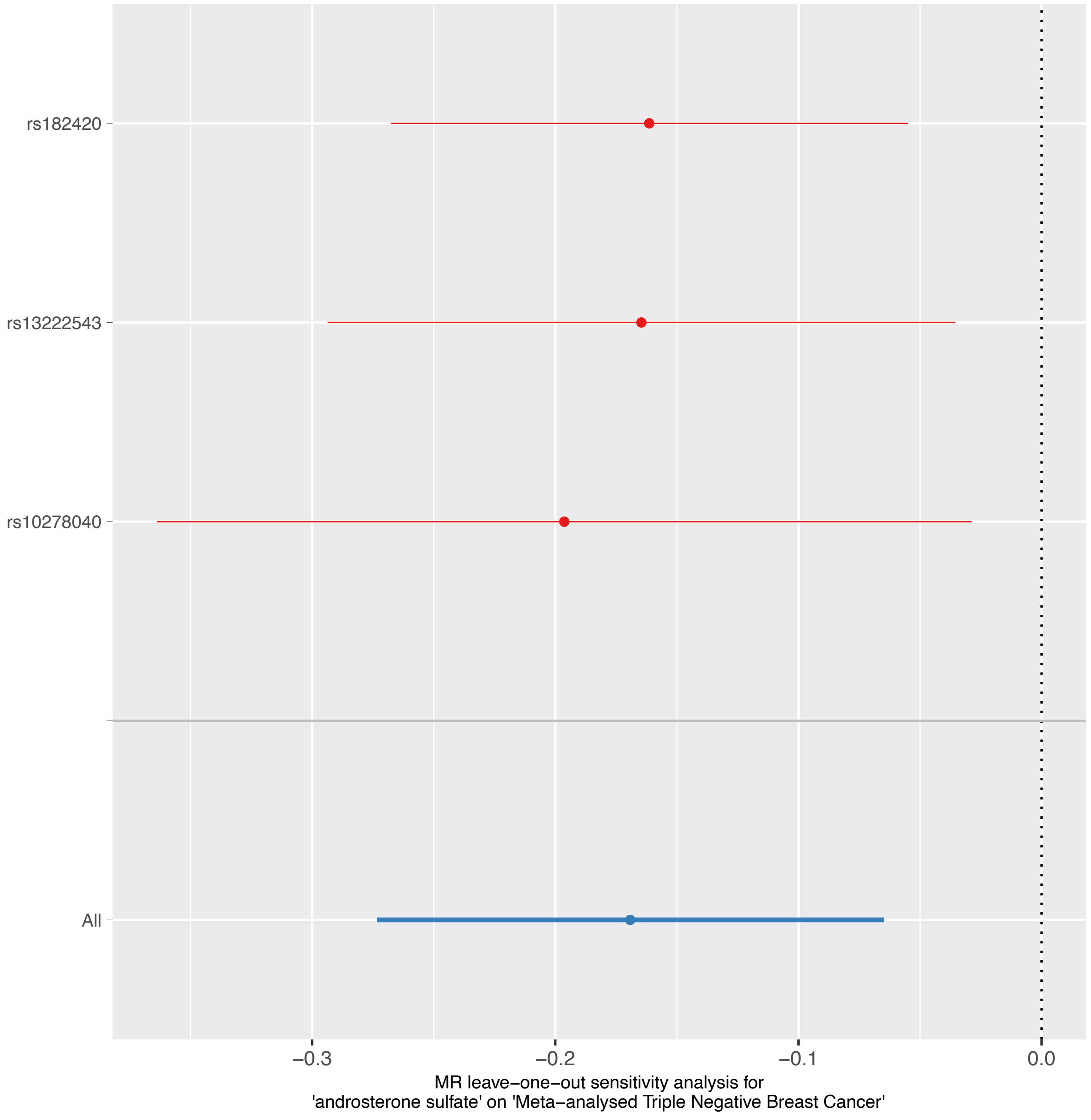

rs182420
rs13222543
rs10278040
All
−0.3
−0.2
−0.1
0.0
MR leave−one−out sensitivity analysis for
'androsterone sulfate' on 'Meta−analysed Triple Negative Breast Cancer'

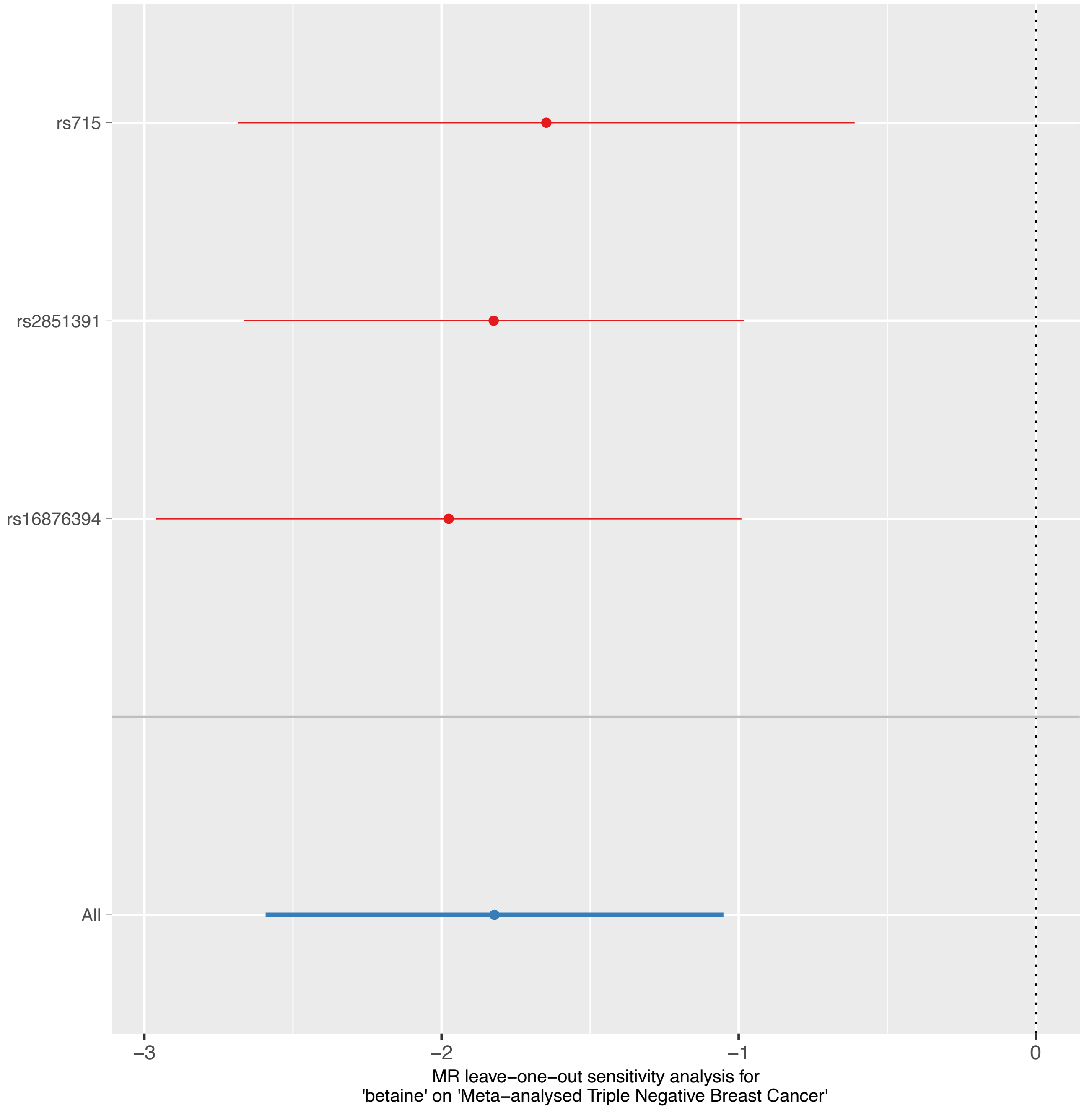
rs715
rs2851391
rs16876394
All
−3
−2
−1
0
MR leave−one−out sensitivity analysis for
'betaine' on 'Meta−analysed Triple Negative Breast Cancer'

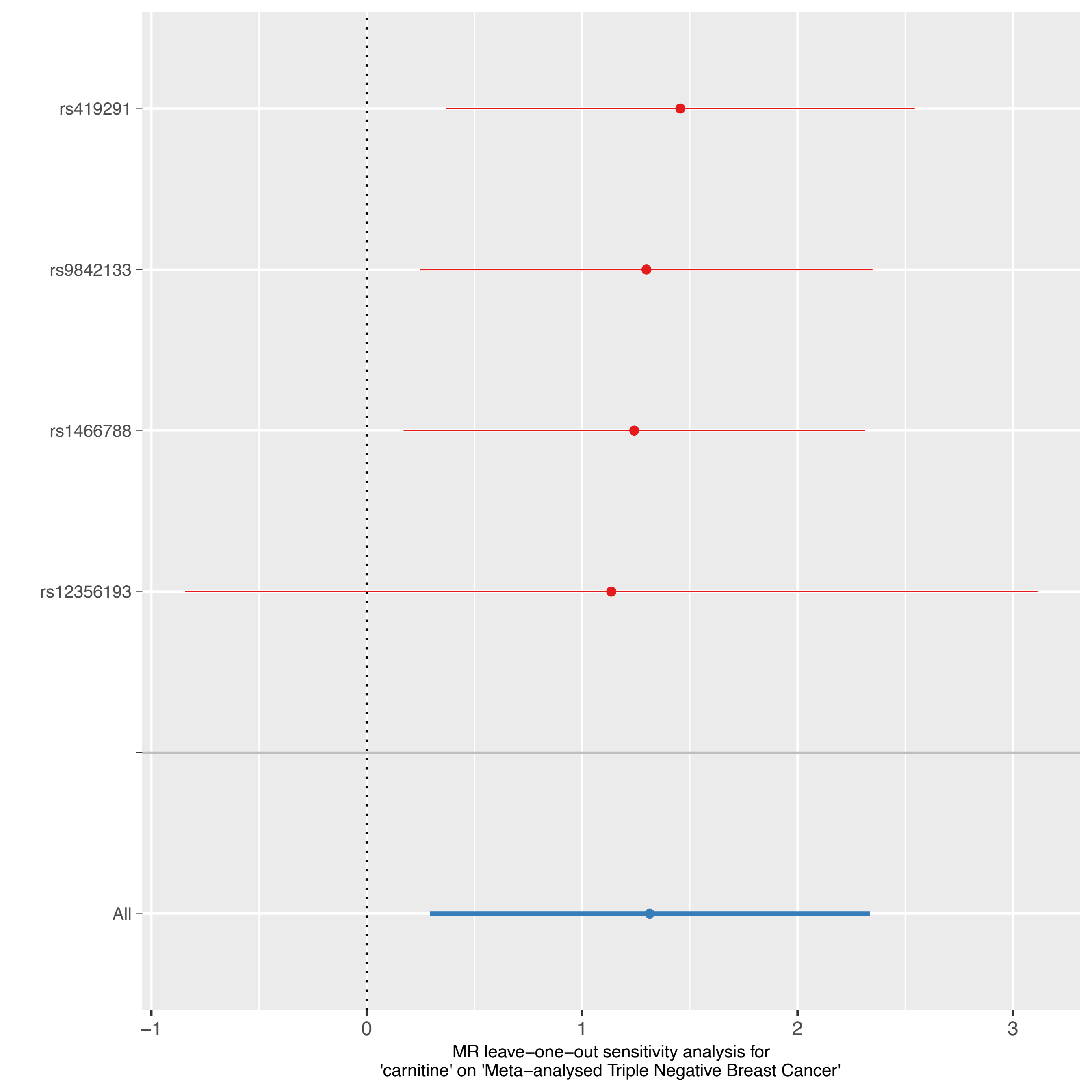

rs419291
rs9842133
rs1466788
rs12356193
All
−1
0
1
2
3
MR leave−one−out sensitivity analysis for
'carnitine' on 'Meta−analysed Triple Negative Breast Cancer'

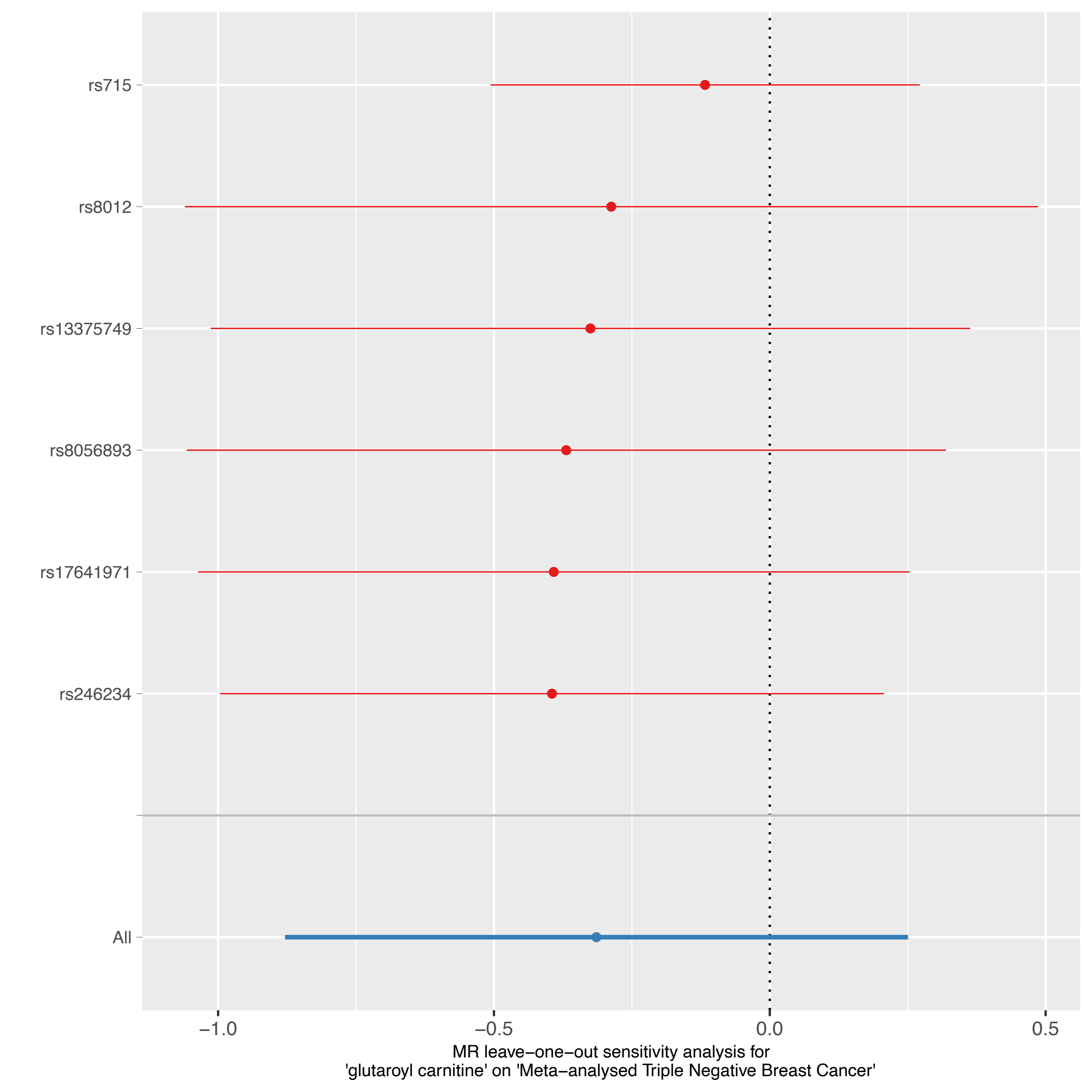
rs715
rs8012
rs13375749
rs8056893
rs17641971
rs246234
All
−1.0
−0.5
0.0
0.5
MR leave−one−out sensitivity analysis for
'glutaroyl carnitine' on 'Meta−analysed Triple Negative Breast Cancer'

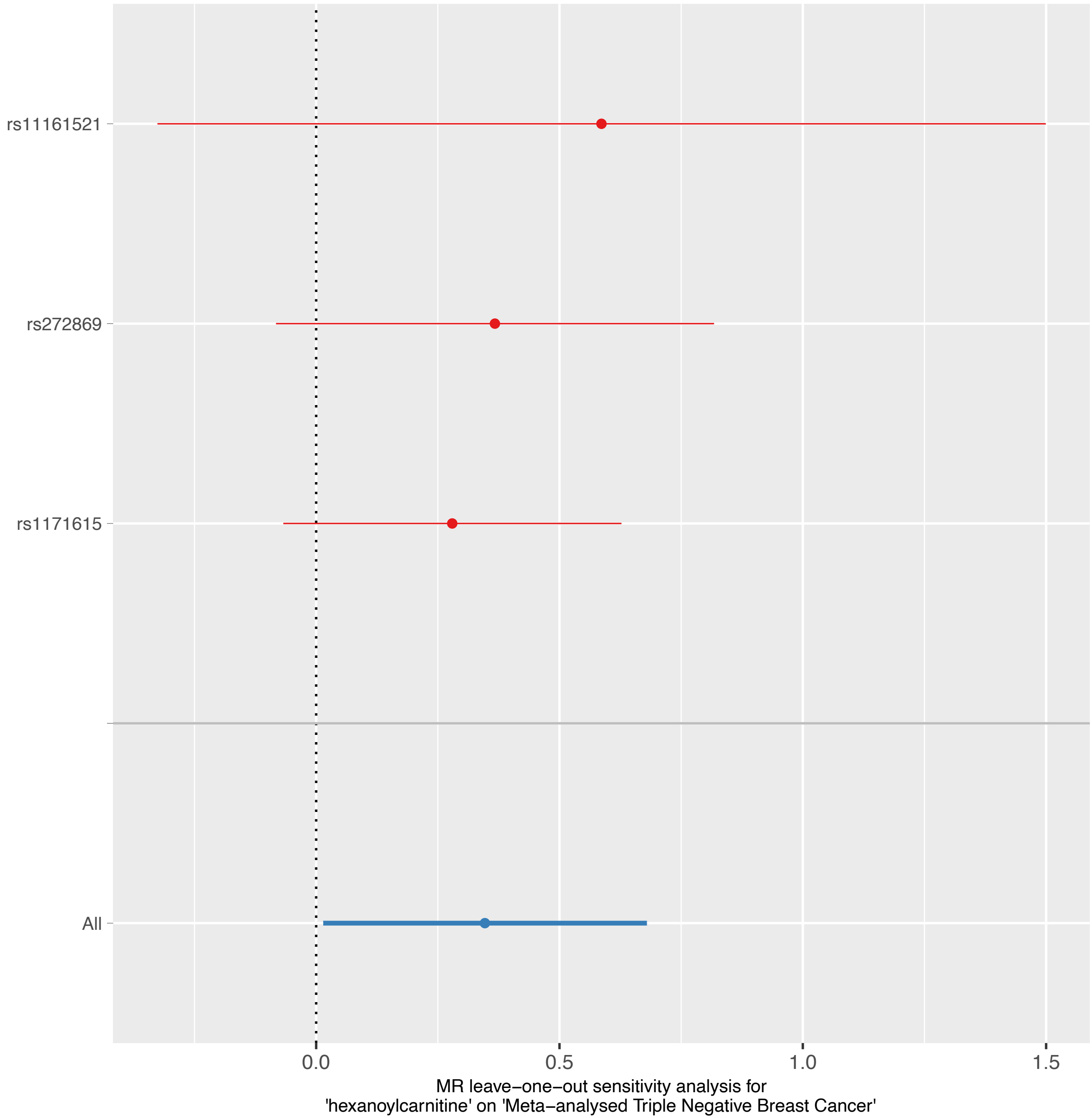
rs11161521
rs272869
rs1171615
All
0.0
0.5
1.0
1.5
MR leave−one−out sensitivity analysis for
'hexanoylcarnitine' on 'Meta−analysed Triple Negative Breast Cancer'

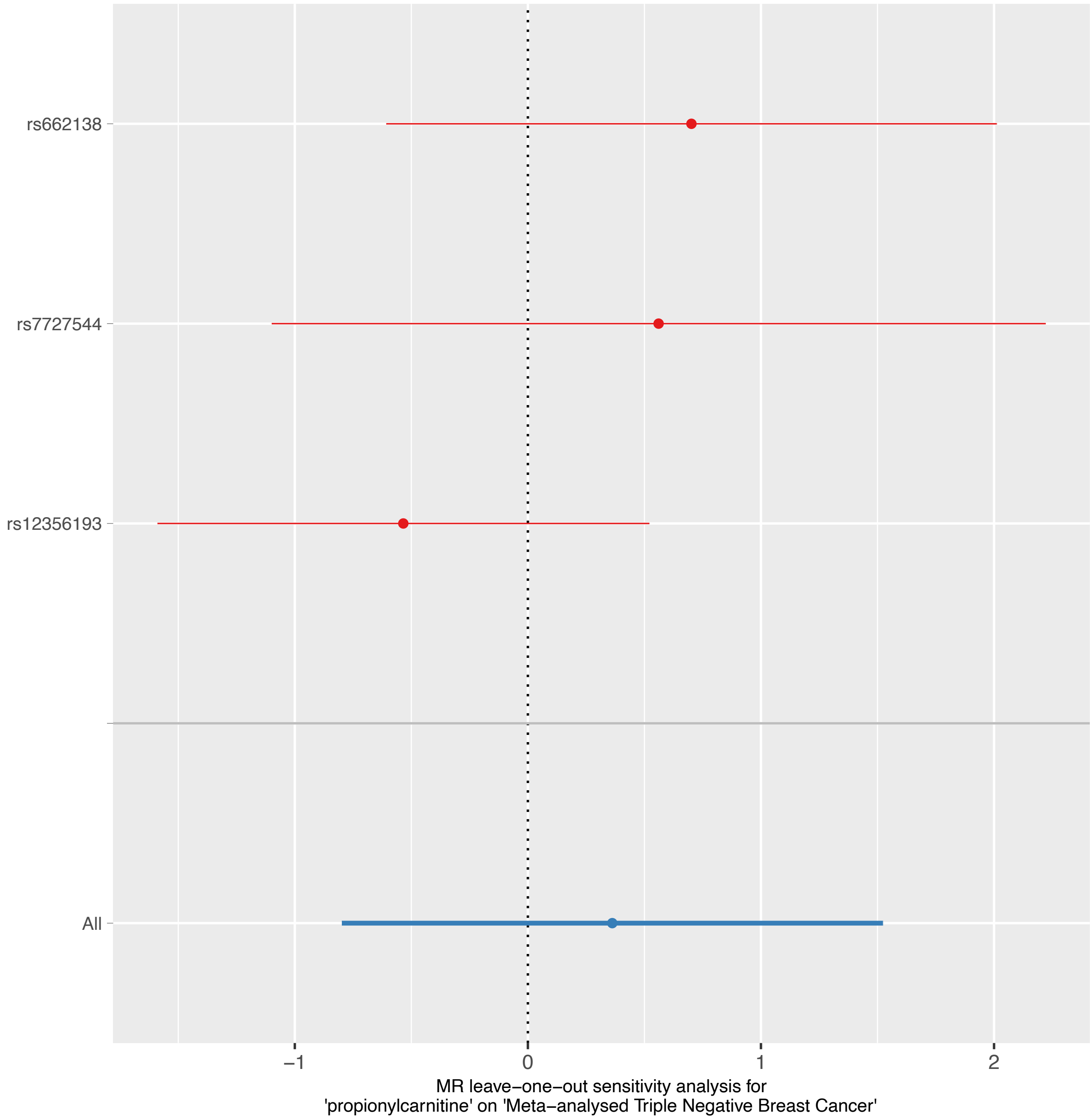

rs662138
rs7727544
rs12356193
All
−1
0
1
2
MR leave−one−out sensitivity analysis for
'propionylcarnitine' on 'Meta−analysed Triple Negative Breast Cancer'

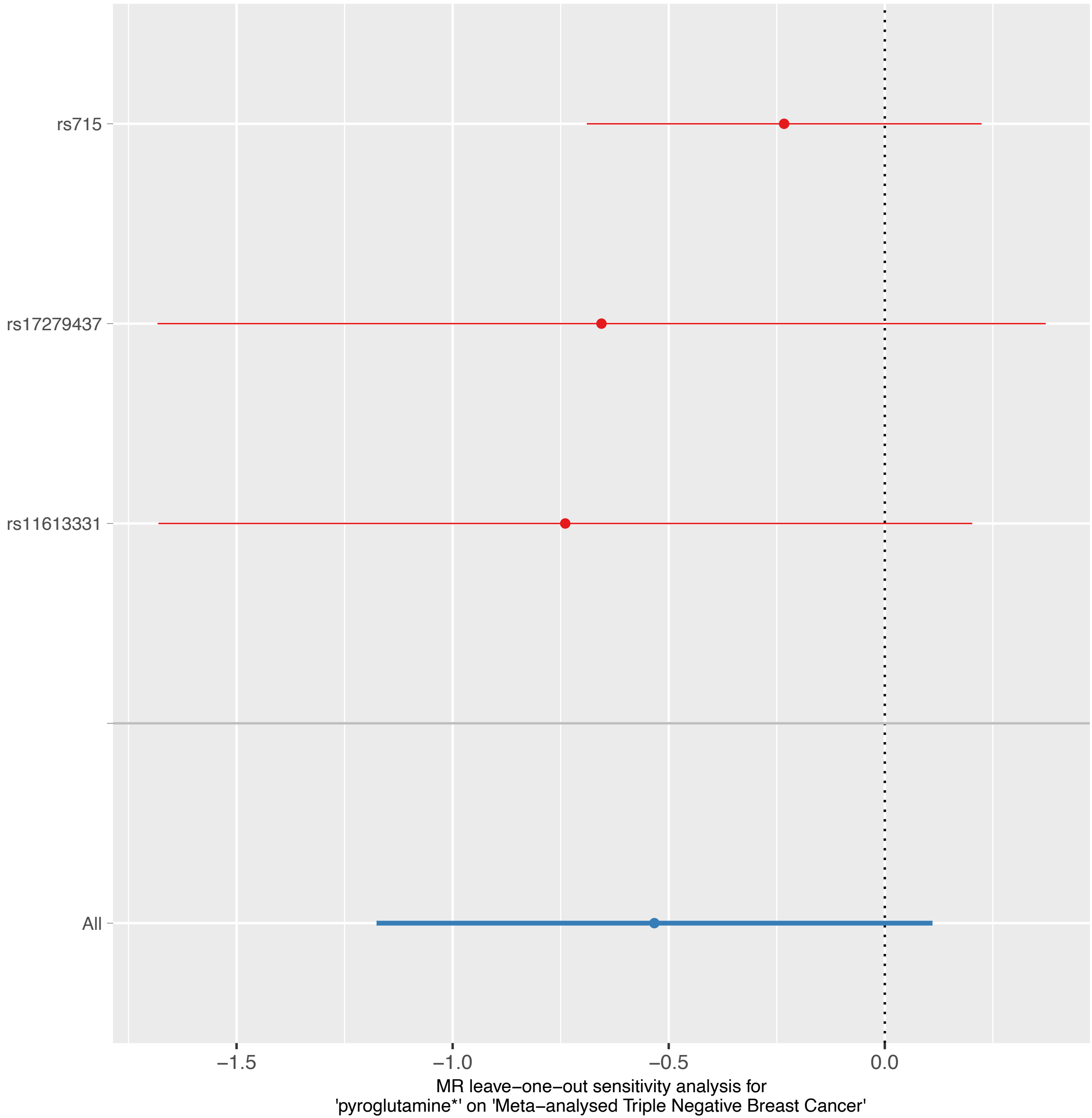
rs715
rs17279437
rs11613331
All
−1.5
−1.0
−0.5
0.0
MR leave−one−out sensitivity analysis for
'pyroglutamine*' on 'Meta−analysed Triple Negative Breast Cancer'

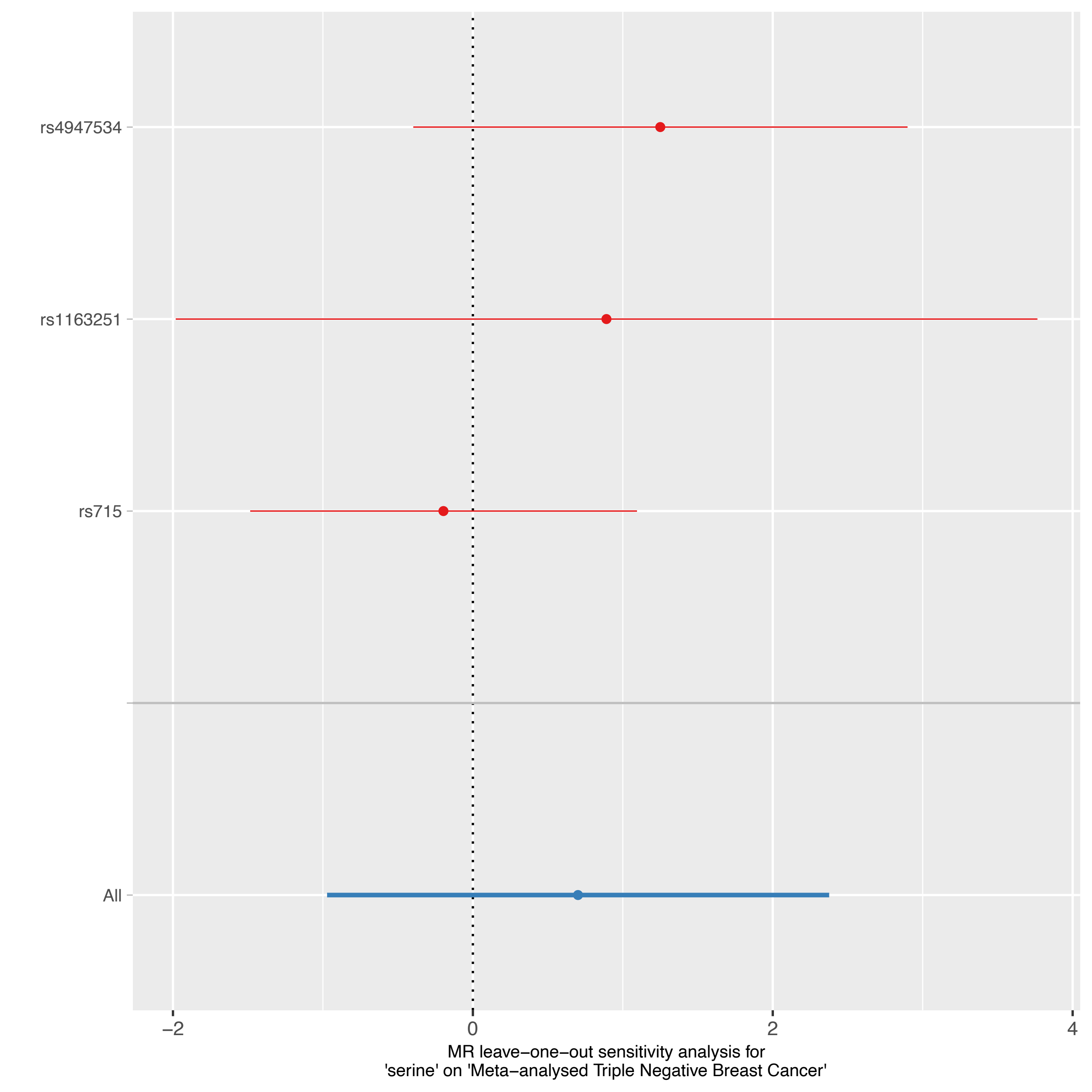
rs4947534
rs1163251
rs715
All
−2
0
2
4
MR leave−one−out sensitivity analysis for
'serine' on 'Meta−analysed Triple Negative Breast Cancer'

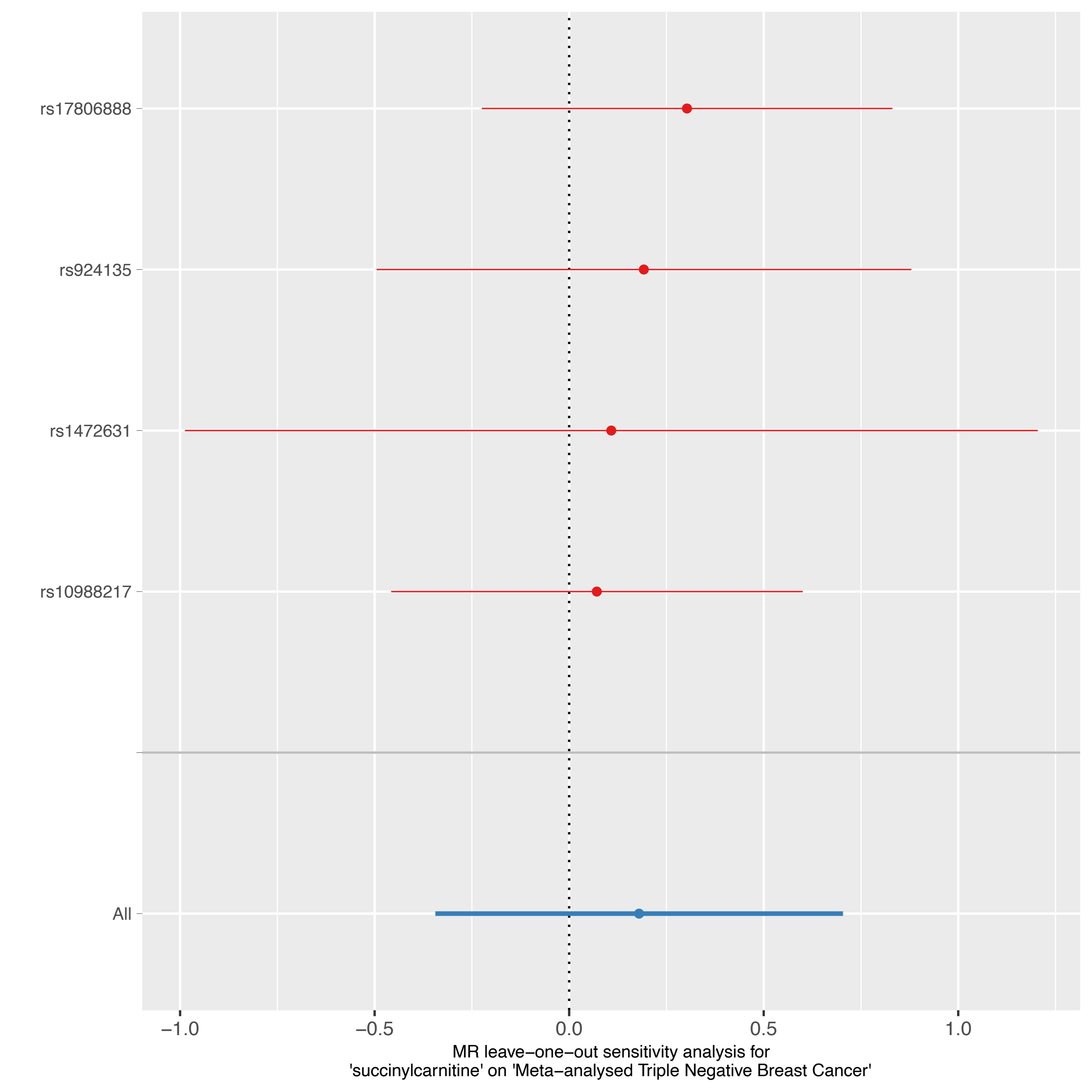
rs17806888
rs924135
rs1472631
rs10988217
All
−1.0
−0.5
0.0
0.5
1.0
MR leave−one−out sensitivity analysis for
'succinylcarnitine' on 'Meta−analysed Triple Negative Breast Cancer'

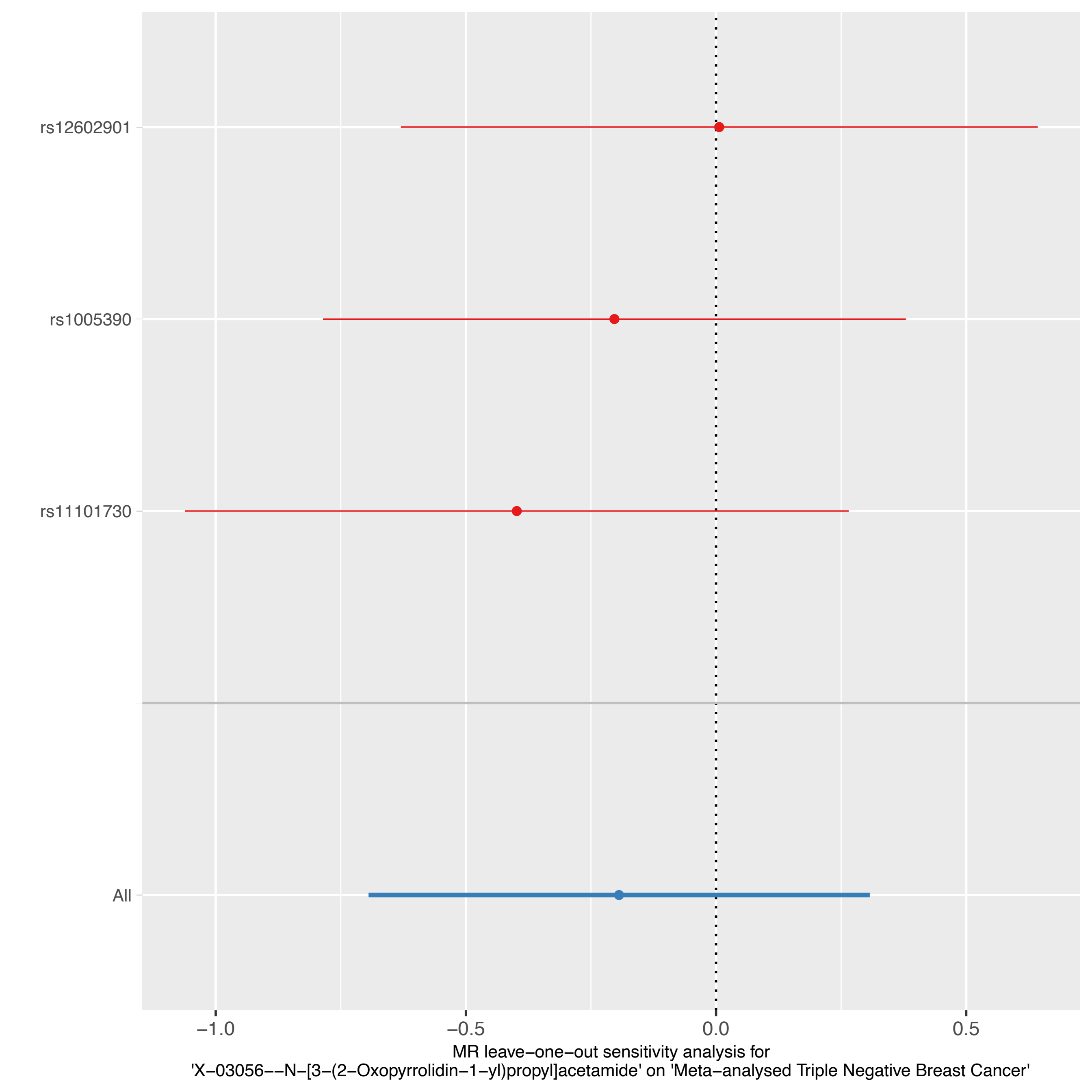
rs12602901
rs1005390
rs11101730
All
−1.0
−0.5
0.0
0.5
MR leave−one−out sensitivity analysis for
'X−03056−−N−[3−(2−Oxopyrrolidin−1−yl)propyl]acetamide' on 'Meta−analysed Triple Negative Breast Cancer'

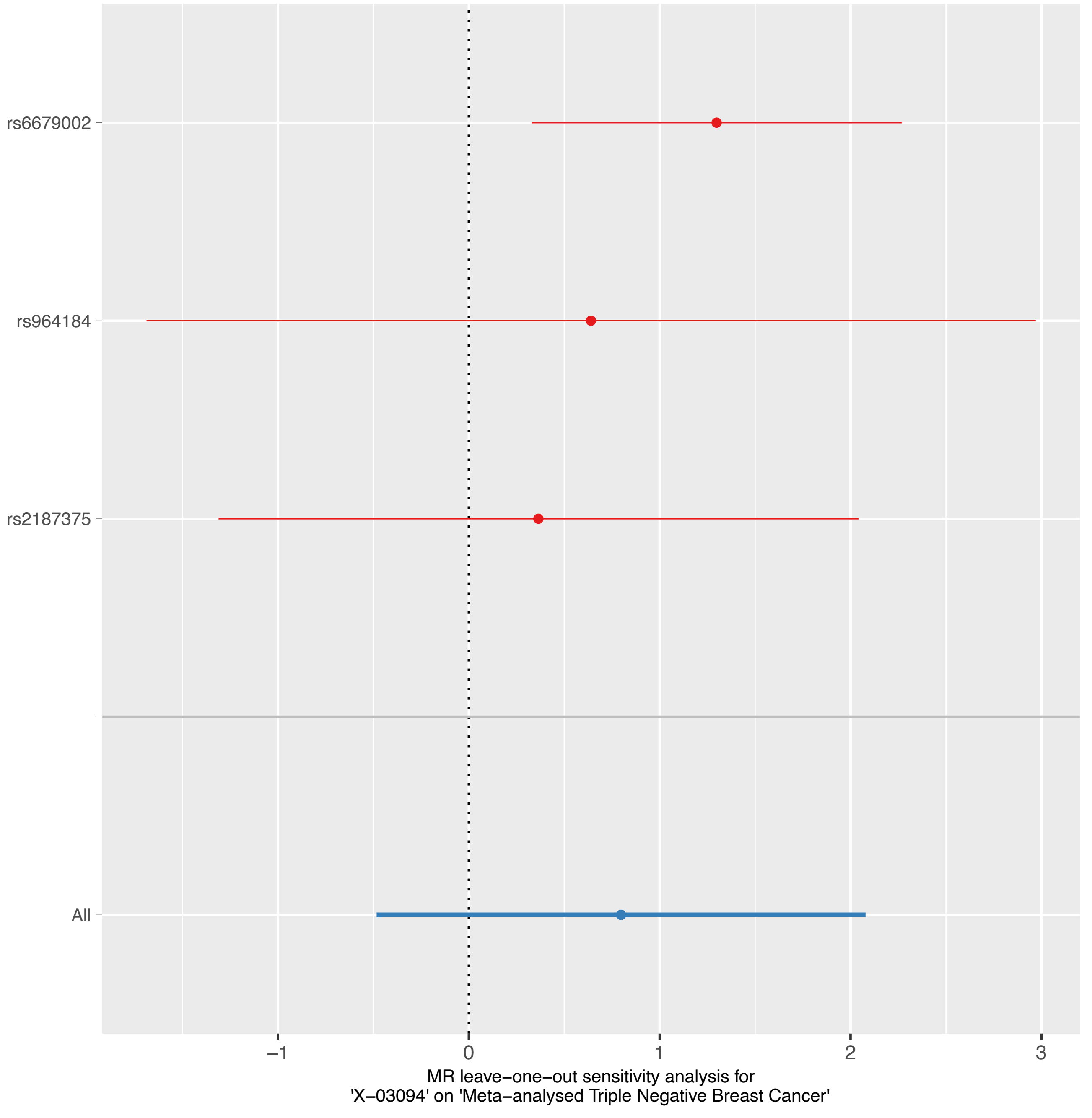

rs6679002
rs964184
rs2187375
All
−1
0
1
2
3
MR leave−one−out sensitivity analysis for
'X−03094' on 'Meta−analysed Triple Negative Breast Cancer'

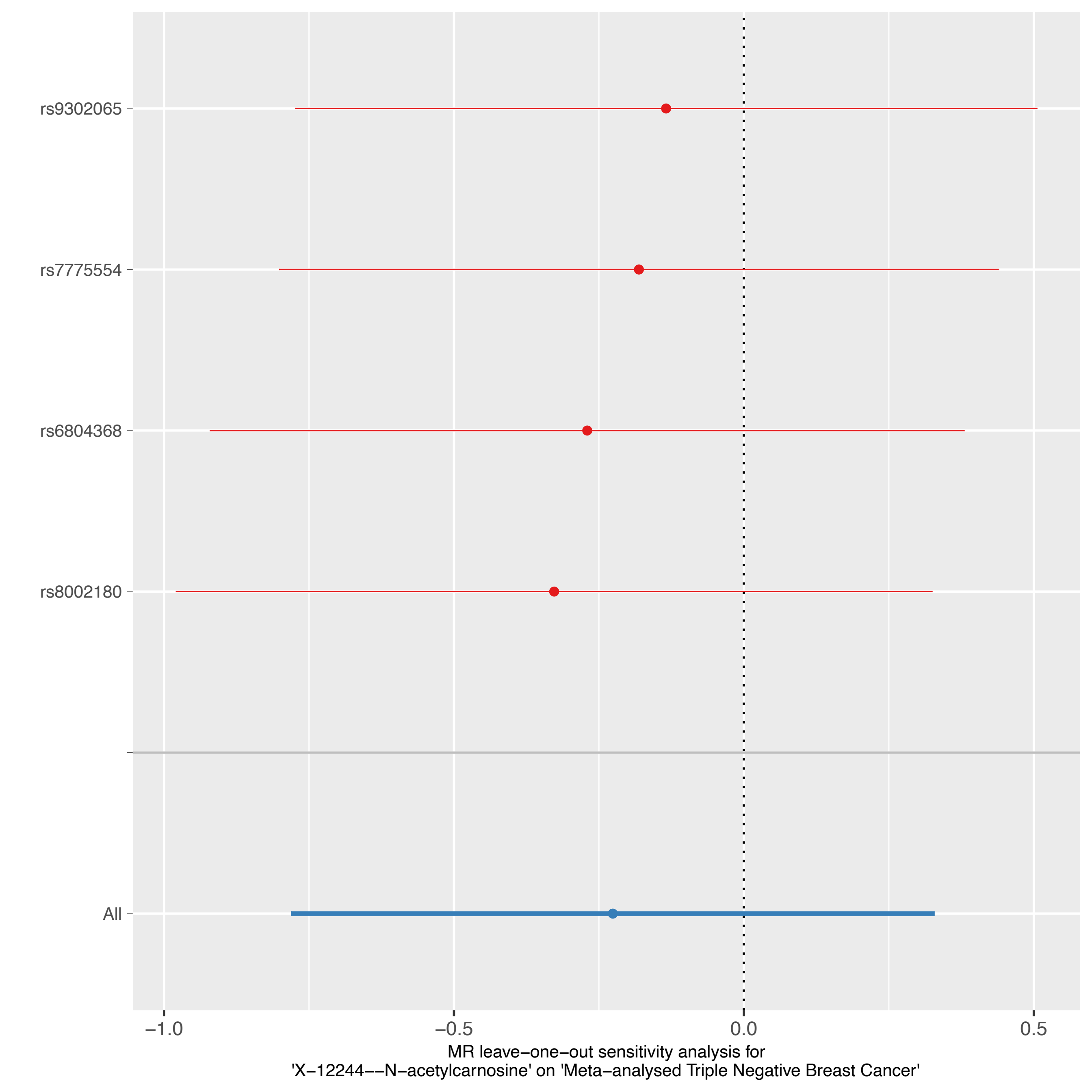

rs9302065
rs7775554
rs6804368
rs8002180
All
−1.0
−0.5
0.0
0.5
MR leave−one−out sensitivity analysis for
'X−12244−−N−acetylcarnosine' on 'Meta−analysed Triple Negative Breast Cancer'

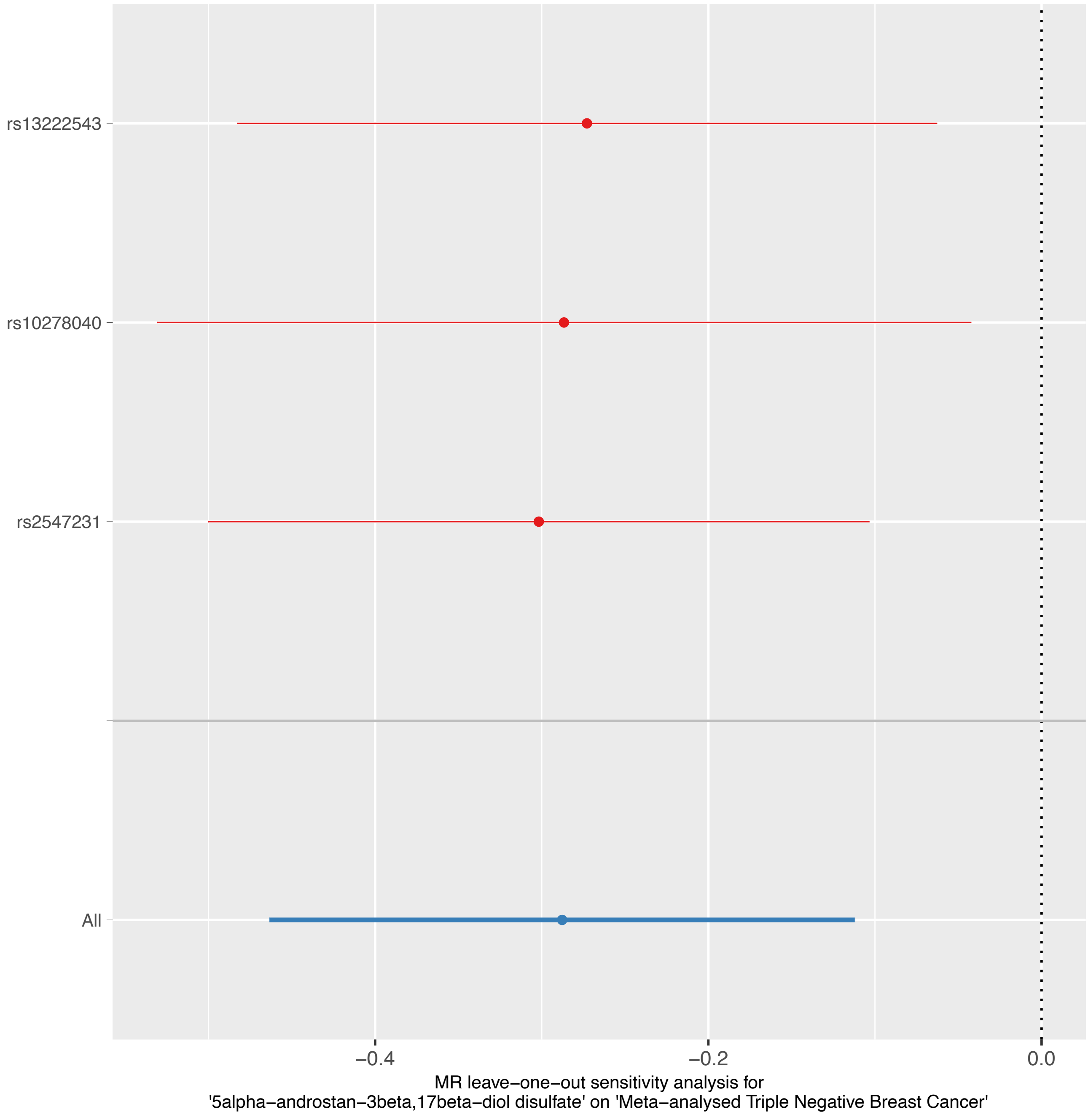
rs13222543
rs10278040
rs2547231
All
−0.4
−0.2
0.0
MR leave−one−out sensitivity analysis for
'5alpha−androstan−3beta,17beta−diol disulfate' on 'Meta−analysed Triple Negative Breast Cancer'

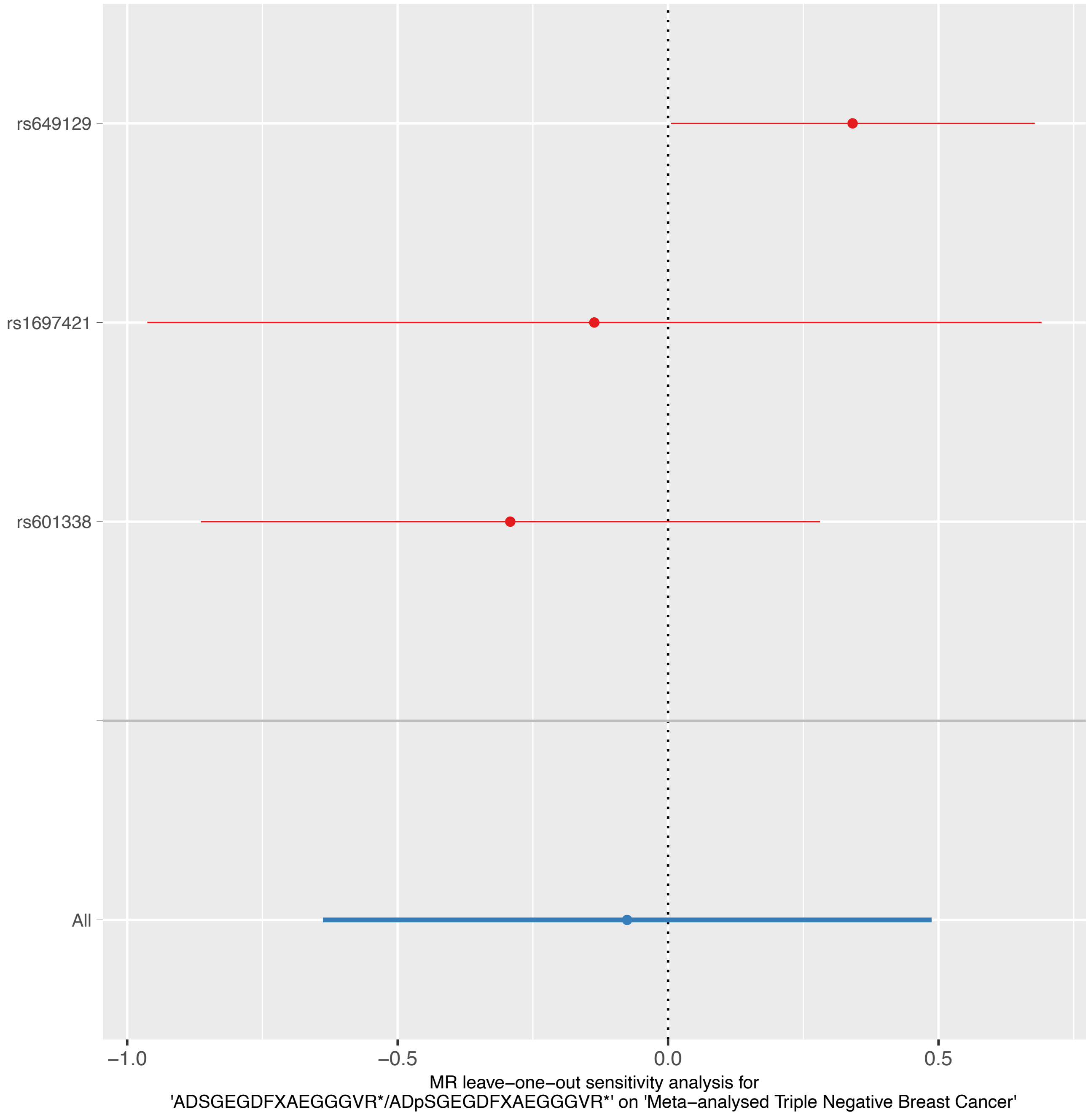
rs649129
rs1697421
rs601338
All
−1.0
−0.5
0.0
0.5
MR leave−one−out sensitivity analysis for
'ADSGEGDFXAEGGGVR*/ADpSGEGDFXAEGGGVR*' on 'Meta−analysed Triple Negative Breast Cancer'

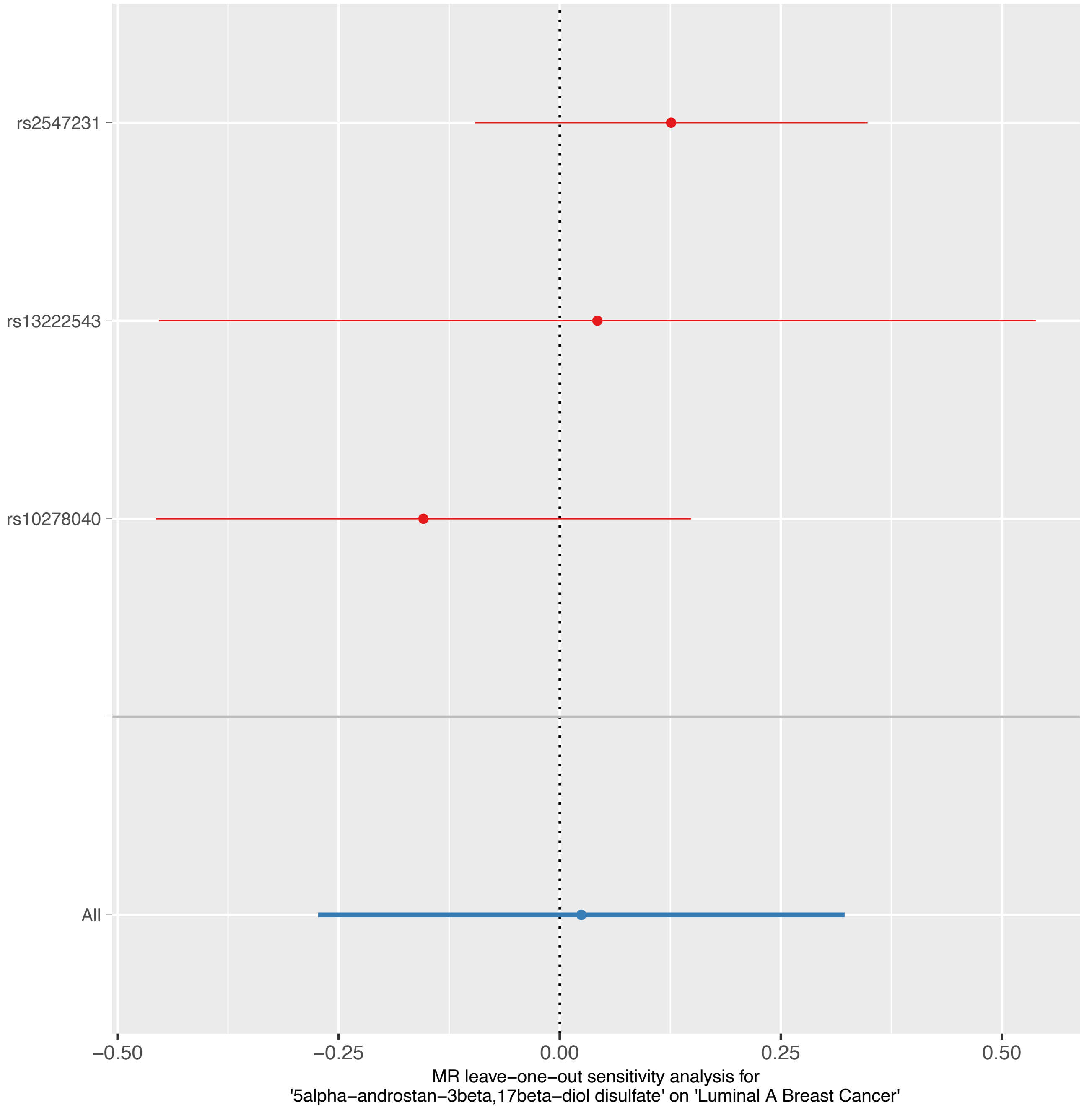

rs2547231
rs13222543
rs10278040
All
−0.50
−0.25
0.00
0.25
0.50
MR leave−one−out sensitivity analysis for
'5alpha−androstan−3beta,17beta−diol disulfate' on 'Luminal A Breast Cancer'

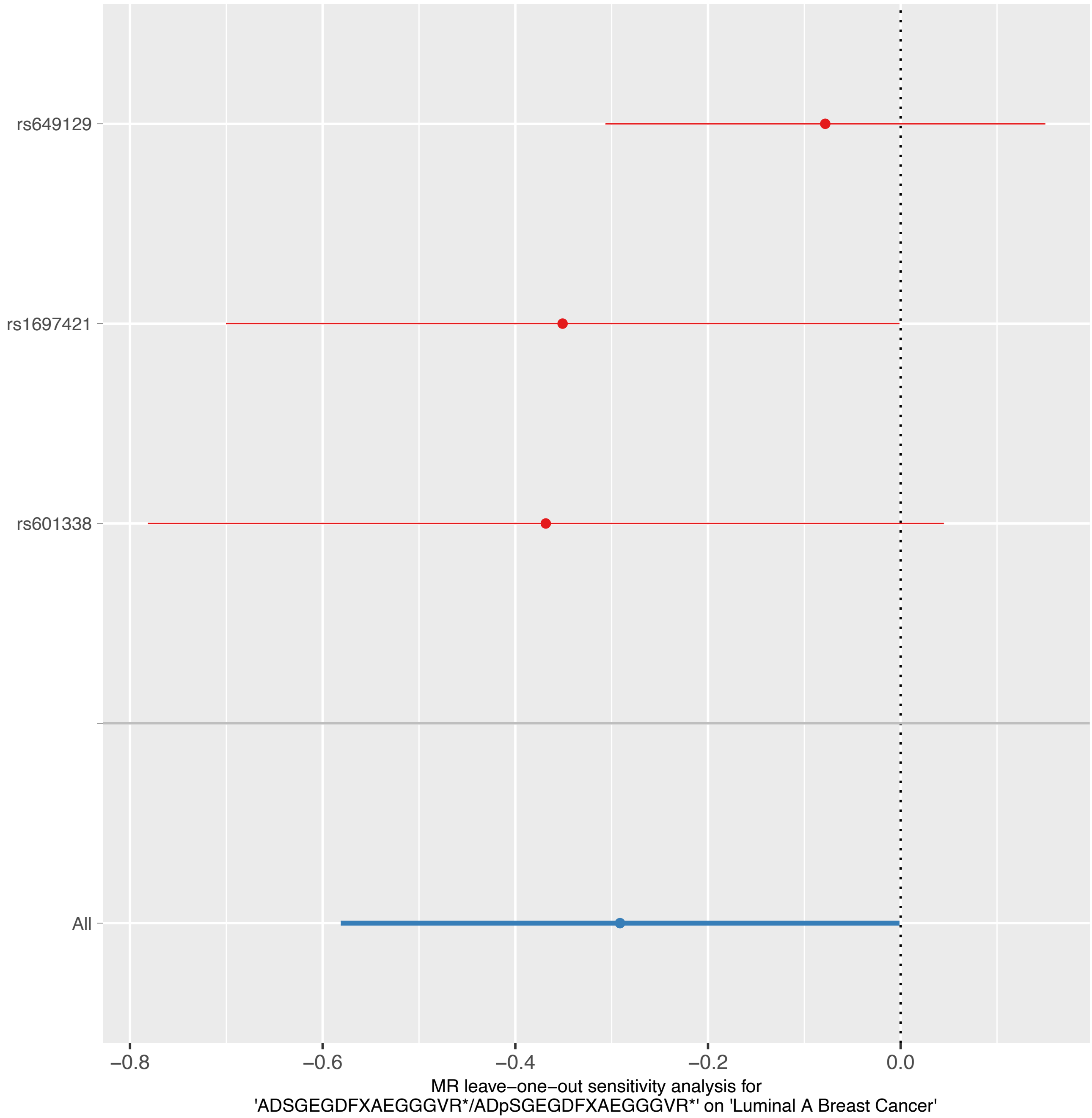
rs649129
rs1697421
rs601338
All
−0.8
−0.6
−0.4
−0.2
0.0
MR leave−one−out sensitivity analysis for
'ADSGEGDFXAEGGGVR*/ADpSGEGDFXAEGGGVR*' on 'Luminal A Breast Cancer'

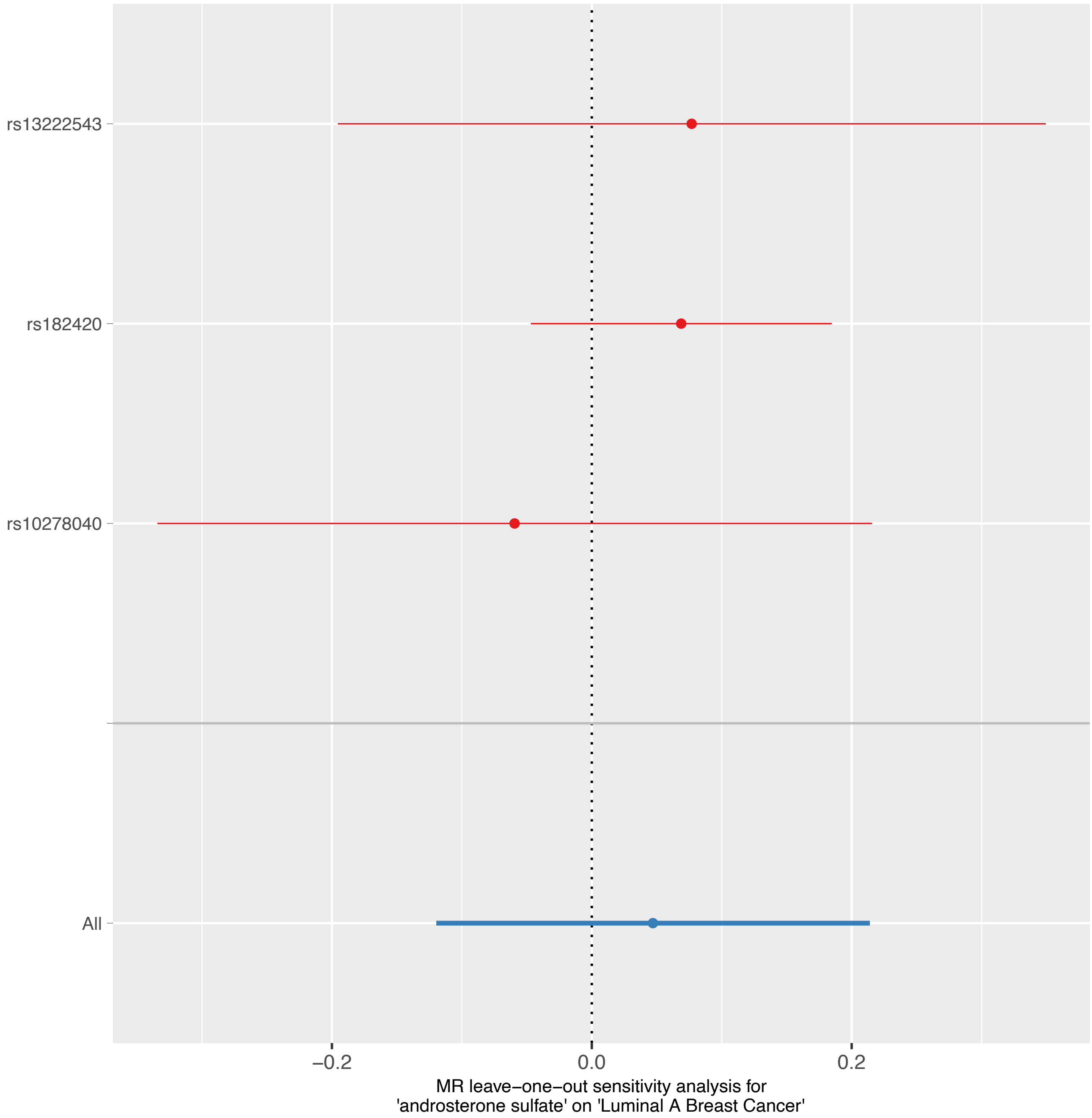
rs13222543
rs182420
rs10278040
All
−0.2
0.0
0.2
MR leave−one−out sensitivity analysis for
'androsterone sulfate' on 'Luminal A Breast Cancer'

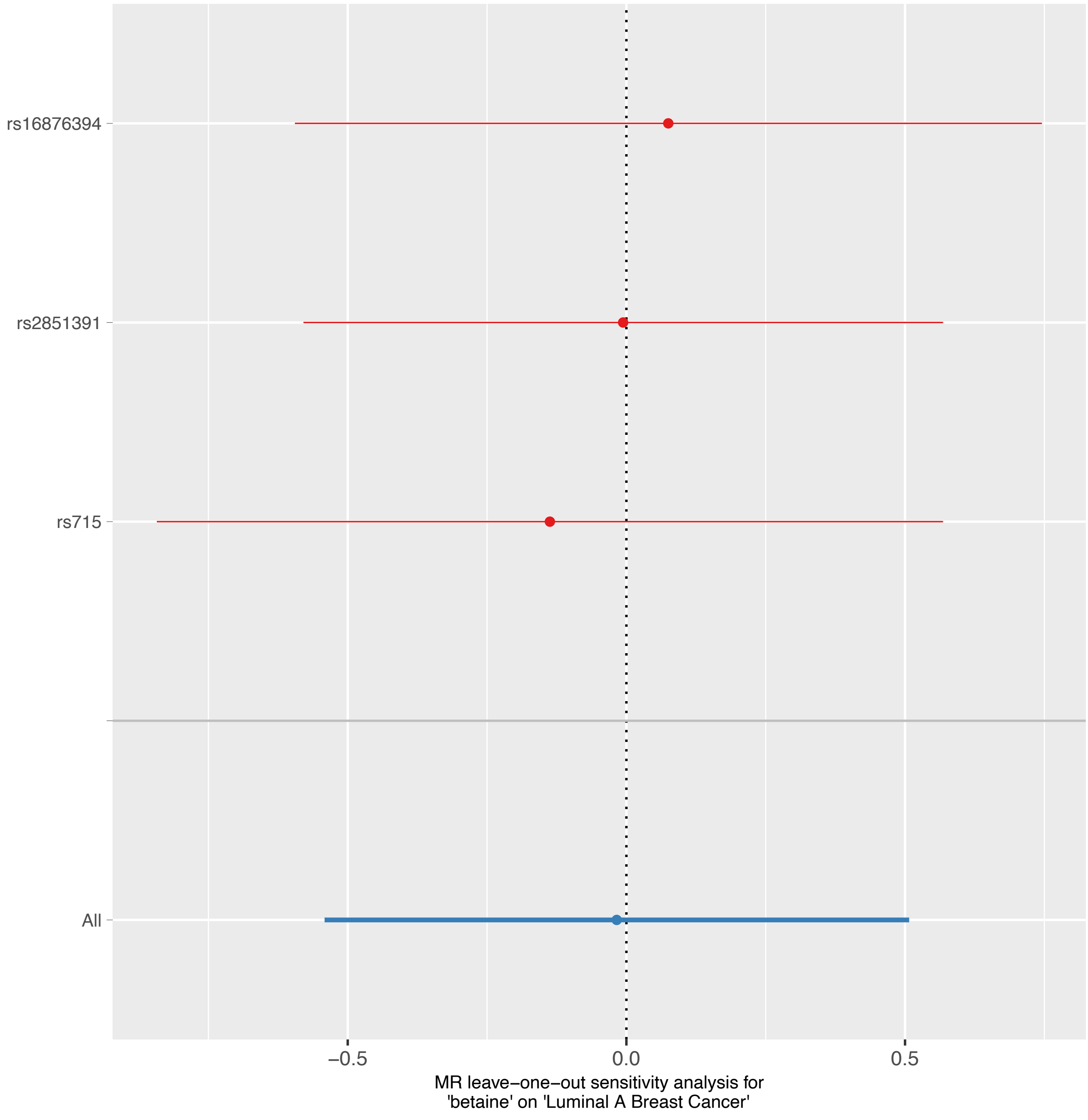
rs16876394
rs2851391
rs715
All
−0.5
0.0
0.5
MR leave−one−out sensitivity analysis for
'betaine' on 'Luminal A Breast Cancer'

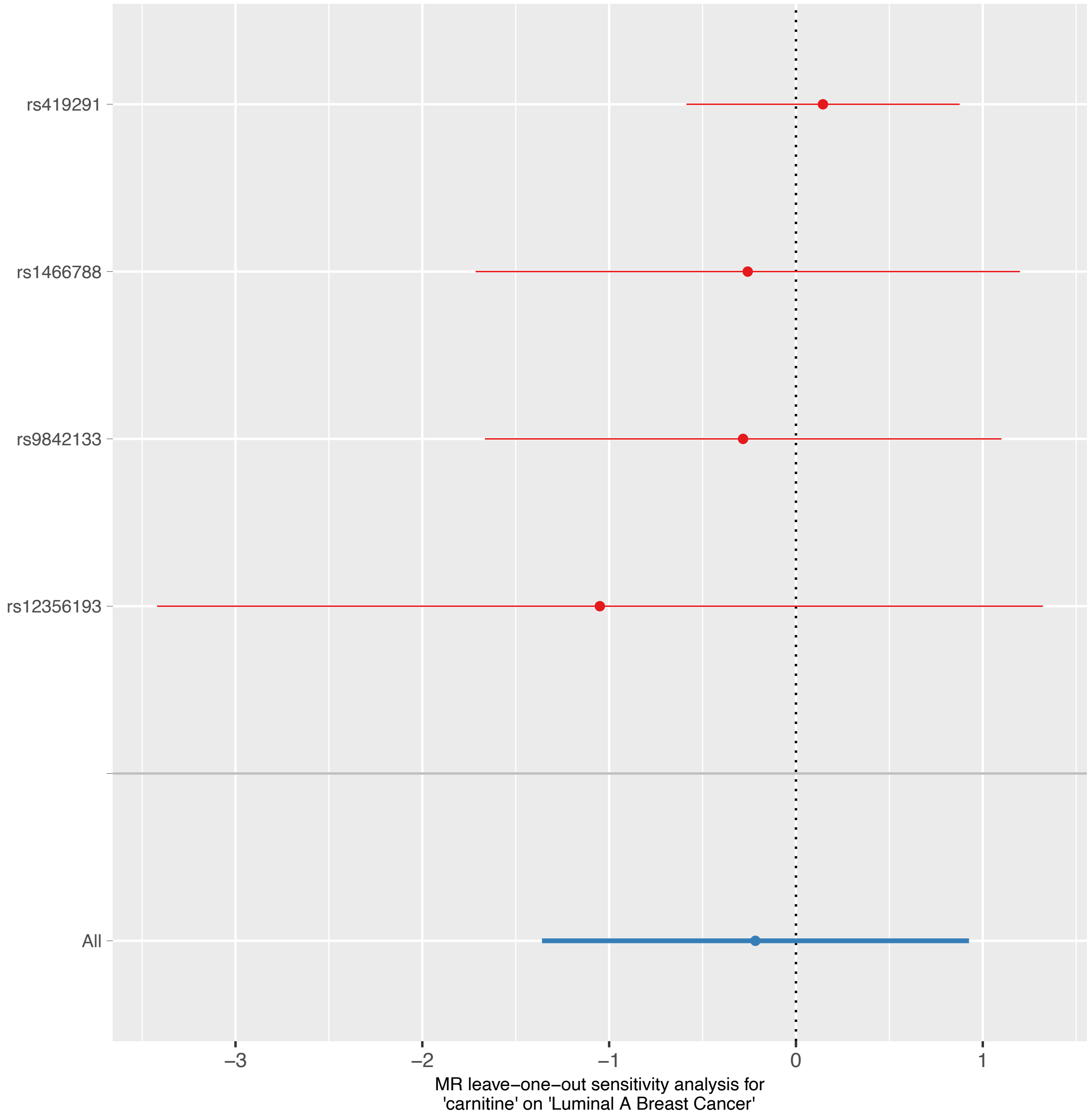
rs419291
rs1466788
rs9842133
rs12356193
All
−3
−2
−1
0
1
MR leave−one−out sensitivity analysis for
'carnitine' on 'Luminal A Breast Cancer'

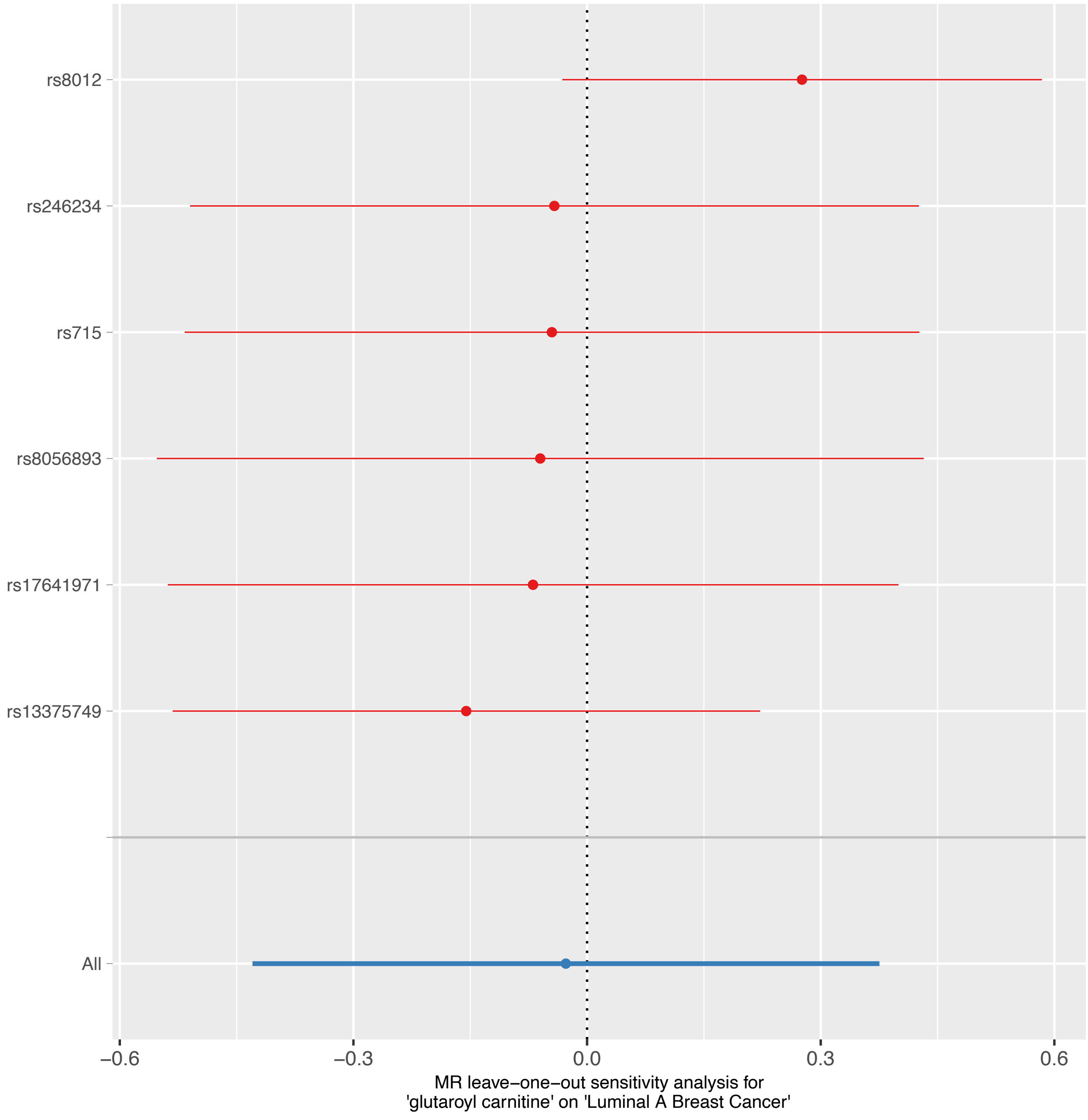

rs8012
rs246234
rs715
rs8056893
rs17641971
rs13375749
All
−0.6
−0.3
0.0
0.3
0.6
MR leave−one−out sensitivity analysis for
'glutaroyl carnitine' on 'Luminal A Breast Cancer'

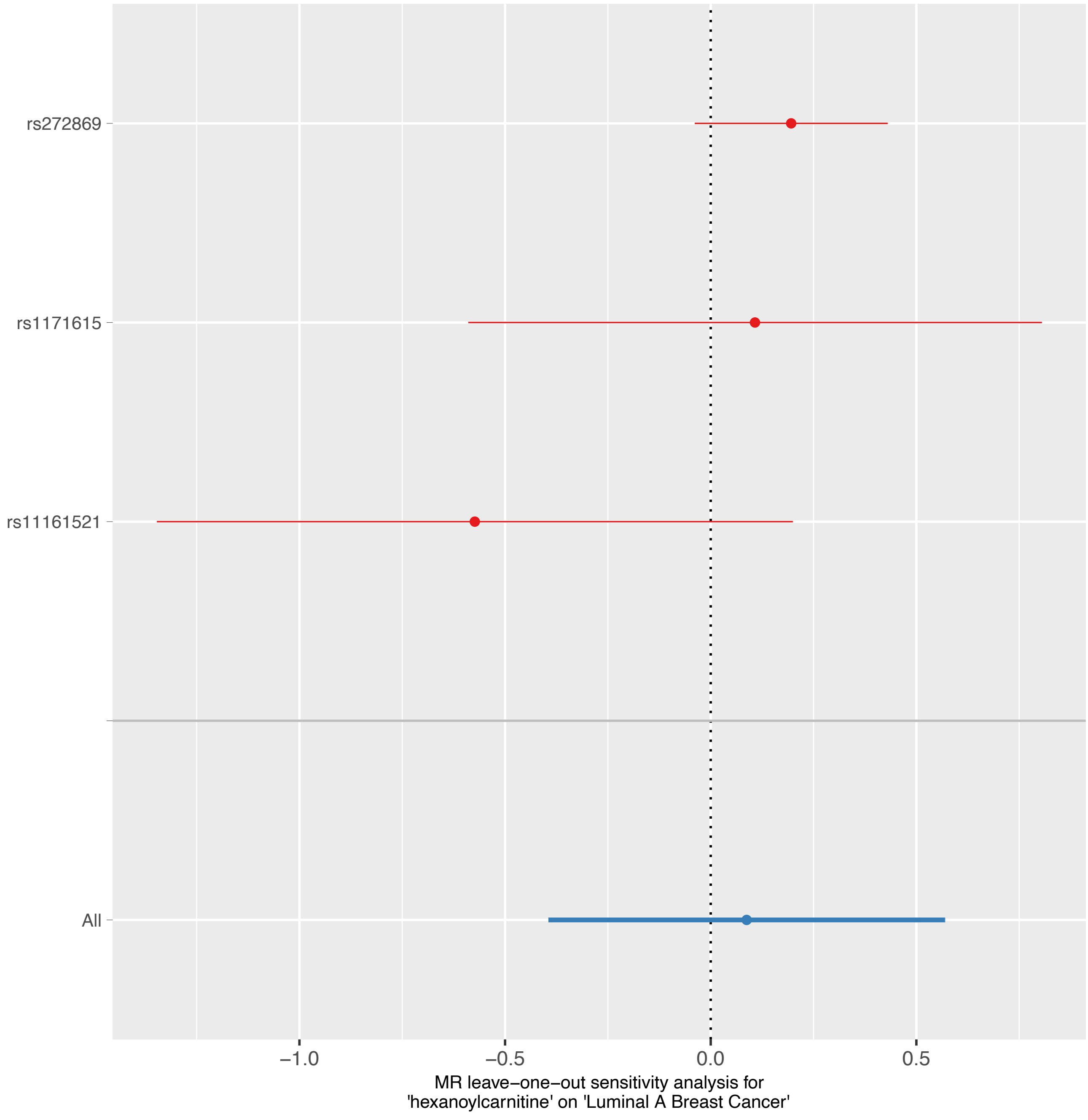

rs272869
rs1171615
rs11161521
All
−1.0
−0.5
0.0
0.5
MR leave−one−out sensitivity analysis for
'hexanoylcarnitine' on 'Luminal A Breast Cancer'

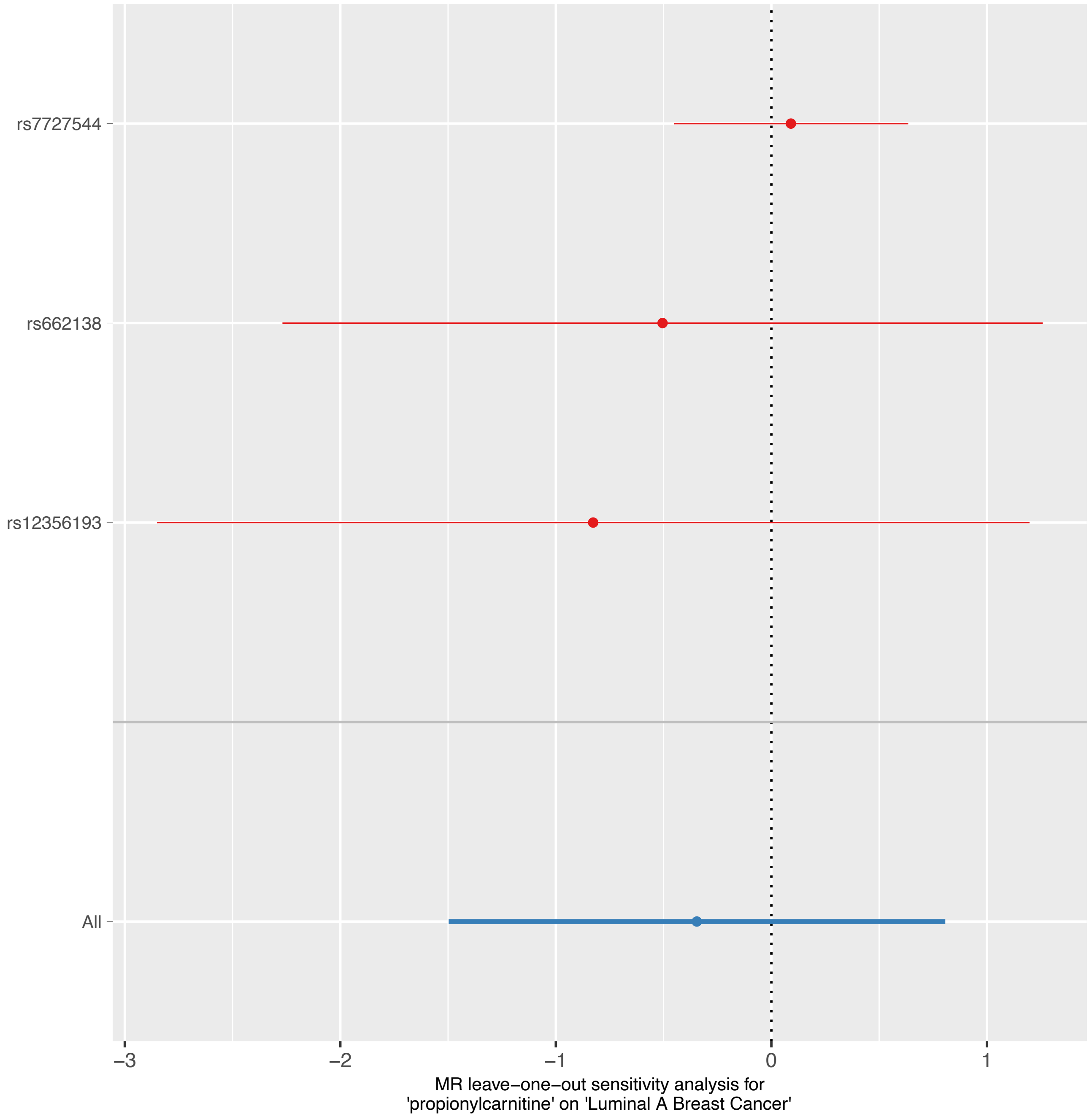
rs7727544
rs662138
rs12356193
All
−3
−2
−1
0
1
MR leave−one−out sensitivity analysis for
'propionylcarnitine' on 'Luminal A Breast Cancer'

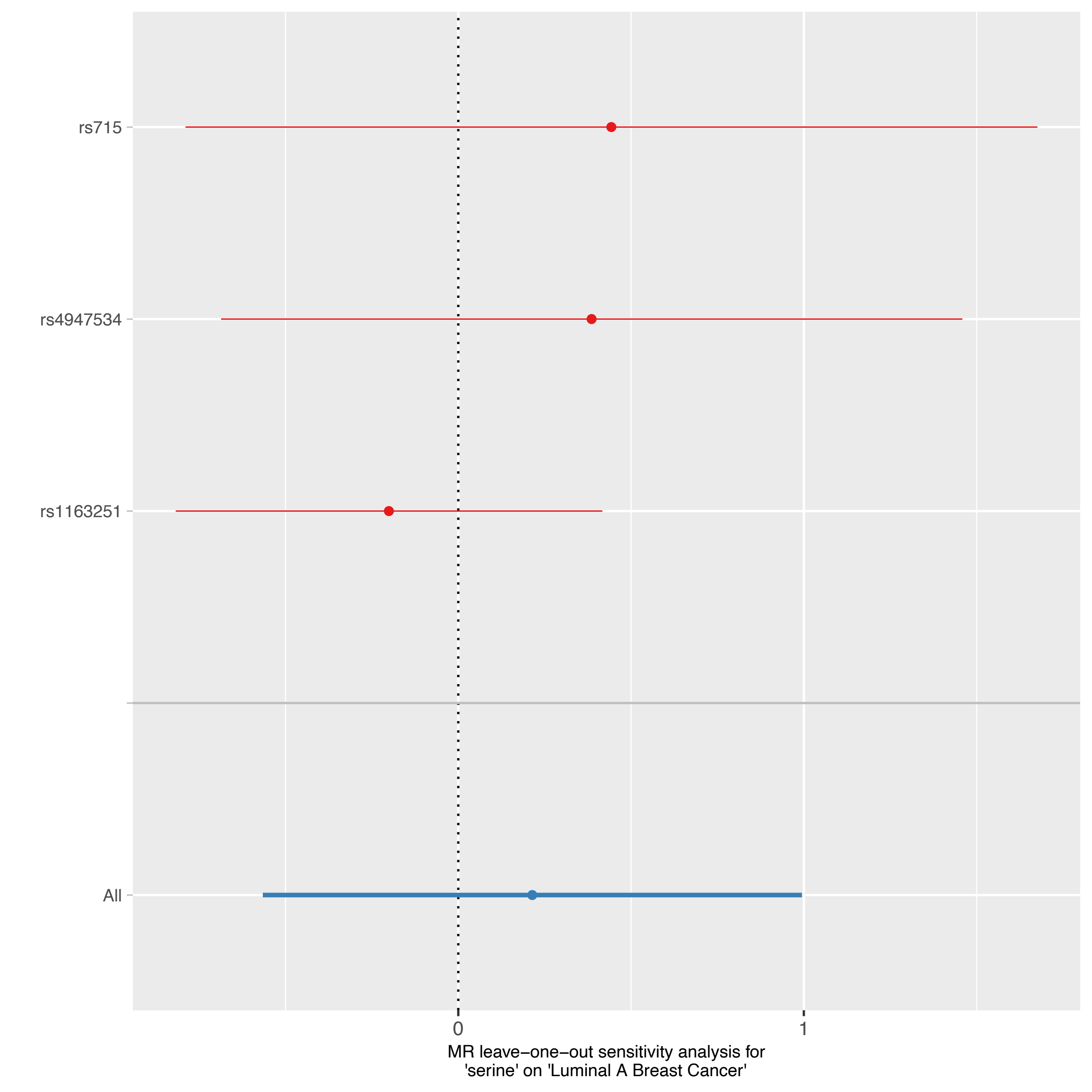

rs715
rs4947534
rs1163251
All
0
1
MR leave−one−out sensitivity analysis for
'serine' on 'Luminal A Breast Cancer'

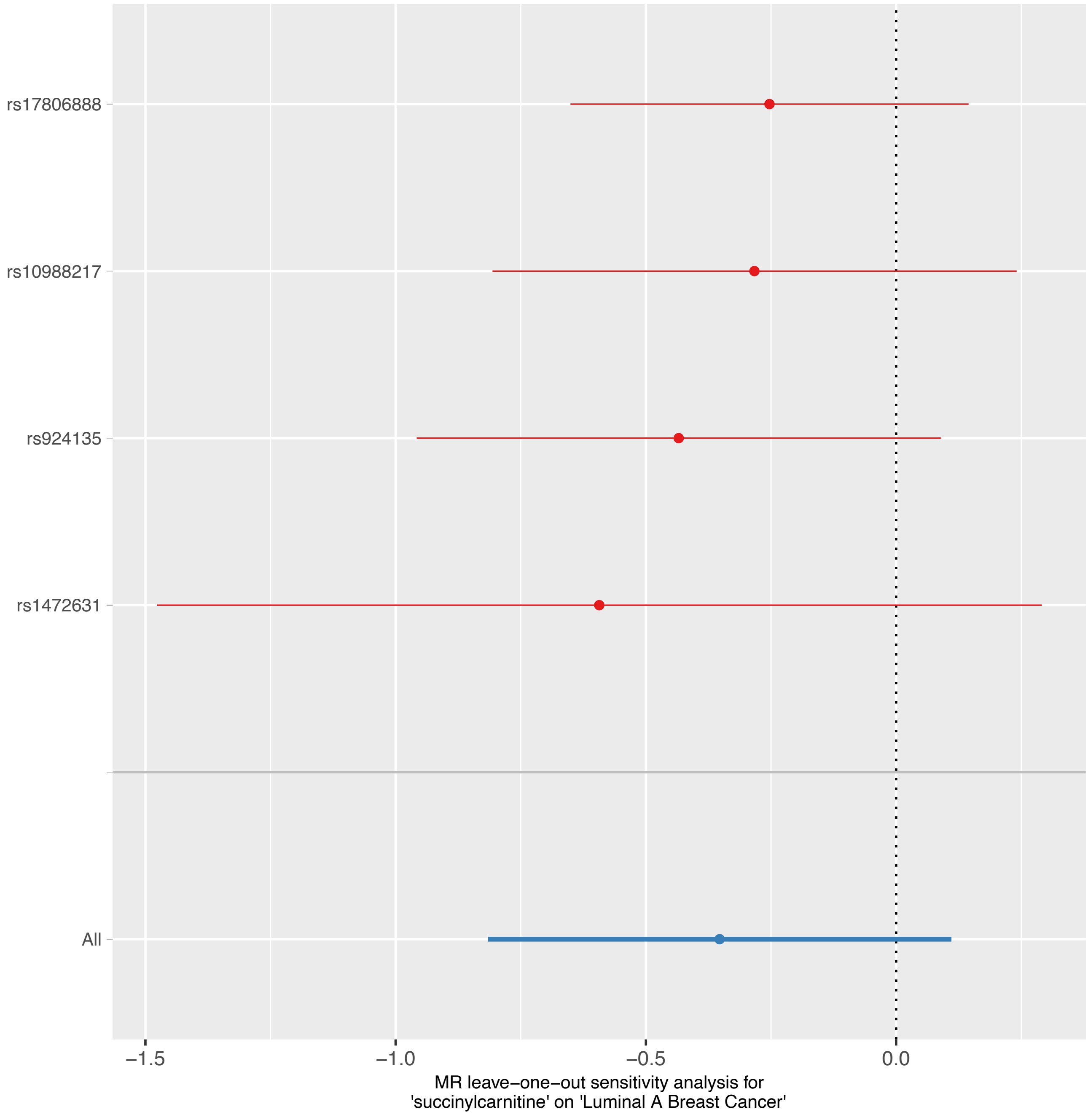
rs17806888
rs10988217
rs924135
rs1472631
All
−1.5
−1.0
−0.5
0.0
MR leave−one−out sensitivity analysis for
'succinylcarnitine' on 'Luminal A Breast Cancer'

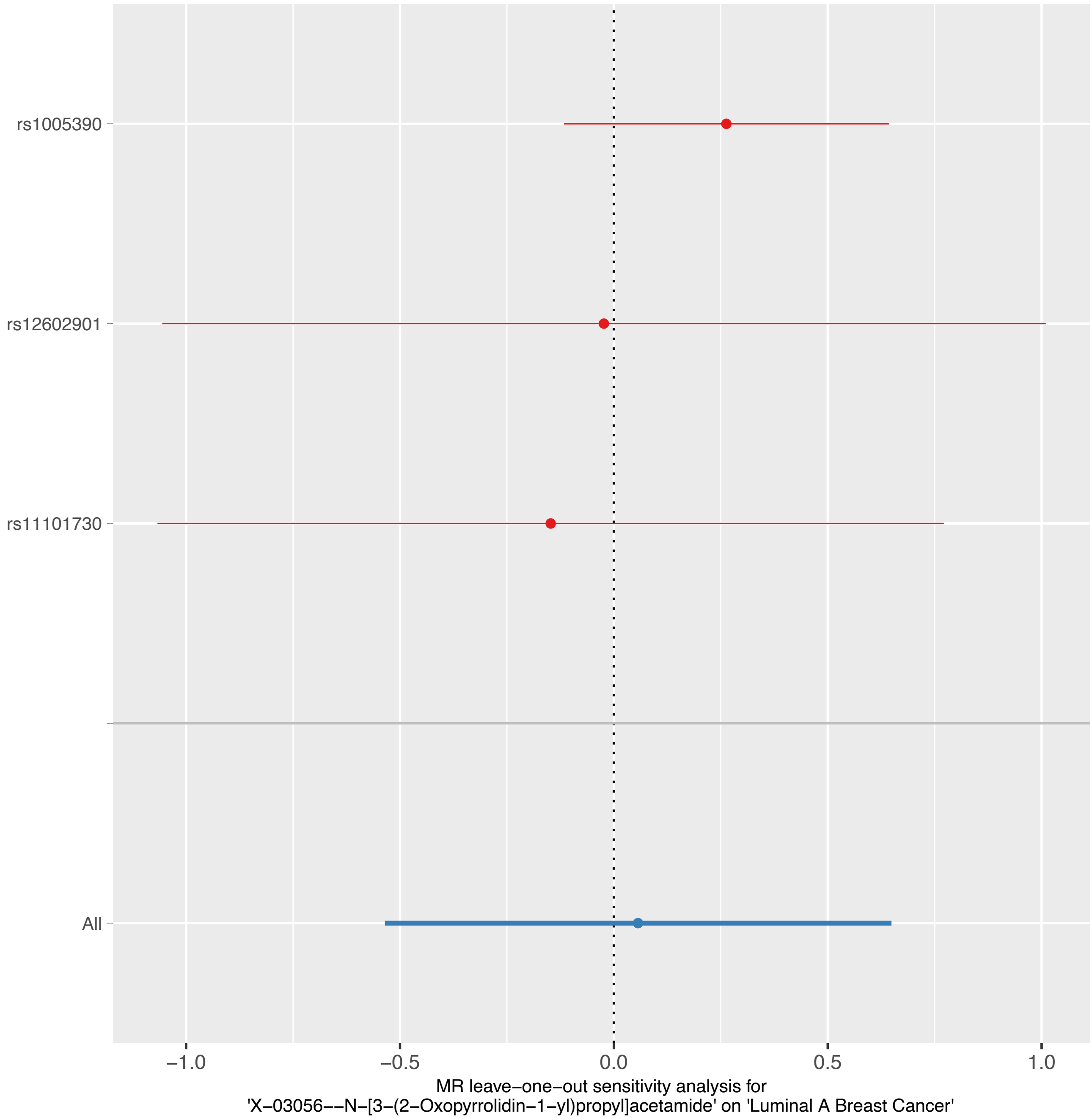
rs1005390
rs12602901
rs11101730
All
−1.0
−0.5
0.0
0.5
1.0
MR leave−one−out sensitivity analysis for
'X−03056−−N−[3−(2−Oxopyrrolidin−1−yl)propyl]acetamide' on 'Luminal A Breast Cancer'

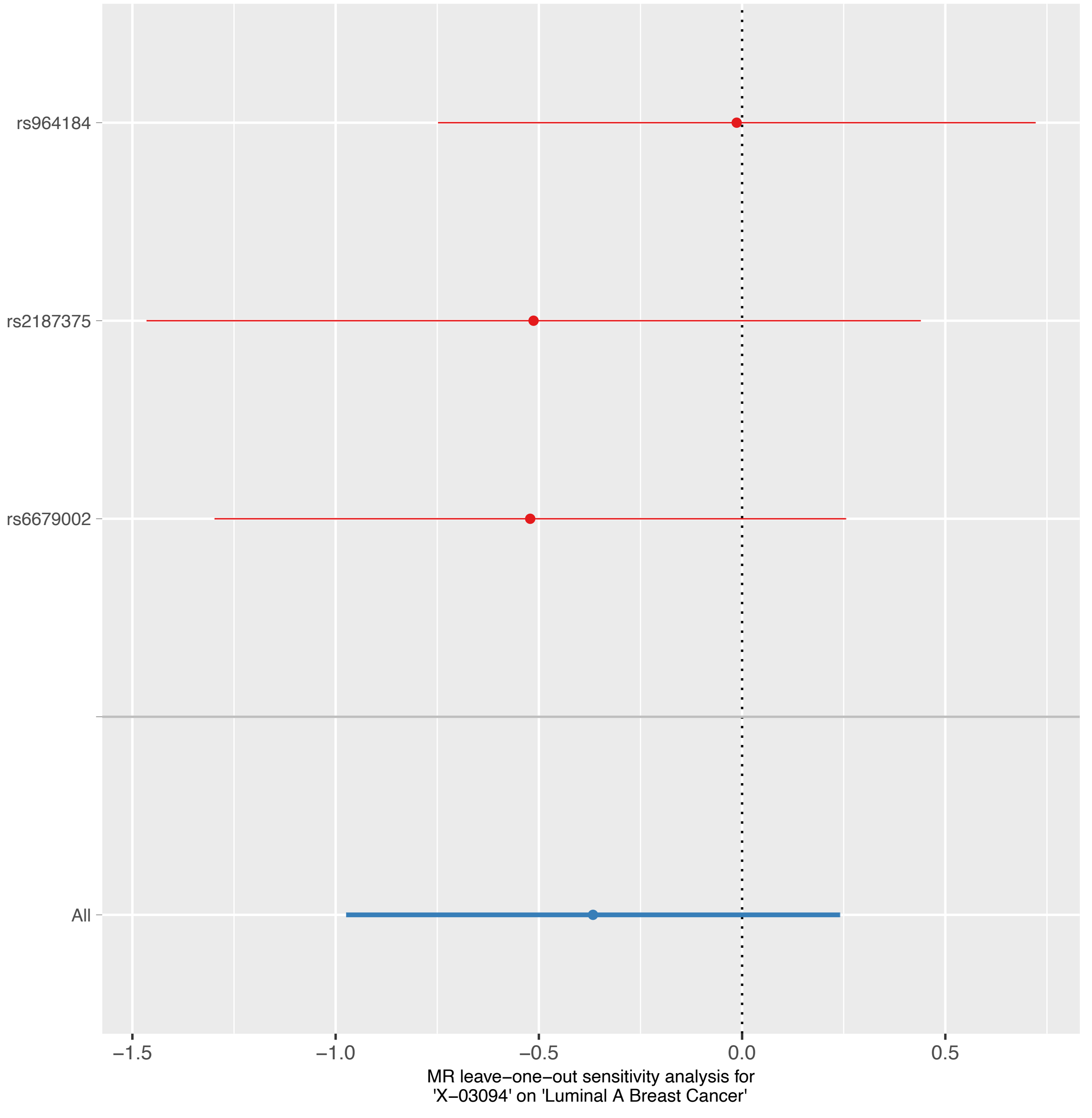
rs964184
rs2187375
rs6679002
All
−1.5
−1.0
−0.5
0.0
0.5
MR leave−one−out sensitivity analysis for
'X−03094' on 'Luminal A Breast Cancer'

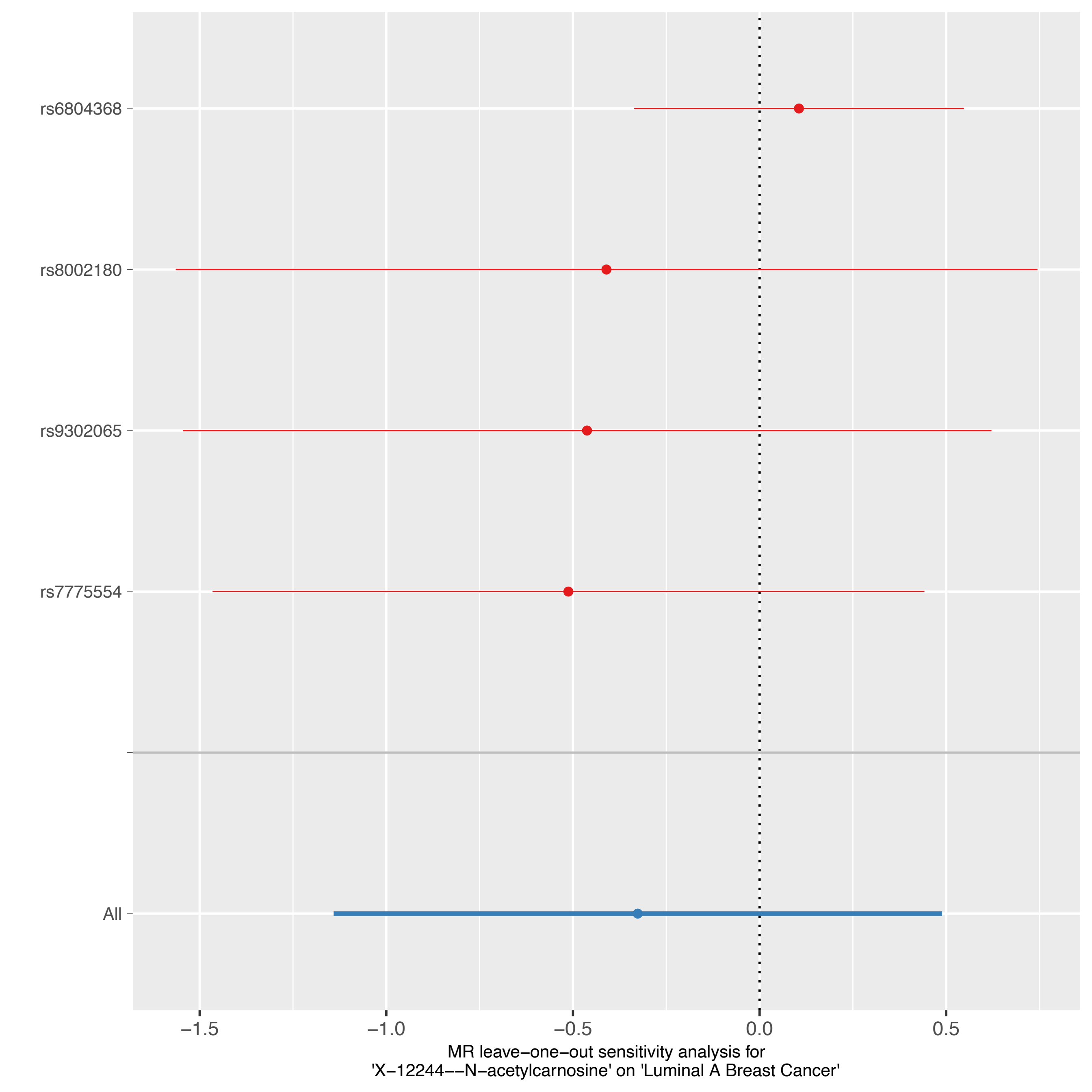

rs6804368
rs8002180
rs9302065
rs7775554
All
−1.5
−1.0
−0.5
0.0
0.5
MR leave−one−out sensitivity analysis for
'X−12244−−N−acetylcarnosine' on 'Luminal A Breast Cancer'

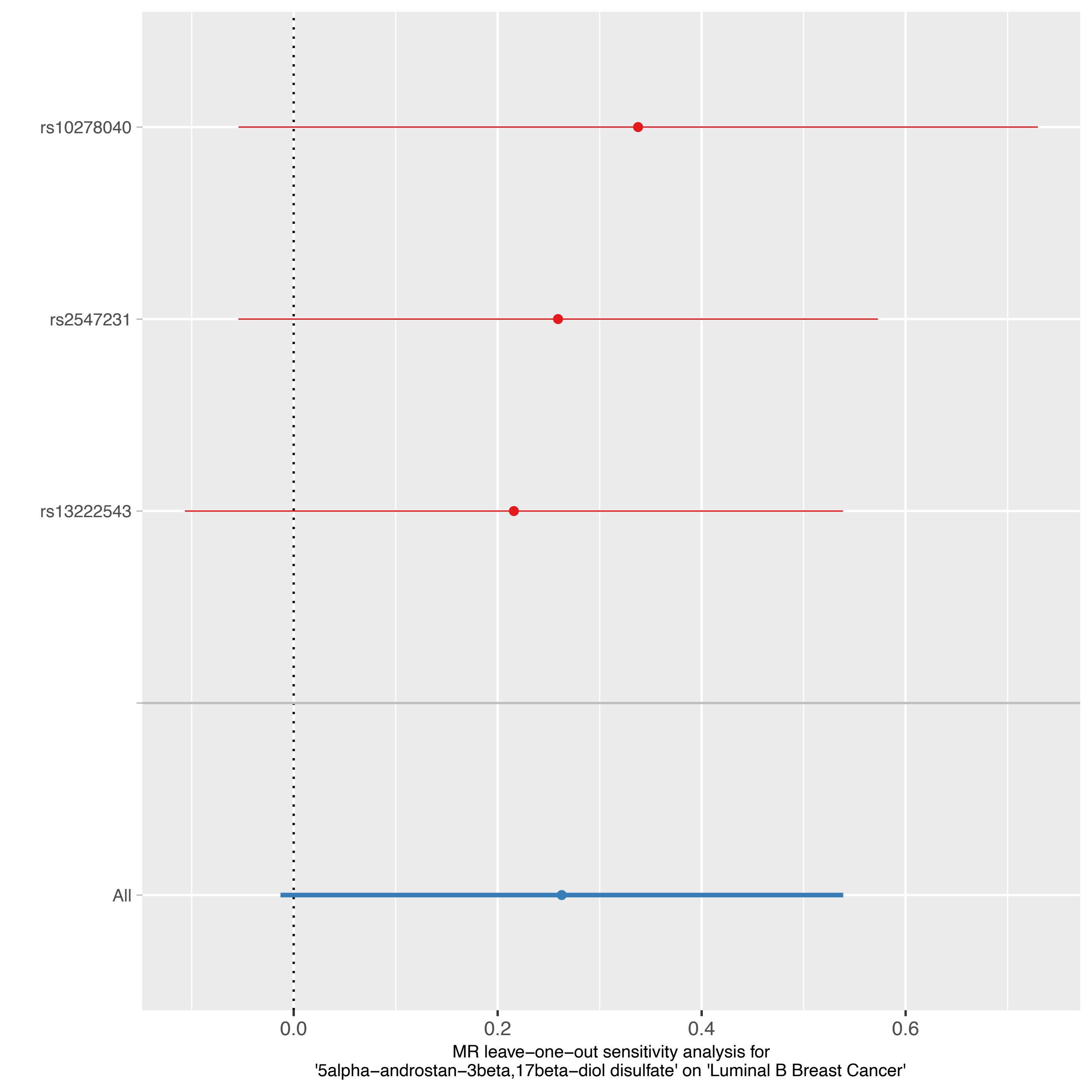
rs10278040
rs2547231
rs13222543
All
0.0
0.2
0.4
0.6
MR leave−one−out sensitivity analysis for
'5alpha−androstan−3beta,17beta−diol disulfate' on 'Luminal B Breast Cancer'

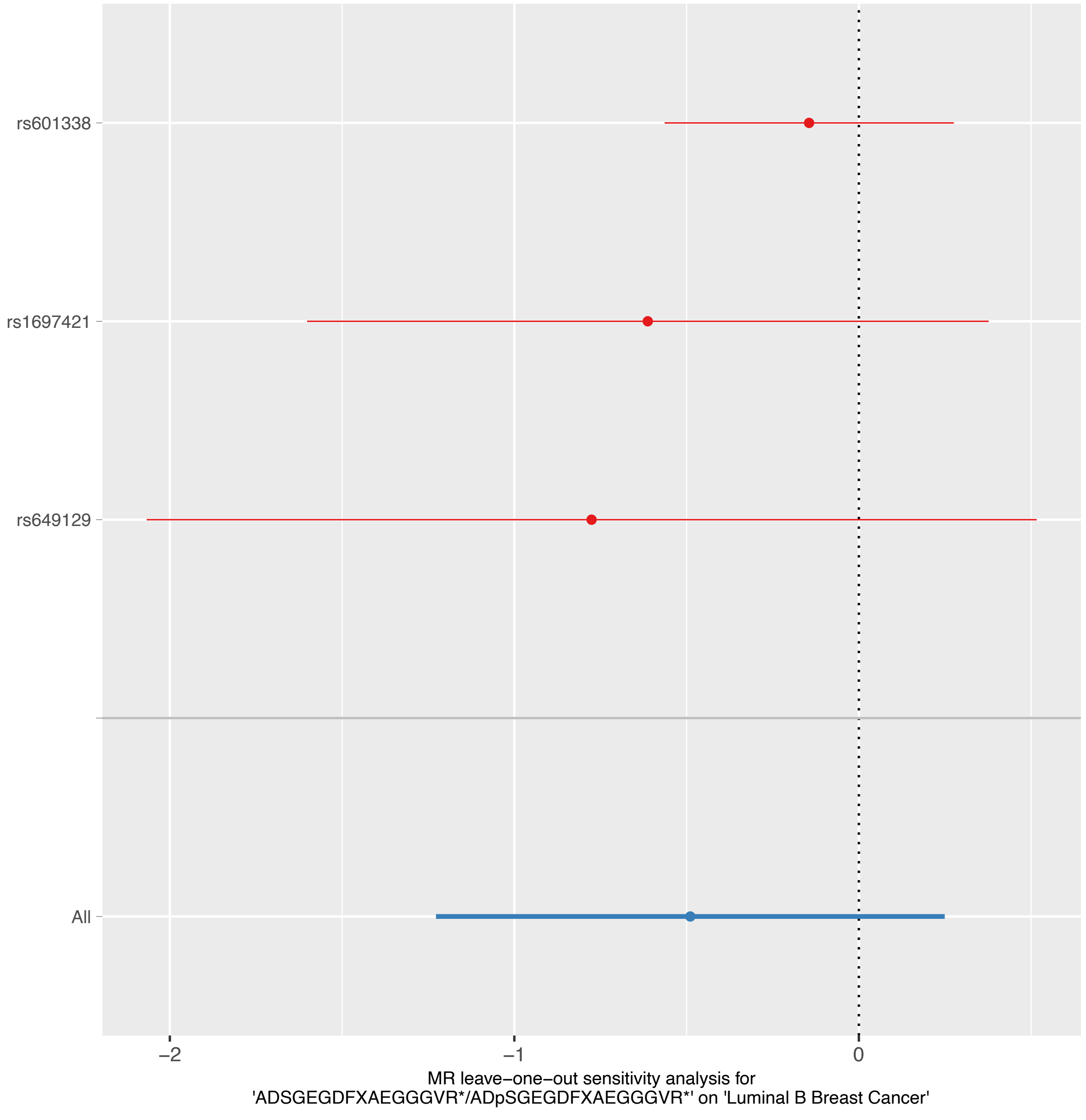
rs601338
rs1697421
rs649129
All
−2
−1
0
MR leave−one−out sensitivity analysis for
'ADSGEGDFXAEGGGVR*/ADpSGEGDFXAEGGGVR*' on 'Luminal B Breast Cancer'

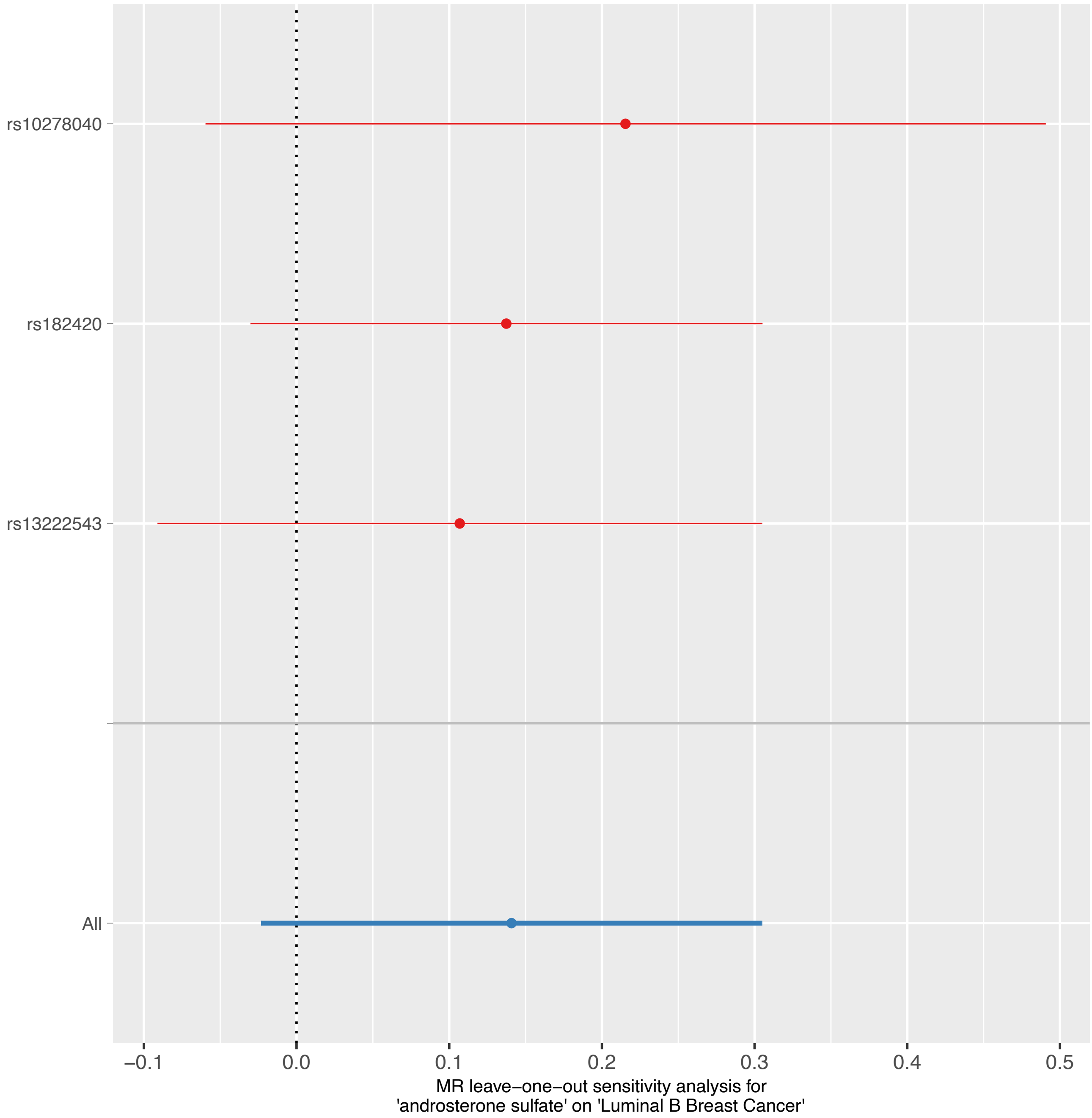

rs10278040
rs182420
rs13222543
All
−0.1
0.0
0.1
0.2
0.3
0.4
0.5
MR leave−one−out sensitivity analysis for
'androsterone sulfate' on 'Luminal B Breast Cancer'

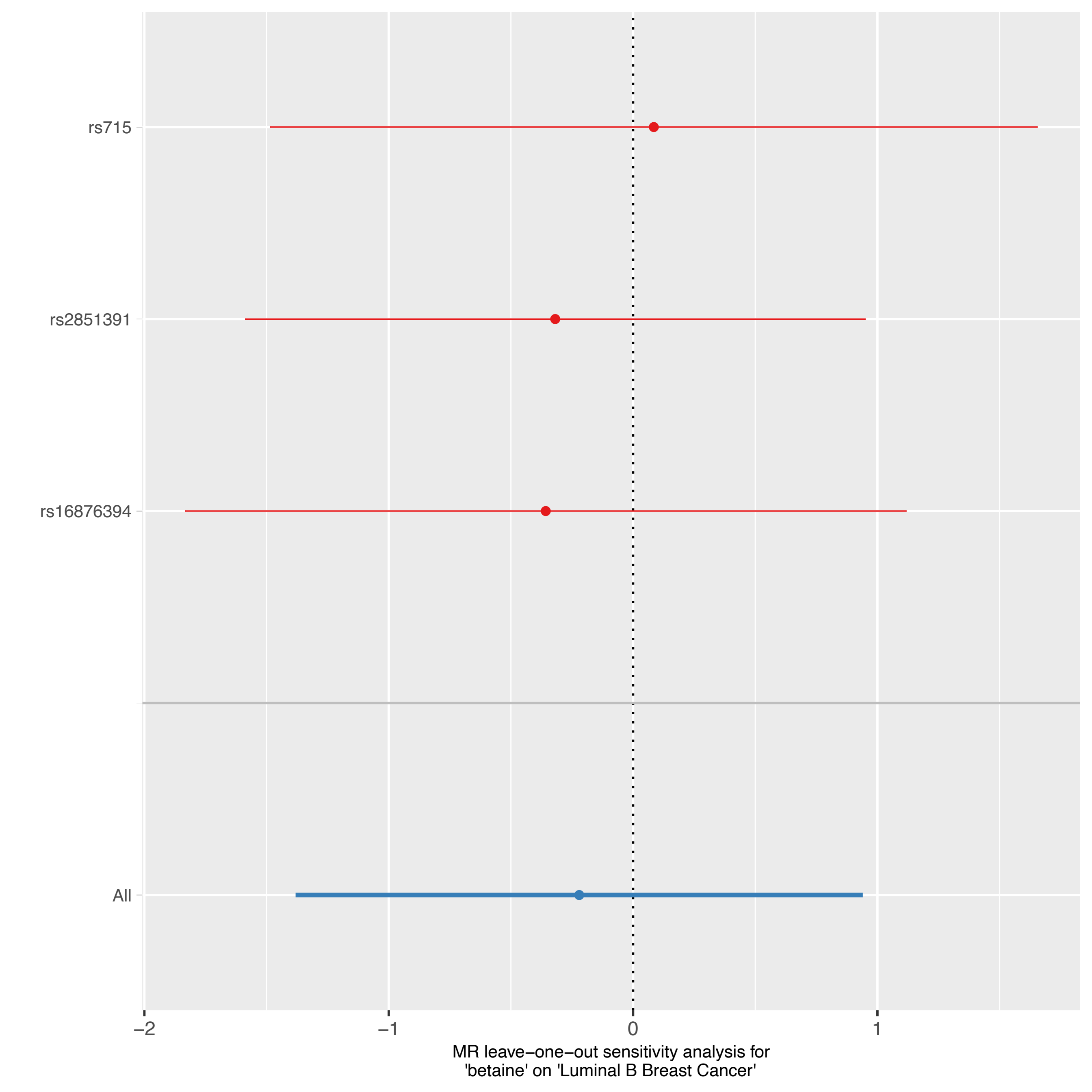
rs715
rs2851391
rs16876394
All
−2
−1
0
1
MR leave−one−out sensitivity analysis for
'betaine' on 'Luminal B Breast Cancer'

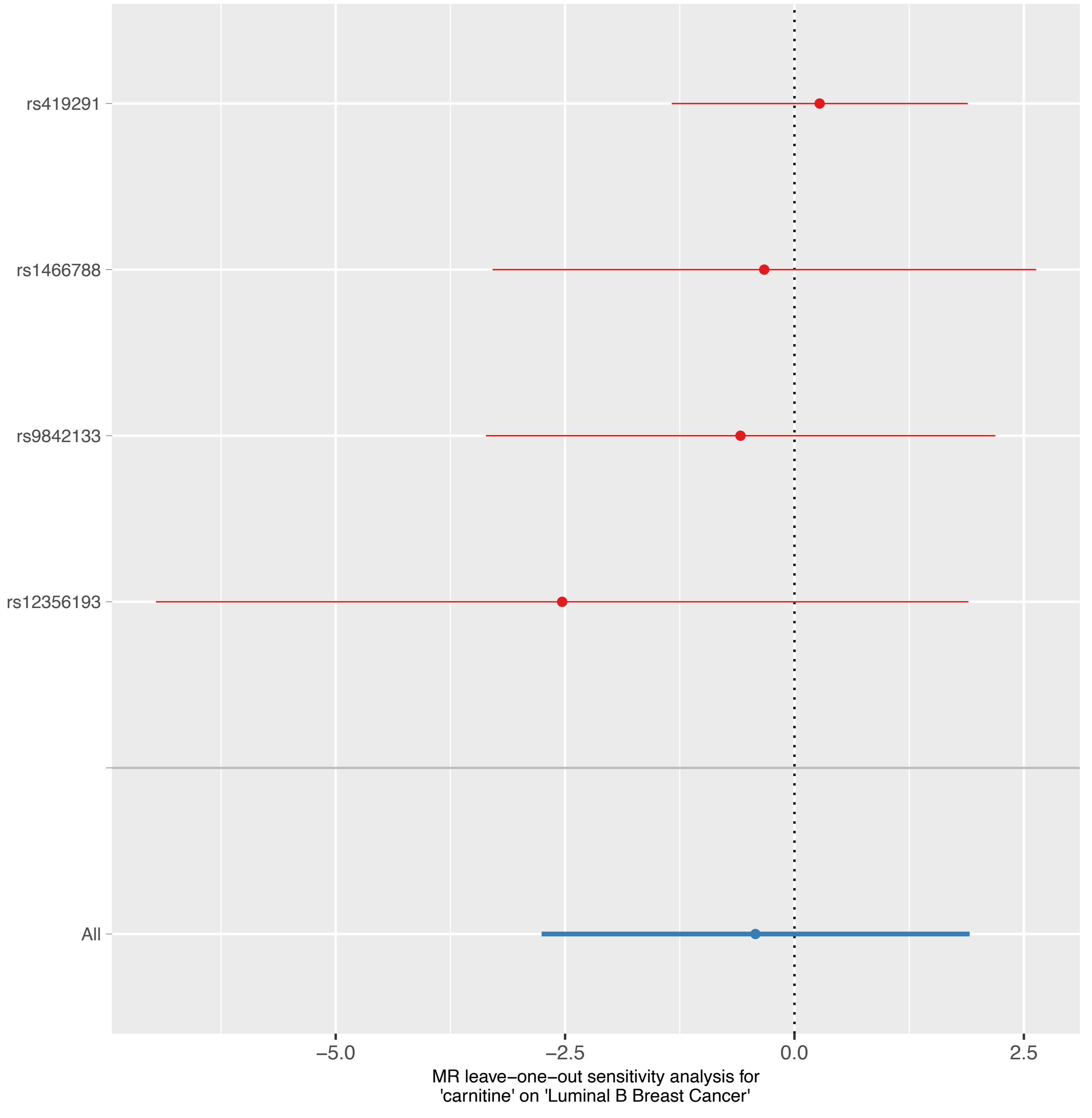
rs419291
rs1466788
rs9842133
rs12356193
All
−5.0
−2.5
0.0
2.5
MR leave−one−out sensitivity analysis for
'carnitine' on 'Luminal B Breast Cancer'

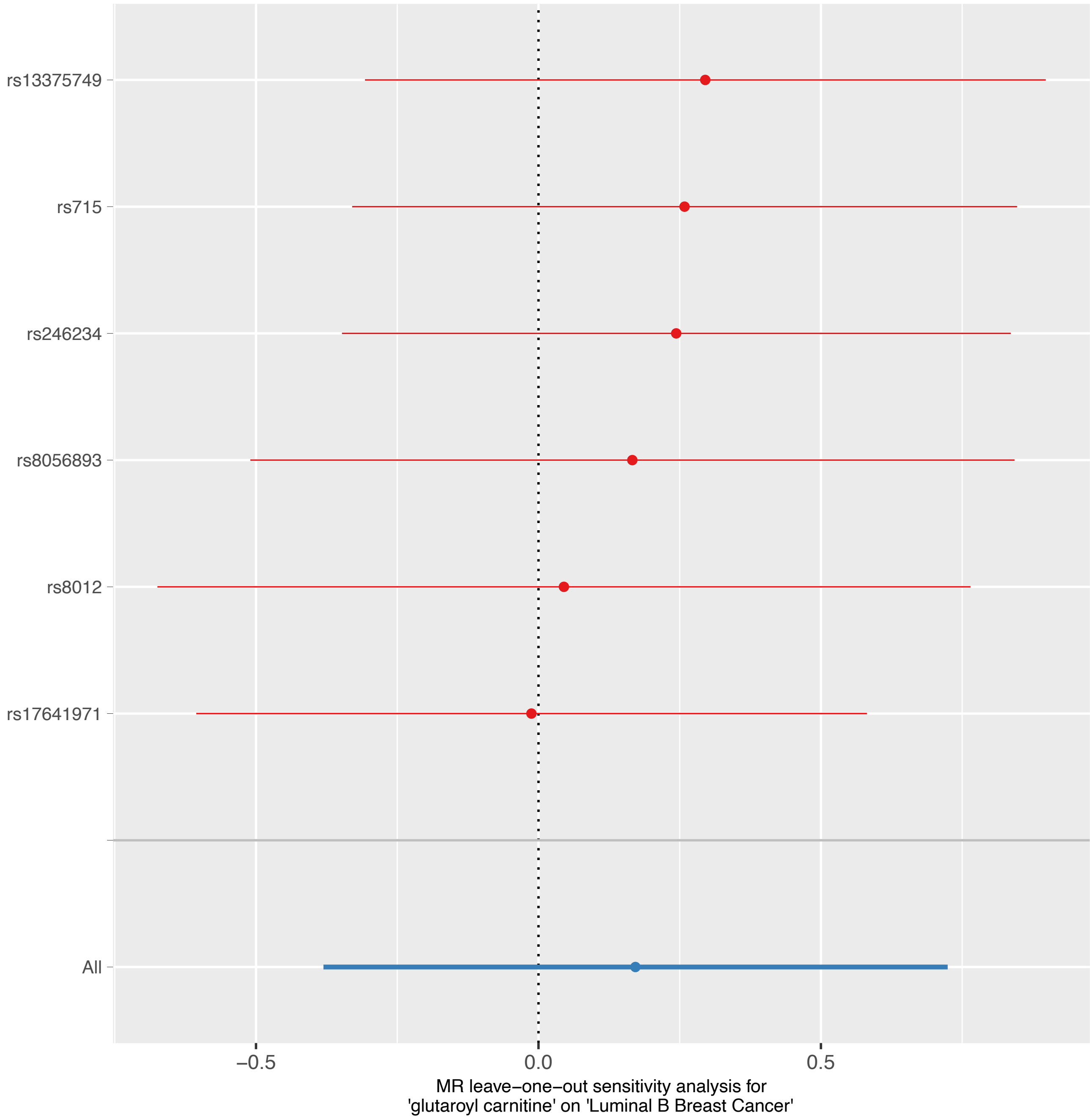

rs13375749
rs715
rs246234
rs8056893
rs8012
rs17641971
All
−0.5
0.0
0.5
MR leave−one−out sensitivity analysis for
'glutaroyl carnitine' on 'Luminal B Breast Cancer'

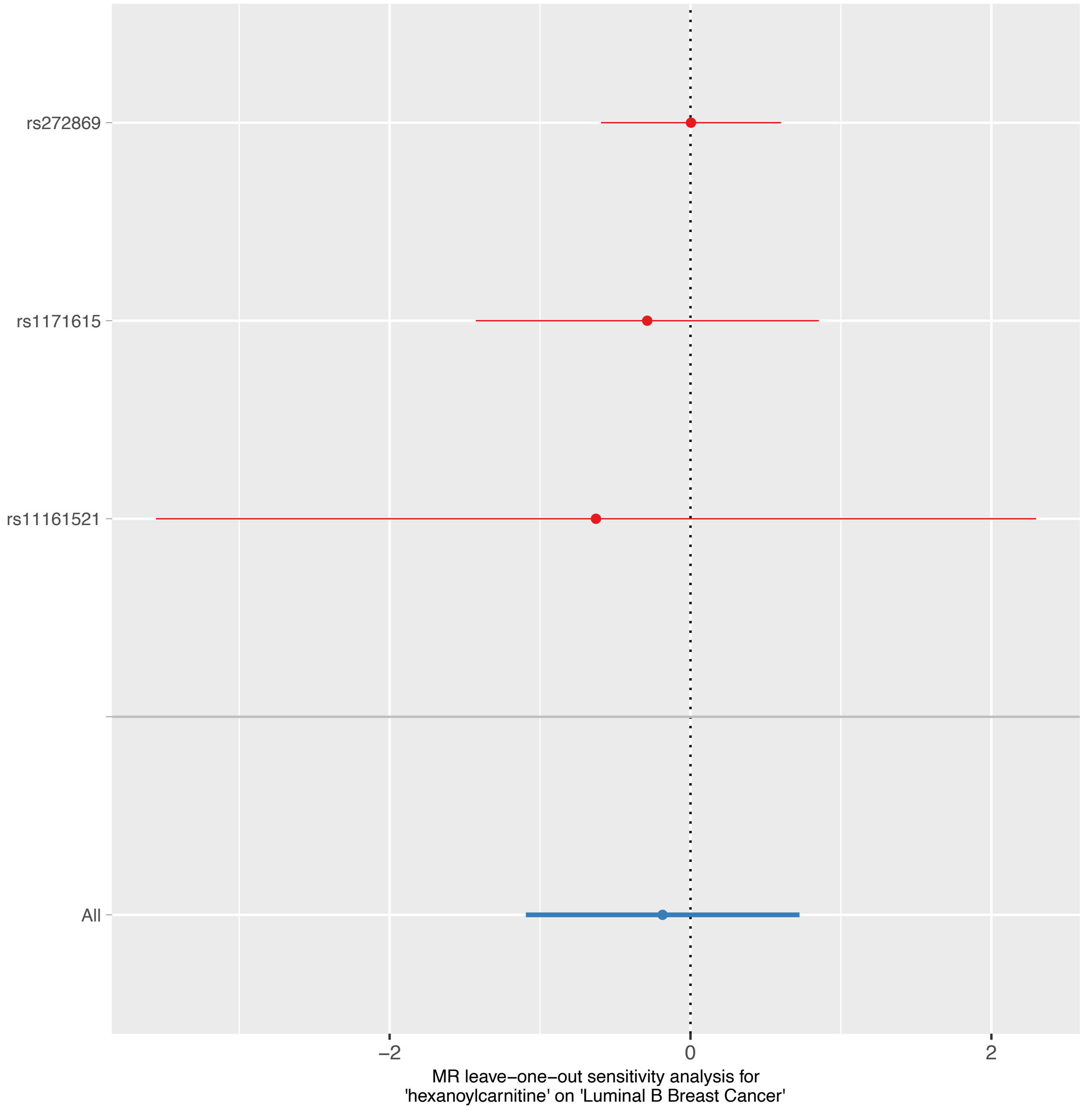
rs272869
rs1171615
rs11161521
All
−2
0
2
MR leave−one−out sensitivity analysis for
'hexanoylcarnitine' on 'Luminal B Breast Cancer'

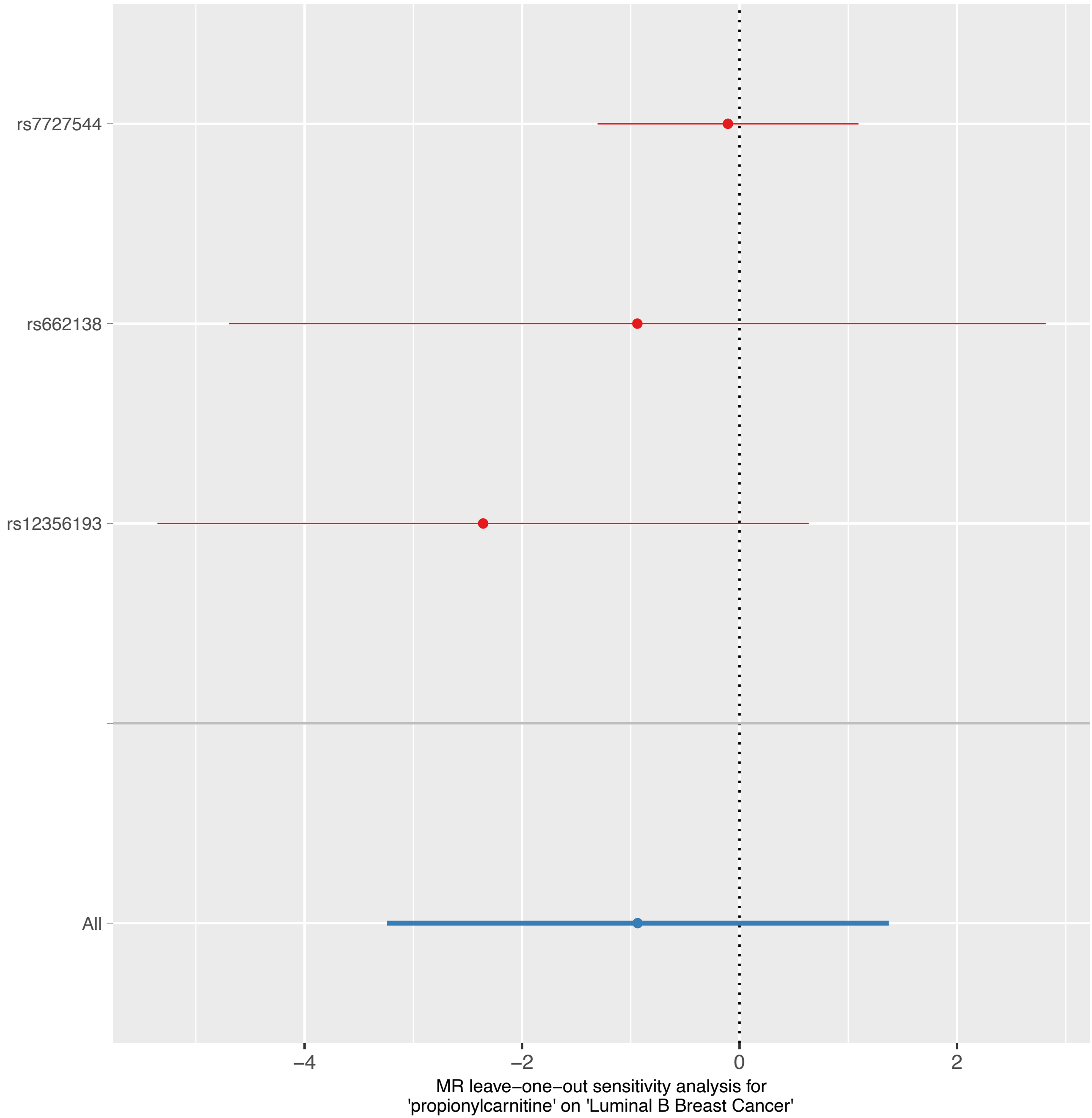
rs7727544
rs662138
rs12356193
All
−4
−2
0
2
MR leave−one−out sensitivity analysis for
'propionylcarnitine' on 'Luminal B Breast Cancer'

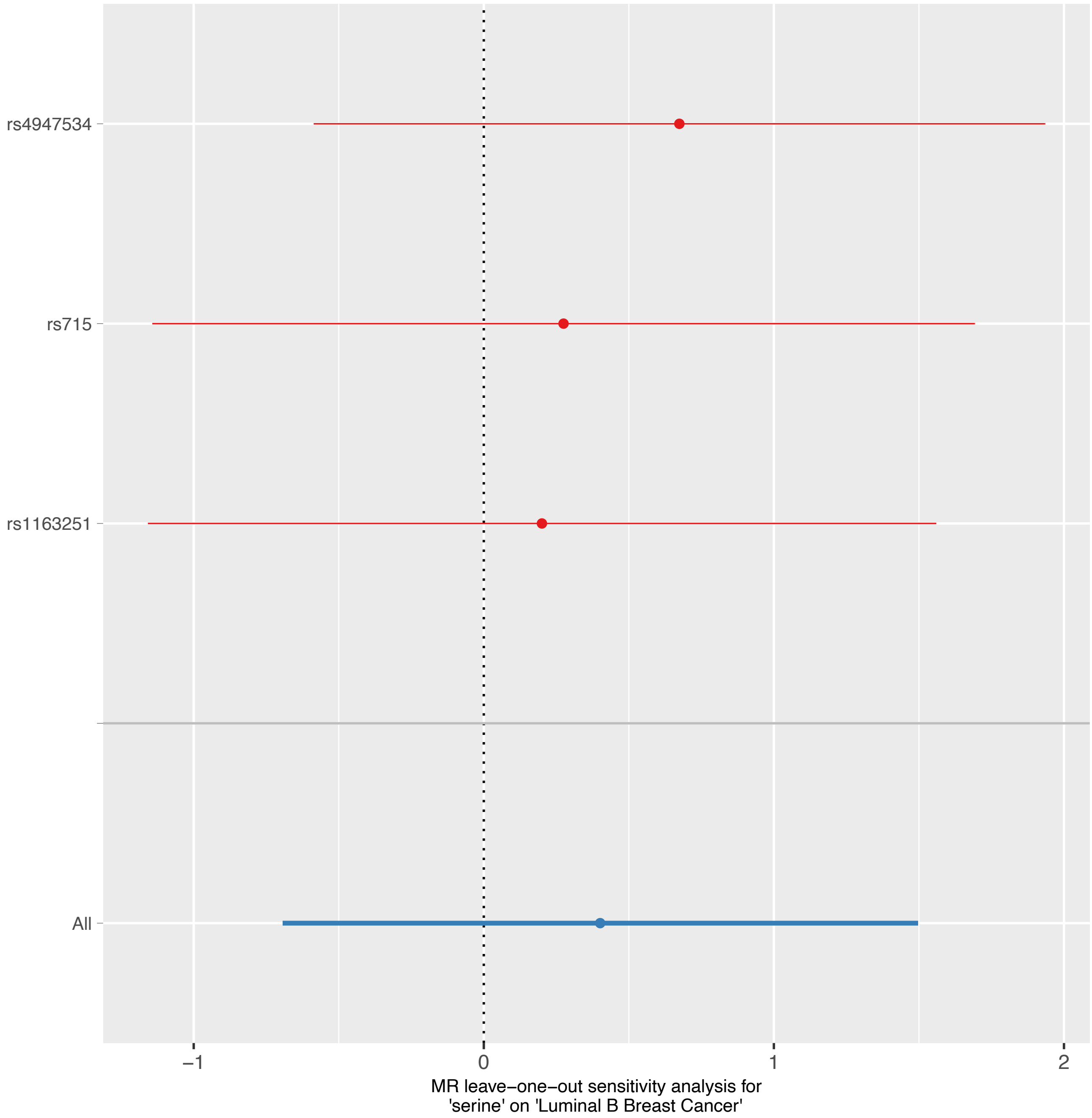

rs4947534
rs715
rs1163251
All
−1
0
1
2
MR leave−one−out sensitivity analysis for
'serine' on 'Luminal B Breast Cancer'

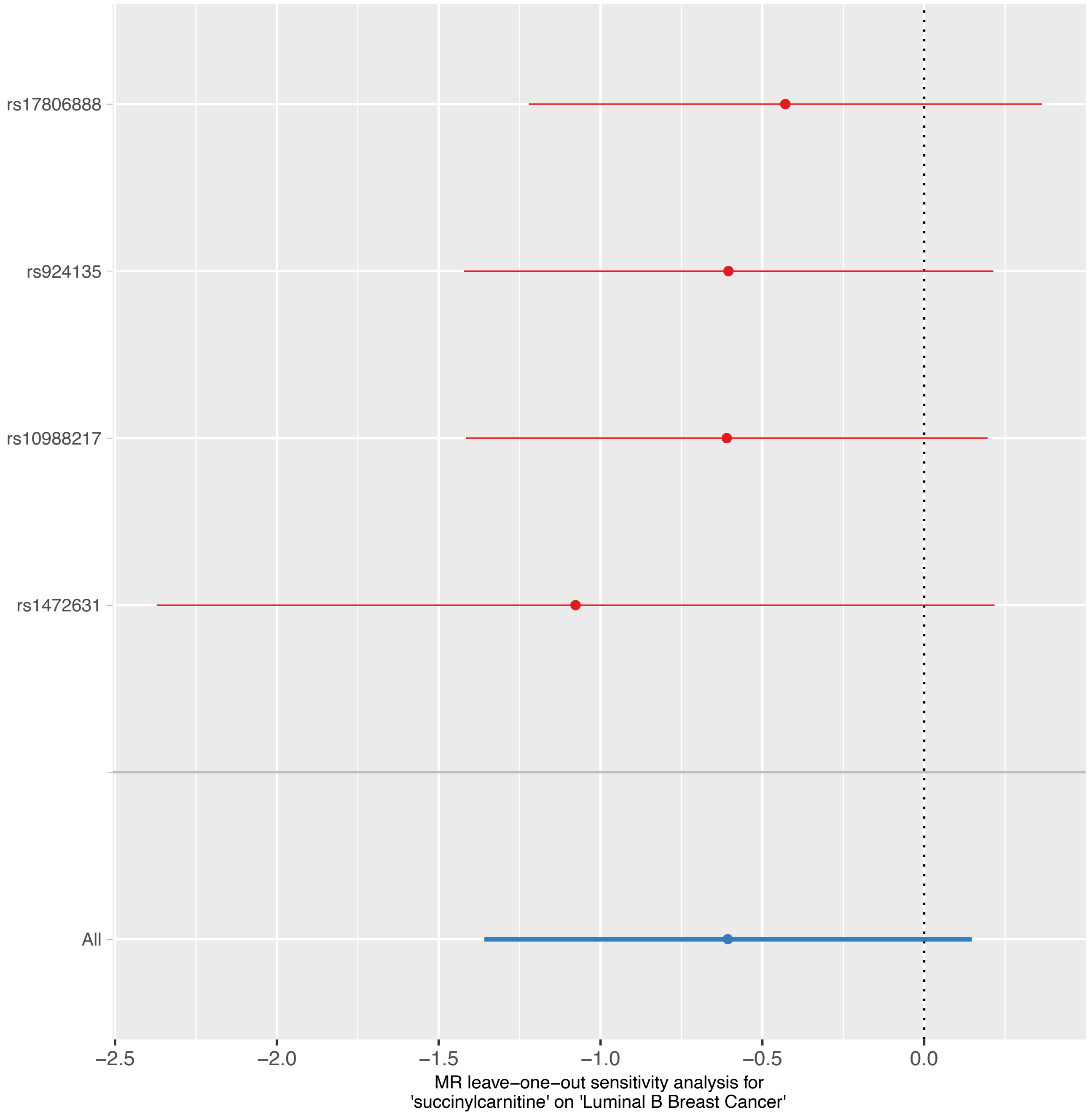

rs17806888
rs924135
rs10988217
rs1472631
All
−2.5
−2.0
−1.5
−1.0
−0.5
0.0
MR leave−one−out sensitivity analysis for
'succinylcarnitine' on 'Luminal B Breast Cancer'

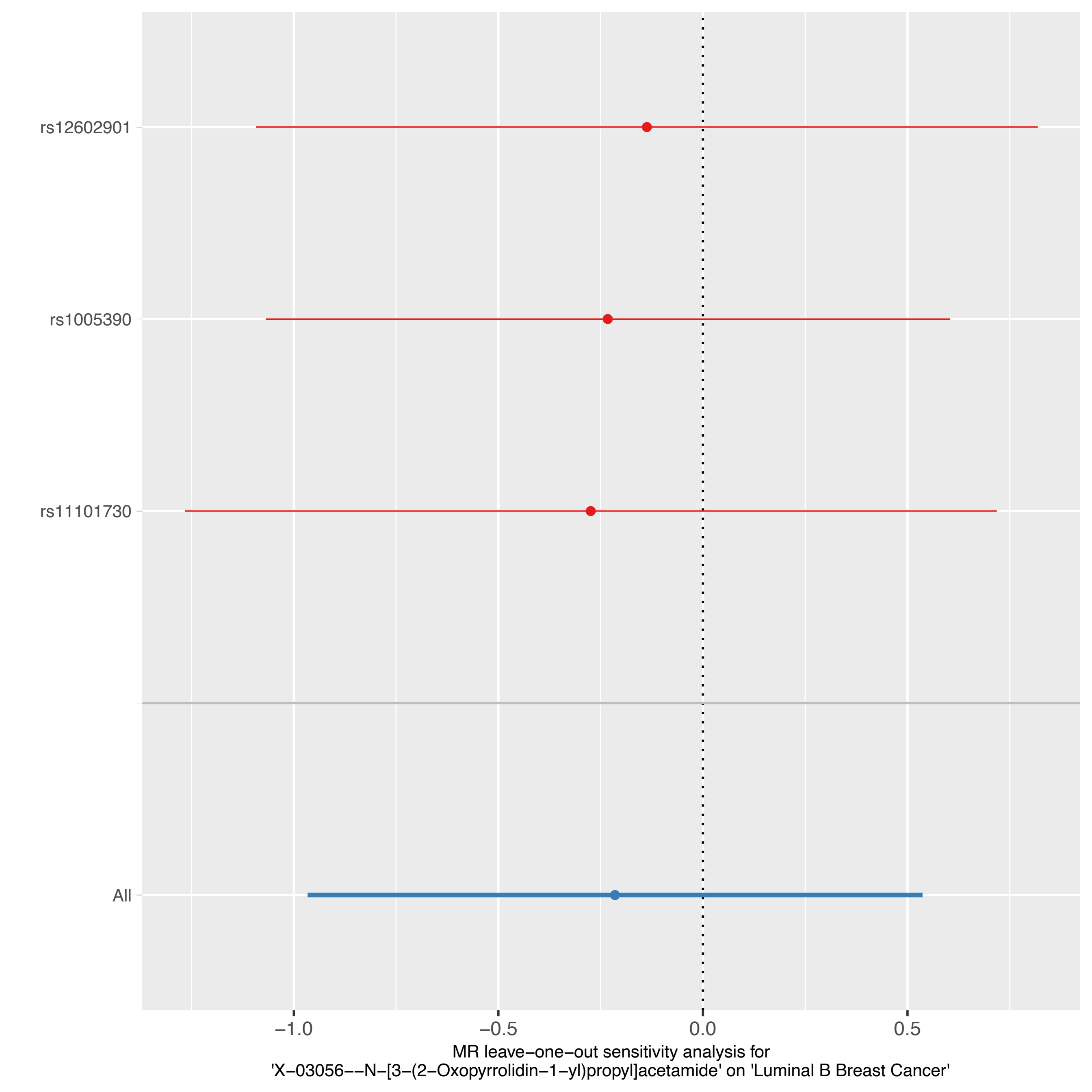

rs12602901
rs1005390
rs11101730
All
−1.0
−0.5
0.0
0.5
MR leave−one−out sensitivity analysis for
'X−03056−−N−[3−(2−Oxopyrrolidin−1−yl)propyl]acetamide' on 'Luminal B Breast Cancer'

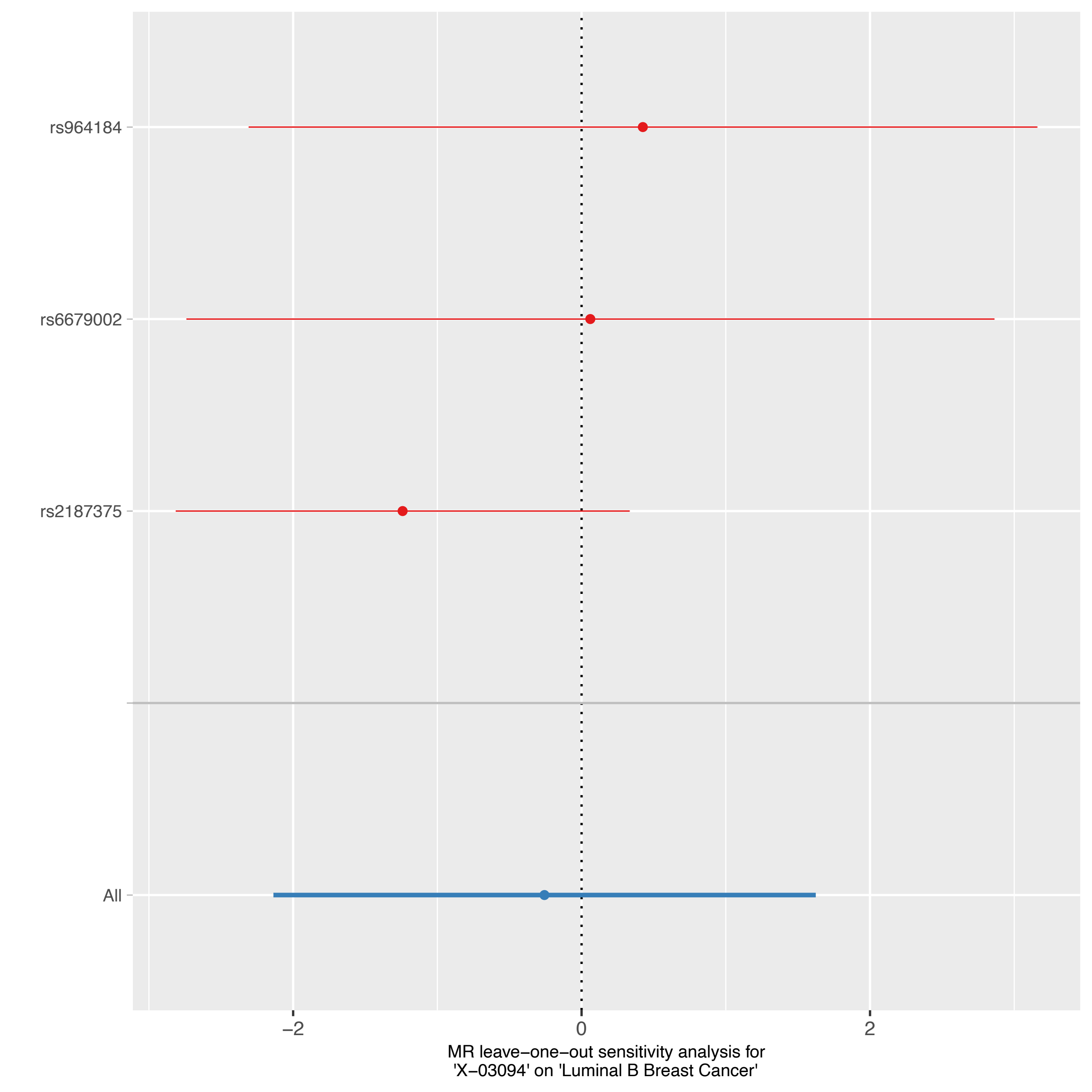
rs964184
rs6679002
rs2187375
All
−2
0
2
MR leave−one−out sensitivity analysis for
'X−03094' on 'Luminal B Breast Cancer'

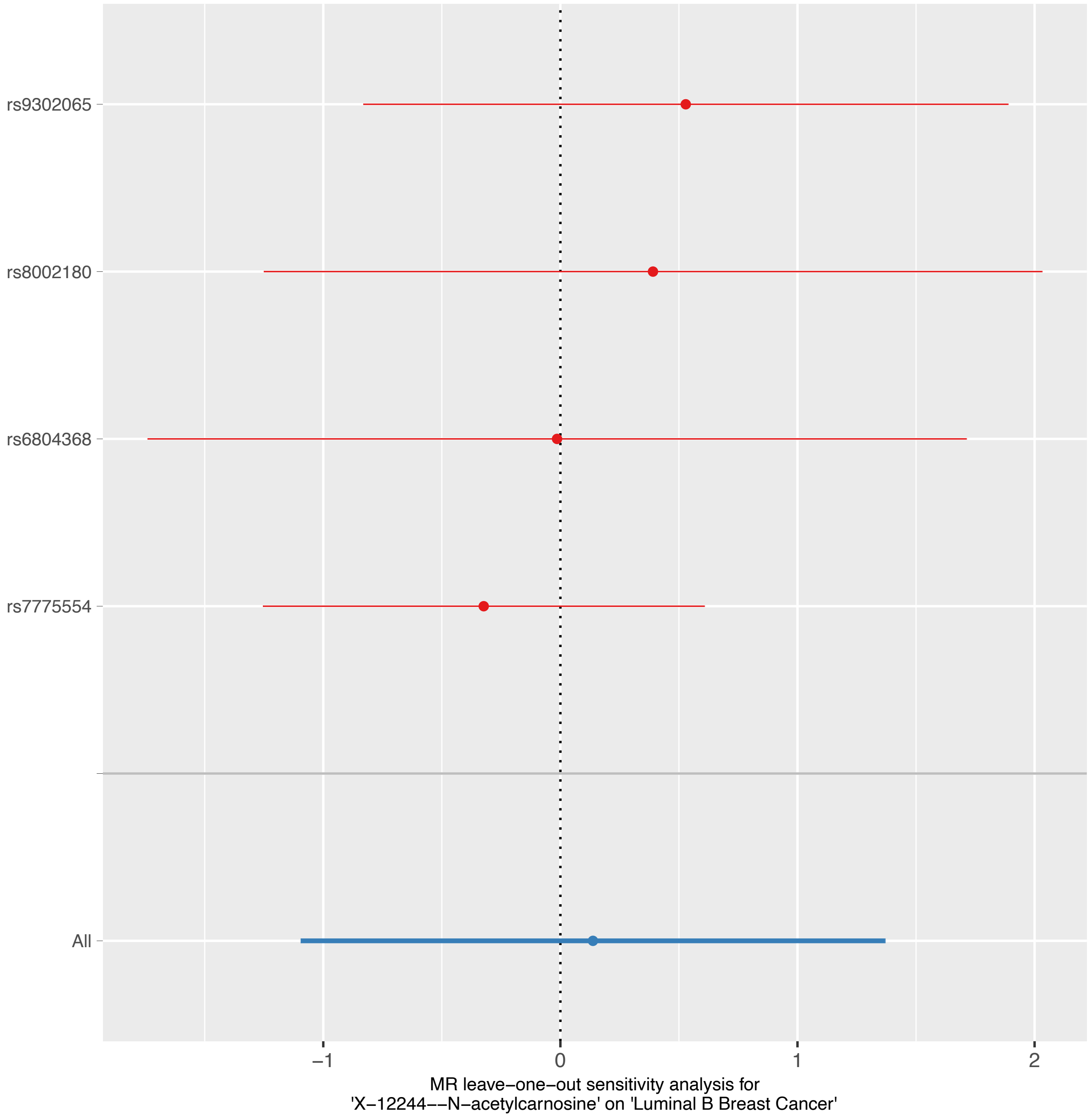
rs9302065
rs8002180
rs6804368
rs7775554
All
−1
0
1
2
MR leave−one−out sensitivity analysis for
'X−12244−−N−acetylcarnosine' on 'Luminal B Breast Cancer'

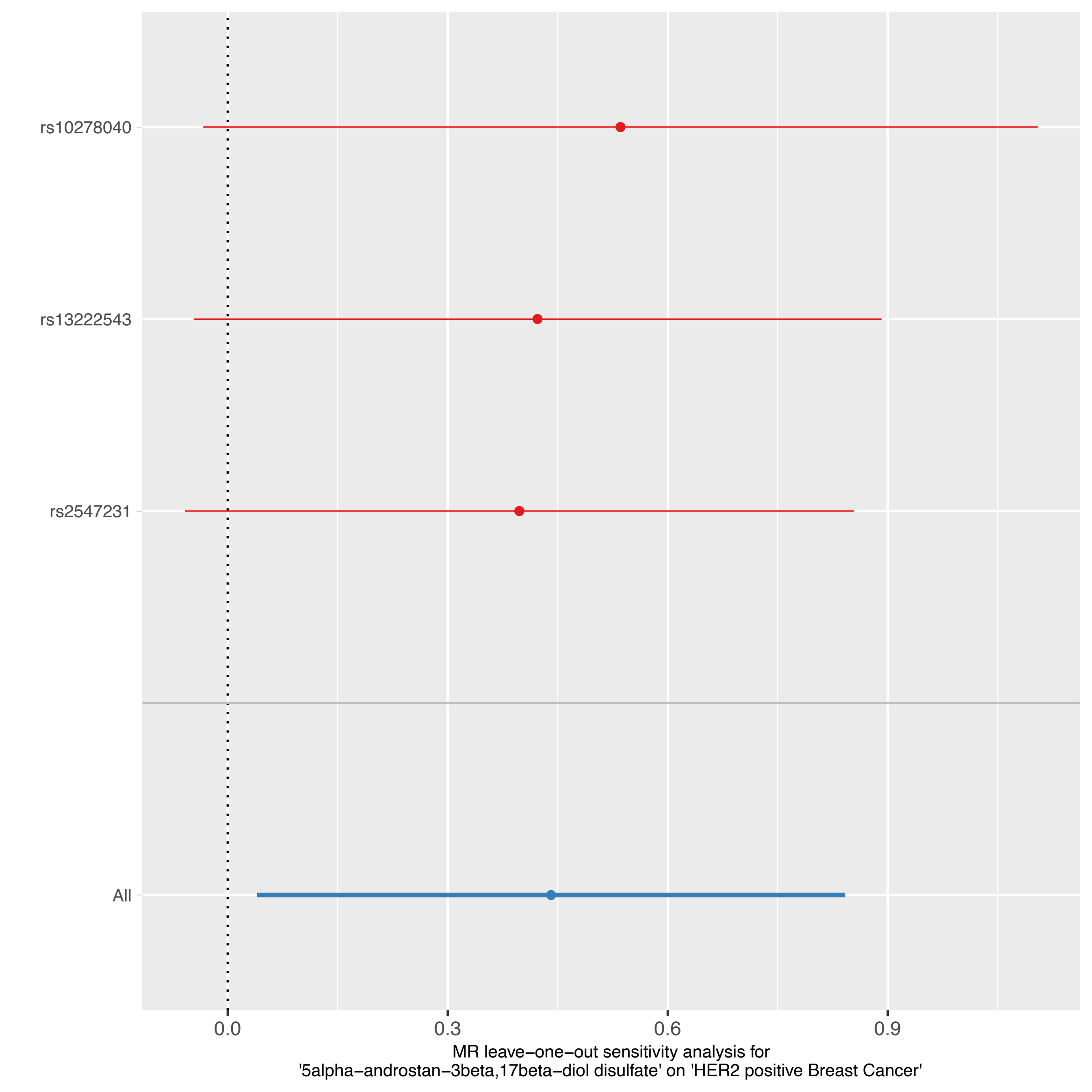

rs10278040
rs13222543
rs2547231
All
0.0
0.3
0.6
0.9
MR leave−one−out sensitivity analysis for
'5alpha−androstan−3beta,17beta−diol disulfate' on 'HER2 positive Breast Cancer'

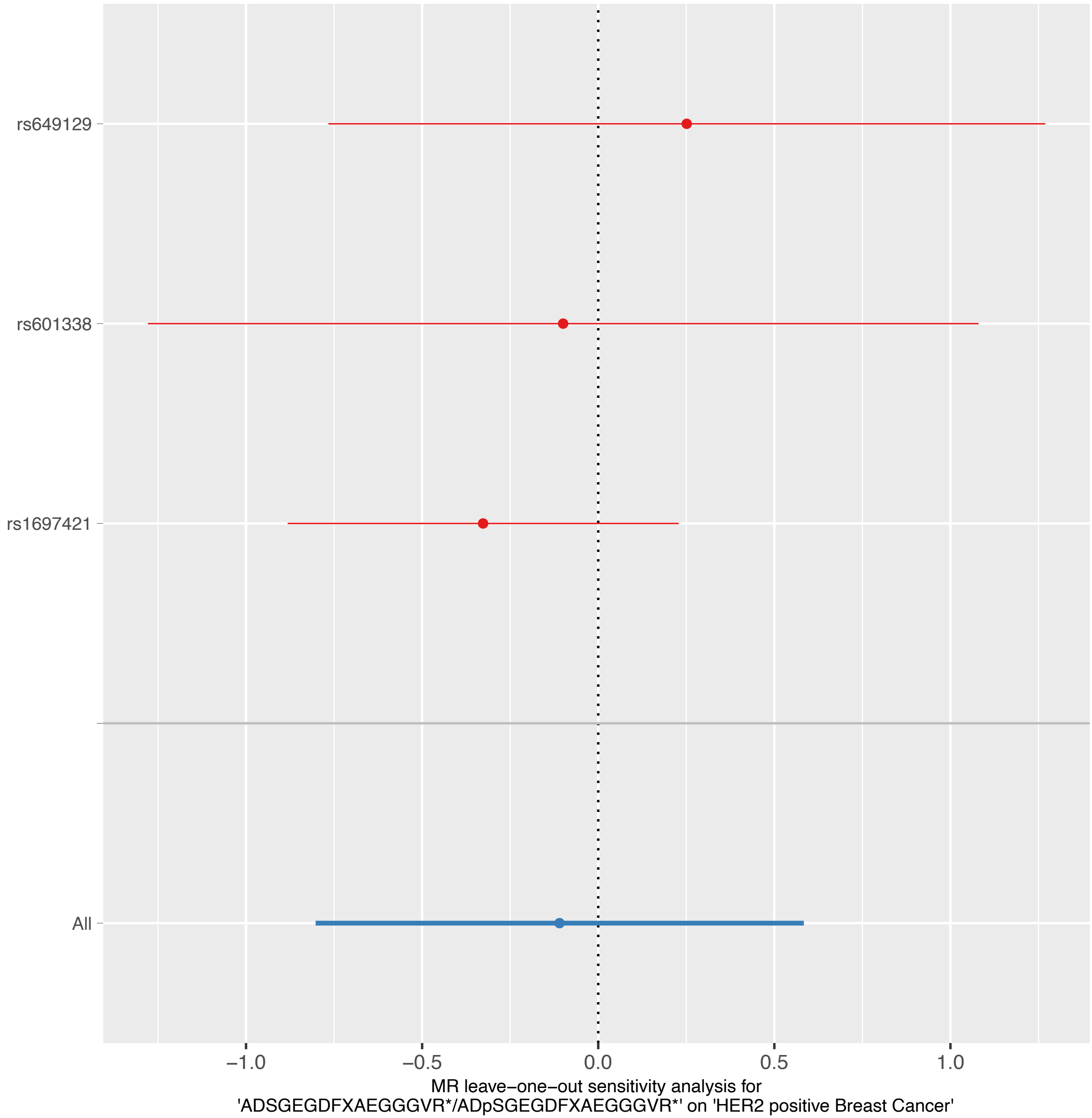

rs649129
rs601338
rs1697421
All
−1.0
−0.5
0.0
0.5
1.0
MR leave−one−out sensitivity analysis for
'ADSGEGDFXAEGGGVR*/ADpSGEGDFXAEGGGVR*' on 'HER2 positive Breast Cancer'

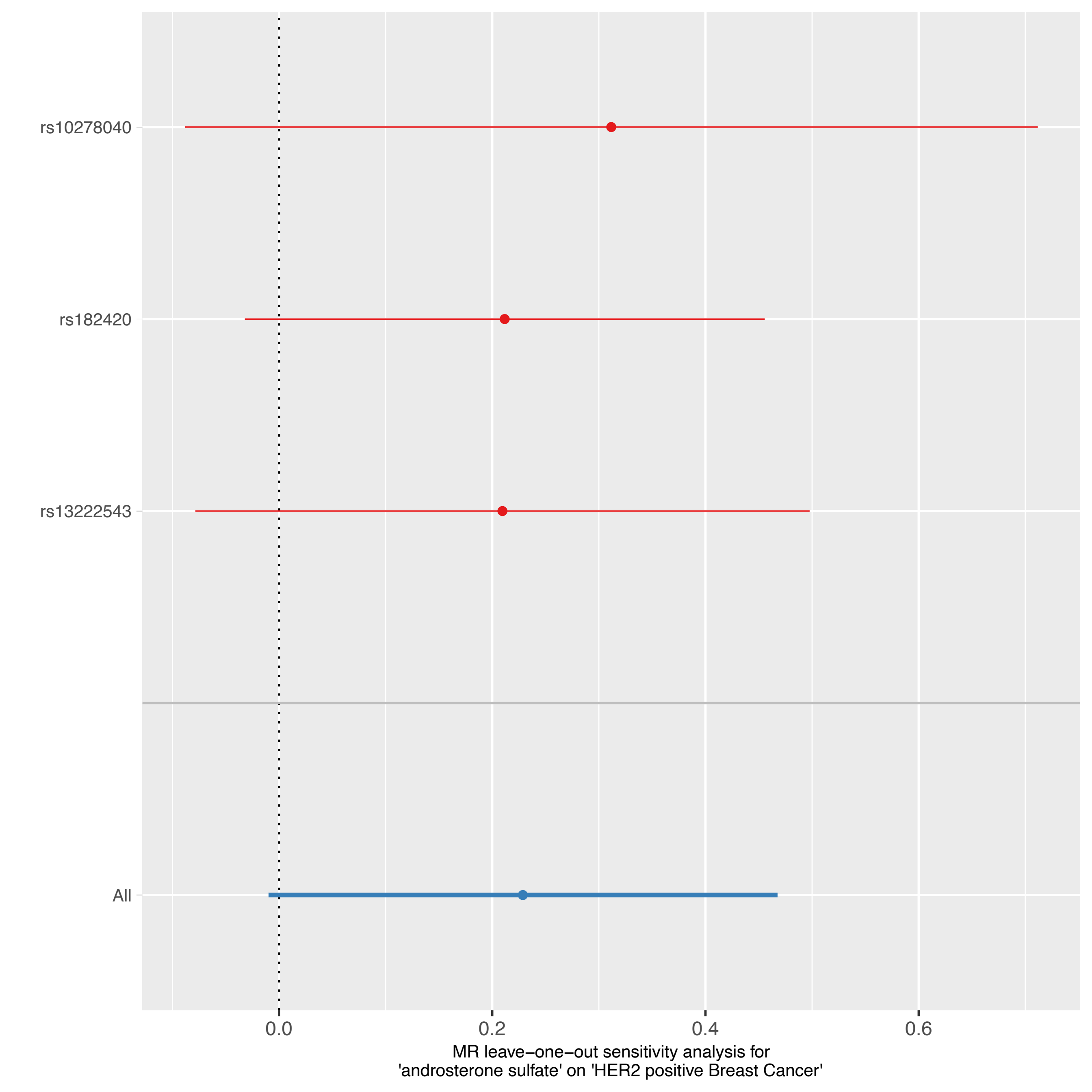
rs10278040
rs182420
rs13222543
All
0.0
0.2
0.4
0.6
MR leave−one−out sensitivity analysis for
'androsterone sulfate' on 'HER2 positive Breast Cancer'

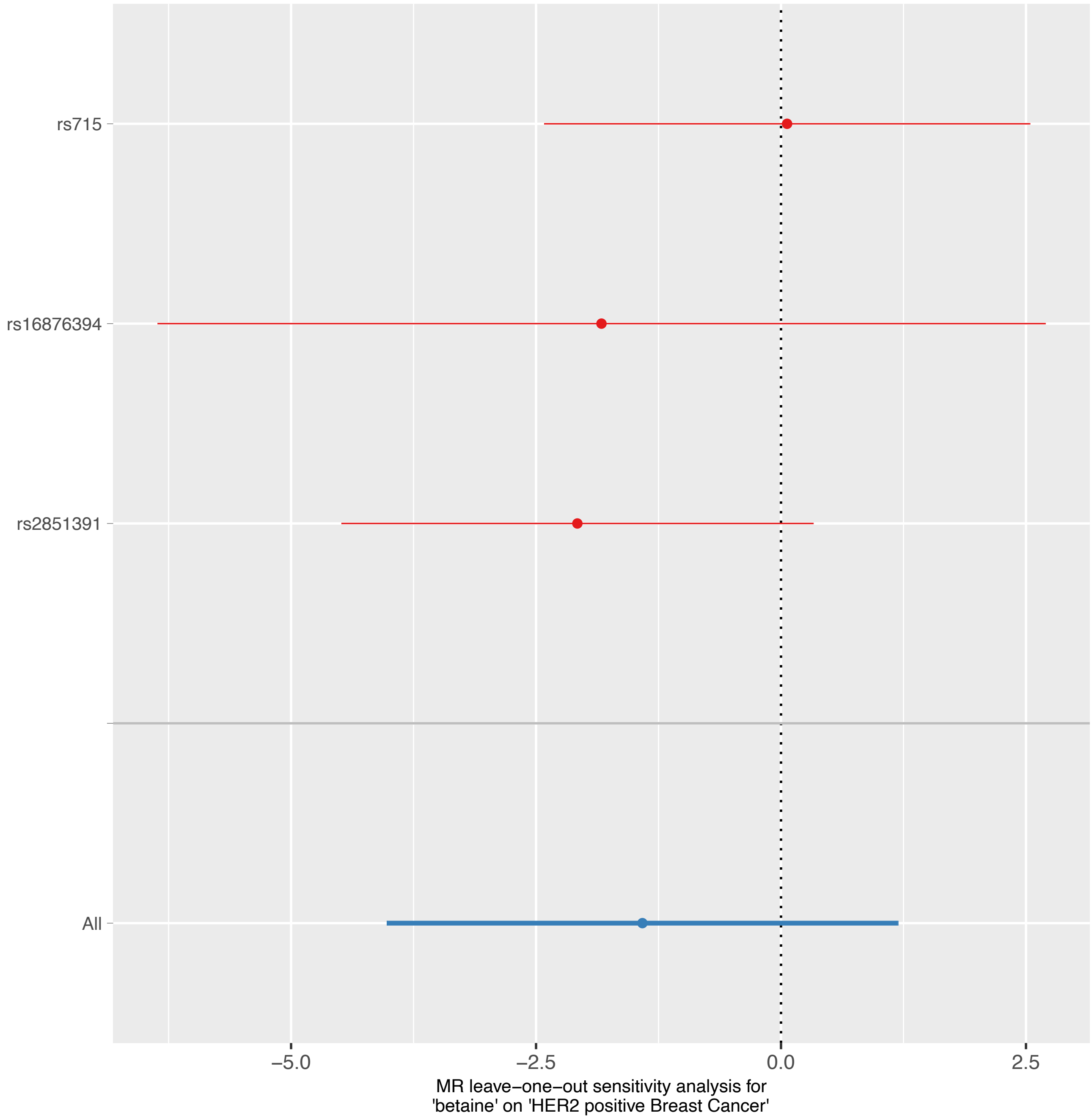
rs715
rs16876394
rs2851391
All
−5.0
−2.5
0.0
2.5
MR leave−one−out sensitivity analysis for
'betaine' on 'HER2 positive Breast Cancer'

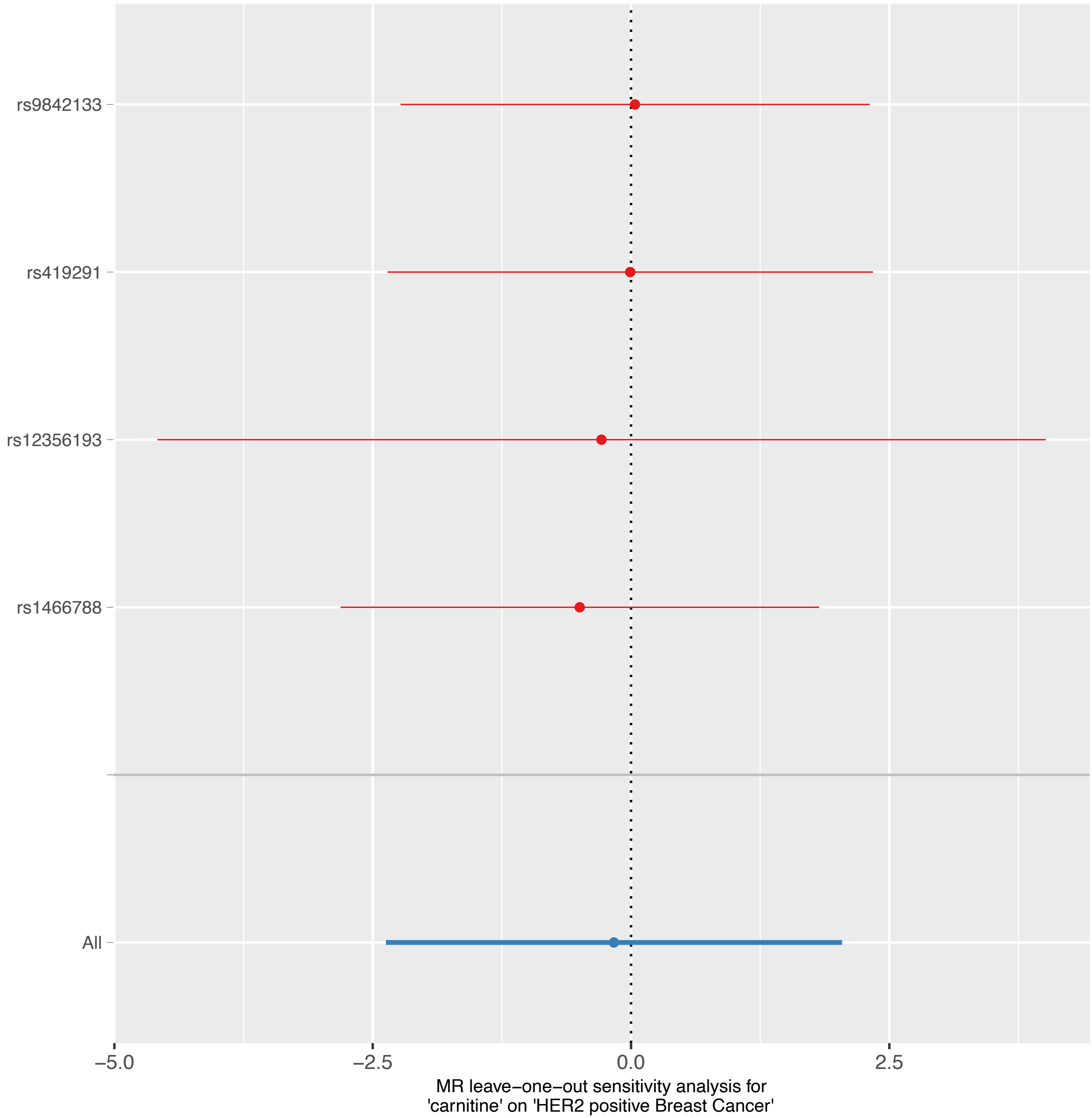
rs9842133
rs419291
rs12356193
rs1466788
All
−5.0
−2.5
0.0
2.5
MR leave−one−out sensitivity analysis for
'carnitine' on 'HER2 positive Breast Cancer'

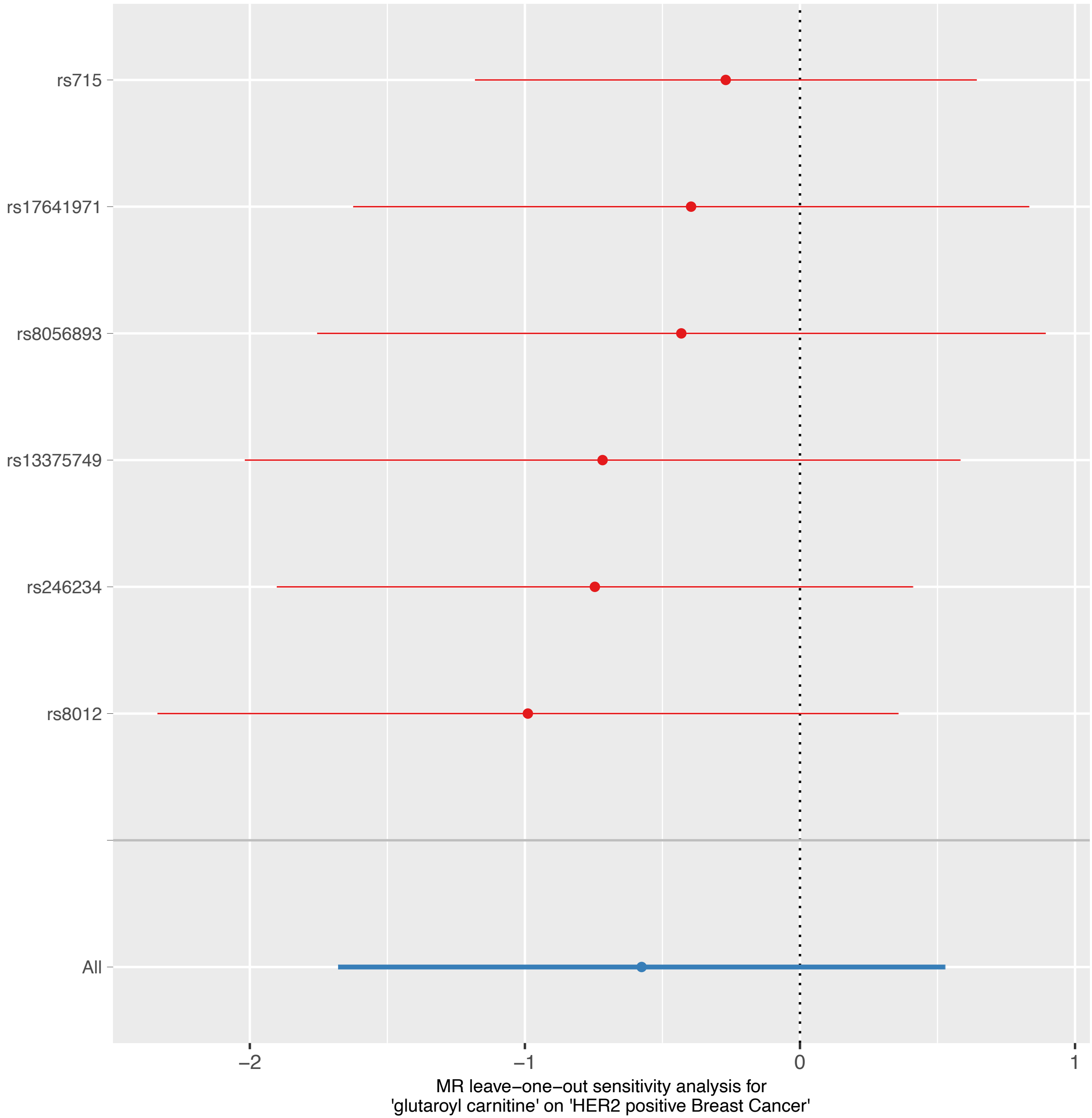

rs715
rs17641971
rs8056893
rs13375749
rs246234
rs8012
All
−2
−1
0
1
MR leave−one−out sensitivity analysis for
'glutaroyl carnitine' on 'HER2 positive Breast Cancer'

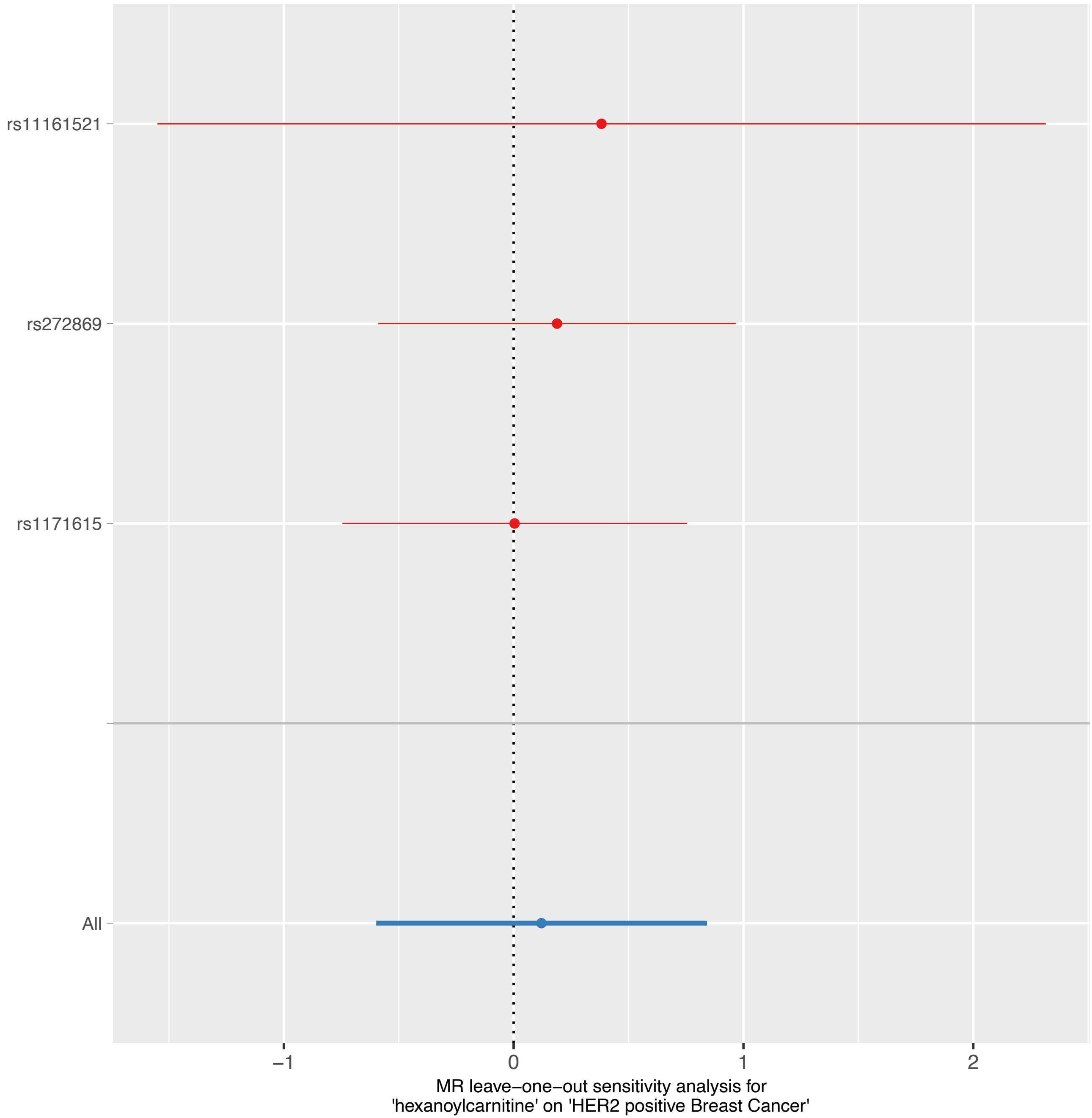

rs11161521
rs272869
rs1171615
All
−1
0
1
2
MR leave−one−out sensitivity analysis for
'hexanoylcarnitine' on 'HER2 positive Breast Cancer'

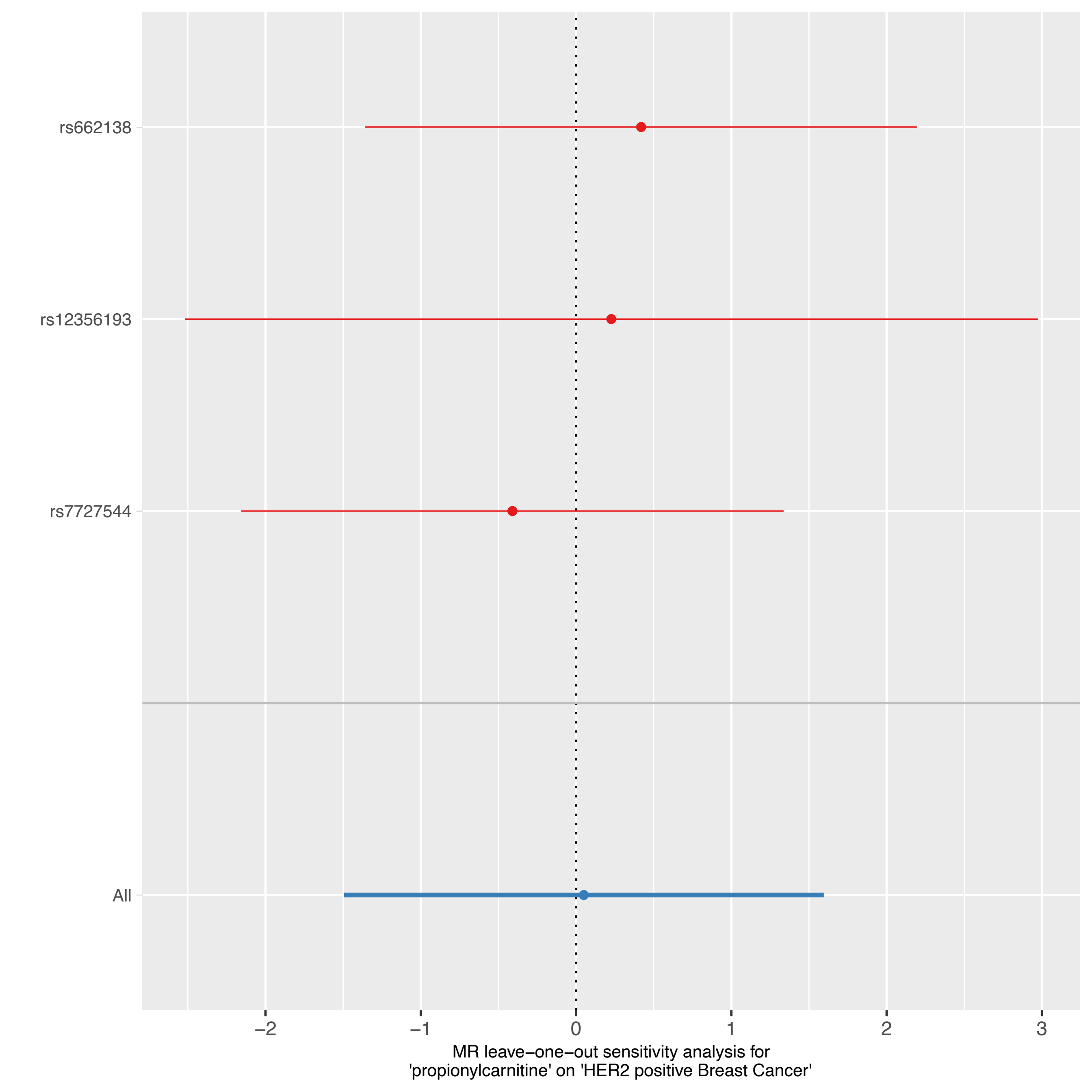

rs662138
rs12356193
rs7727544
All
−2
−1
0
1
2
3
MR leave−one−out sensitivity analysis for
'propionylcarnitine' on 'HER2 positive Breast Cancer'

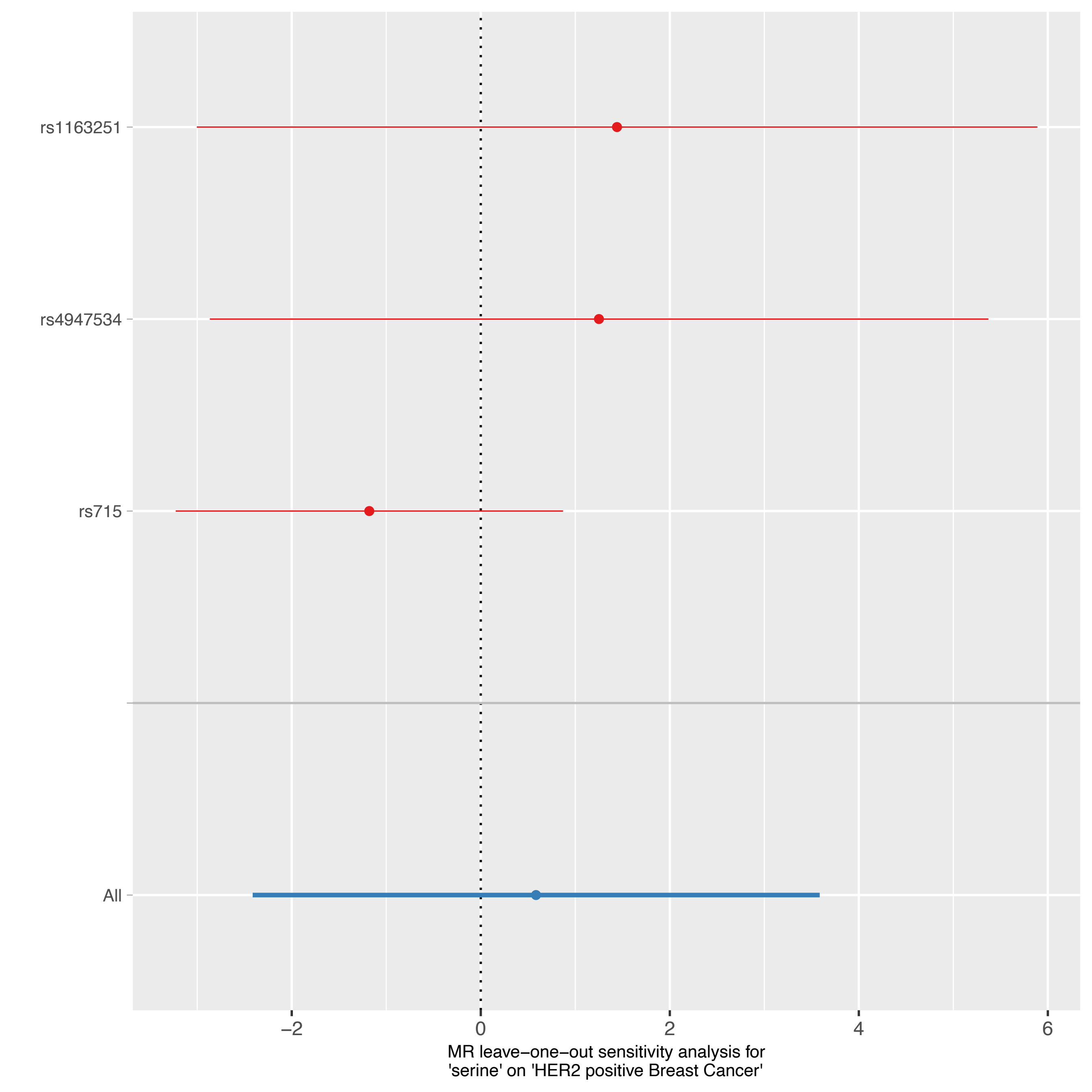
rs1163251
rs4947534
rs715
All
−2
0
2
4
6
MR leave−one−out sensitivity analysis for
'serine' on 'HER2 positive Breast Cancer'

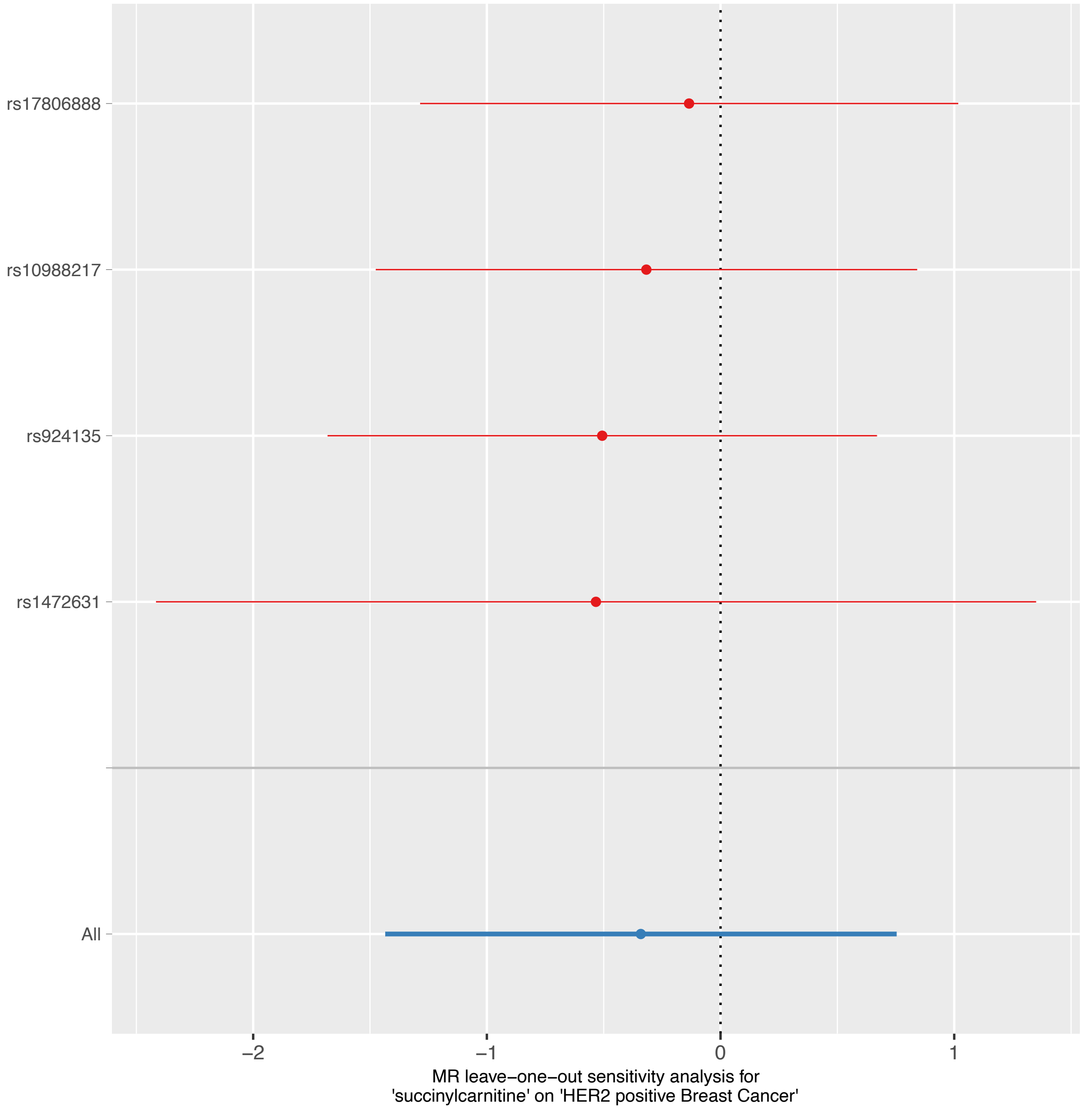

rs17806888
rs10988217
rs924135
rs1472631
All
−2
−1
0
1
MR leave−one−out sensitivity analysis for
'succinylcarnitine' on 'HER2 positive Breast Cancer'

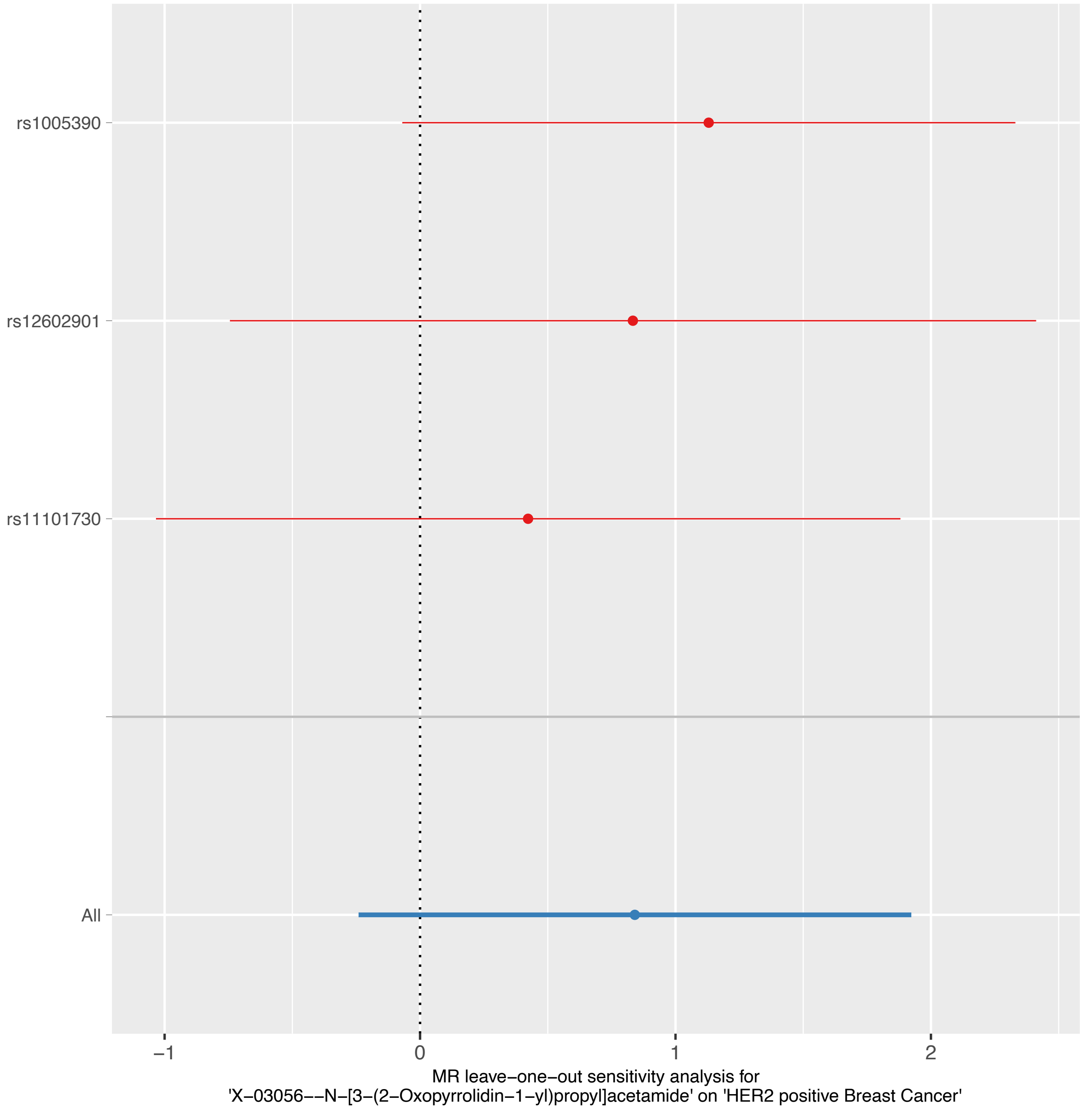
rs1005390
rs12602901
rs11101730
All
−1
0
1
2
MR leave−one−out sensitivity analysis for
'X−03056−−N−[3−(2−Oxopyrrolidin−1−yl)propyl]acetamide' on 'HER2 positive Breast Cancer'

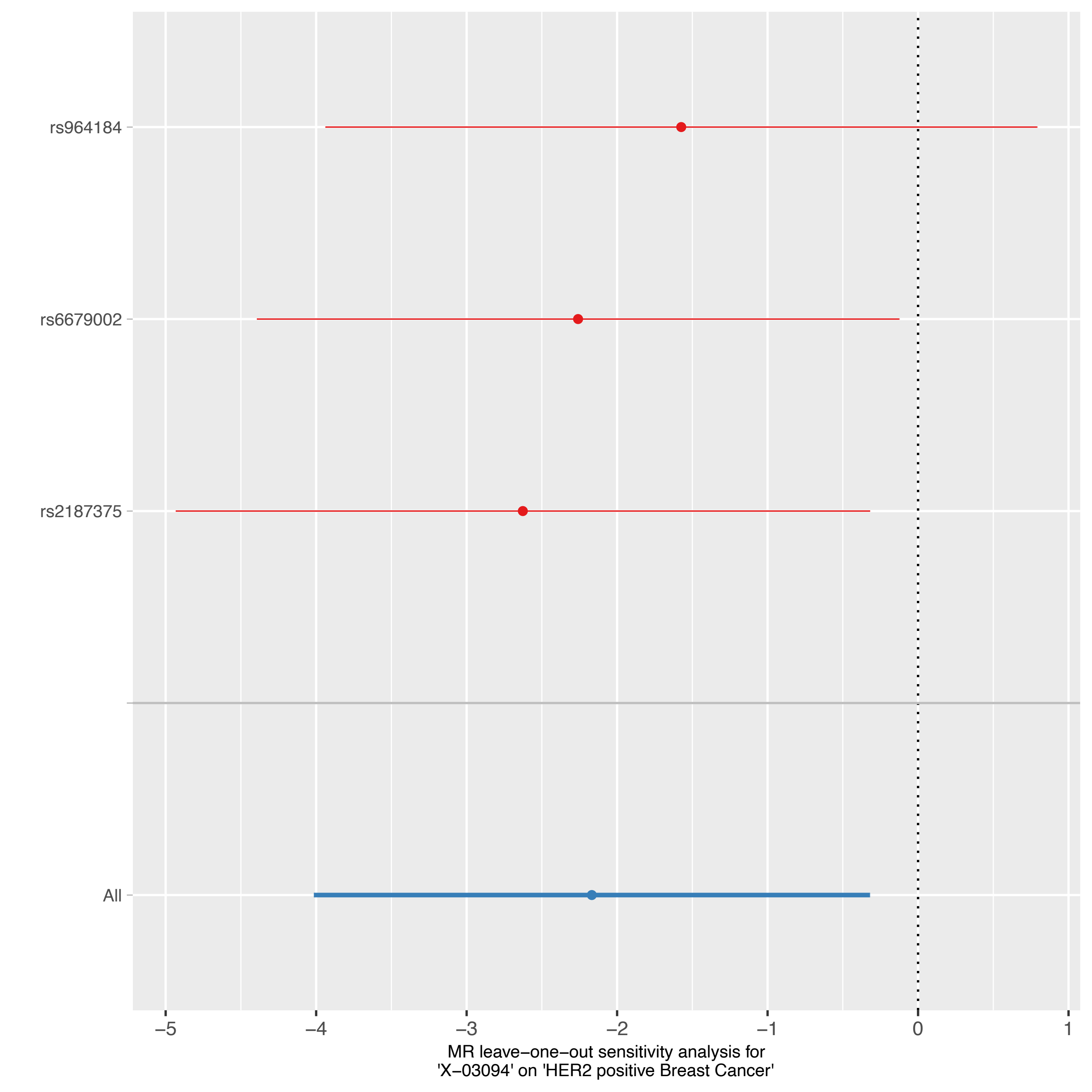

rs964184
rs6679002
rs2187375
All
−5
−4
−3
−2
−1
0
1
MR leave−one−out sensitivity analysis for
'X−03094' on 'HER2 positive Breast Cancer'

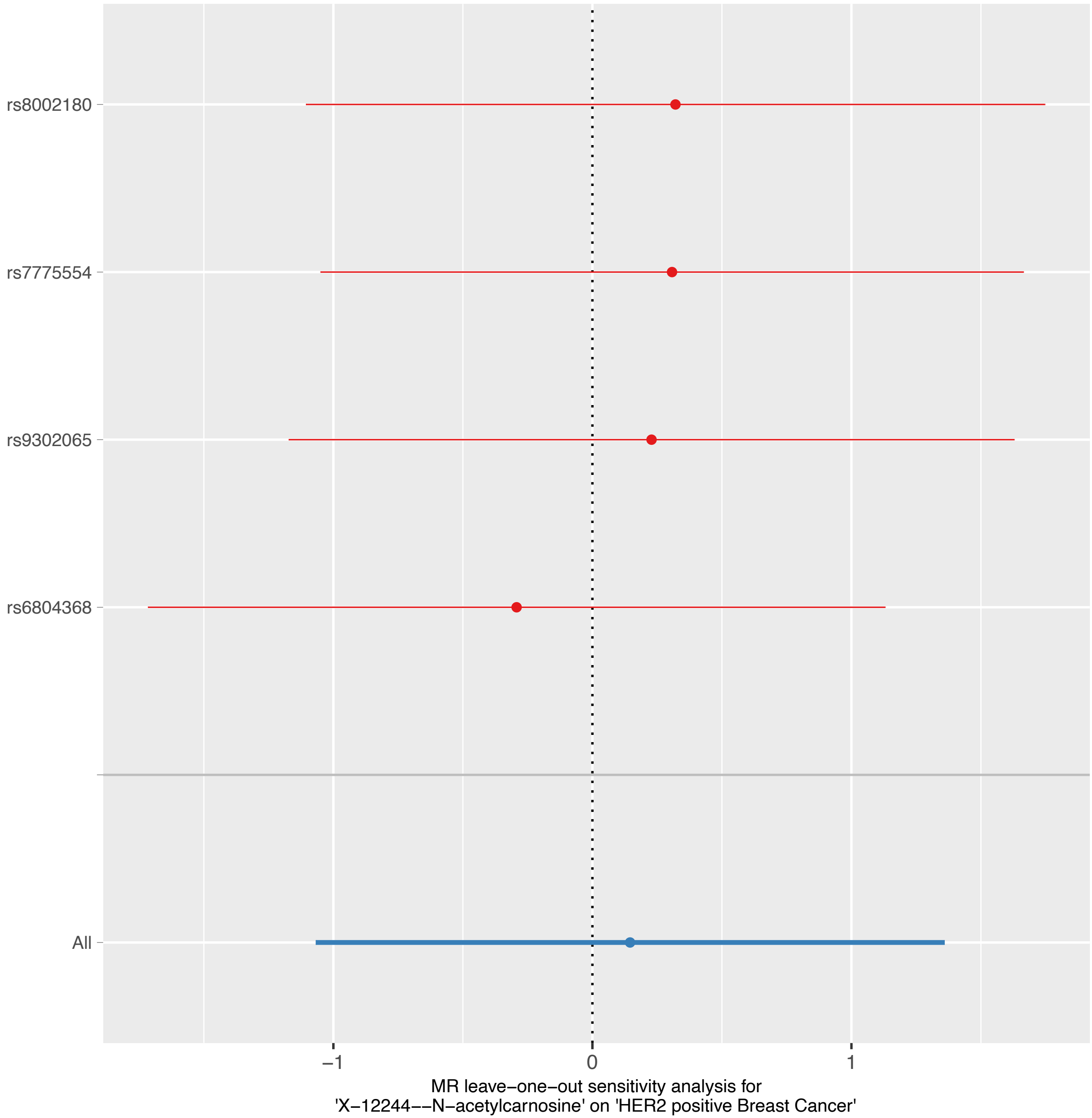

rs8002180
rs7775554
rs9302065
rs6804368
All
−1
0
1
MR leave−one−out sensitivity analysis for
'X−12244−−N−acetylcarnosine' on 'HER2 positive Breast Cancer'
