## Supplementary material for "Exploring the causal role of the human gut microbiome in breast cancer risk": STROBE-MR Reporting

**STROBE-MR checklist of recommended items to address in reports of Mendelian randomization studies**^1^ ^2^

| **Item No.** | **Section** | **Checklist item** | **Page No.** | **Relevant text from manuscript** |
| --- | --- | --- | --- | --- |
| 1 | **TITLE and ABSTRACT** | Indicate Mendelian randomization (MR) as the study’s design in the title and/or the abstract if that is a main purpose of the study | 1-2 | Abstract defines Mendelian randomization (MR) as study design |
|  | **INTRODUCTION** |  |  |  |
| 2 | **Background** | Explain the scientific background and rationale for the reported study. What is the exposure? Is a potential causal relationship between exposure and outcome plausible? Justify why MR is a helpful method to address the study question | 3-4 | Introduction states the background information relevant to breast cancer (BC) and its risk factors, existing studies of the microbiome and BC and their limitations, and MR as a study design |
| 3 | **Objectives** | State specific objectives clearly, including pre-specified causal hypotheses (if any). State that MR is a method that, under specific assumptions, intends to estimate causal effects | 4 | Last paragraph outlines the objectives |
|  | **METHODS** |  |  |  |
| 4 | **Study design and data sources** | Present key elements of the study design early in the article. Consider including a table listing sources of data for all phases of the study. For each data source contributing to the analysis, describe the following: |  |  |
|  | a) | Setting: Describe the study design and the underlying population, if possible. Describe the setting, locations, and relevant dates, including periods of recruitment, exposure, follow-up, and data collection, when available. | 4 | First paragraph of the methods and Figures 1-2 outline the study design |
|  | b) | Participants: Give the eligibility criteria, and the sources and methods of selection of participants. Report the sample size, and whether any power or sample size calculations were carried out prior to the main analysis | 5-8 | The “Breast cancer”, “Gut microbiome” and “Circulating metabolites” sections outline the studies used in this two-sample MR analysis |
|  | c) | Describe measurement, quality control and selection of genetic variants | 6-8 | Instrument selection for both the gut microbiome and circulating metabolites are described in sections named “Gut microbiome” and “Circulating metabolites” as well as in the first paragraph in the “Mendelian randomization” section of the Statistical analyses. |
|  | d) | For each exposure, outcome, and other relevant variables, describe methods of assessment and diagnostic criteria for diseases | 5-8 | The “Breast cancer”, “Gut microbiome” and “Circulating metabolites” sections outline the measures used and the studies from which the summary statistics were obtained |
|  | e) | Provide details of ethics committee approval and participant informed consent, if relevant | 5 | Ethical approval statement provided |
| 5 | **Assumptions** | Explicitly state the three core IV assumptions for the main analysis (relevance, independence and exclusion restriction) as well assumptions for any additional or sensitivity analysis | 8 | The first paragraph in the “Mendelian randomization” section of the Statistical analyses section states the assumptions |
| 6 | **Statistical methods: main analysis** | Describe statistical methods and statistics used | 8-11 | All MR and multivariable regression analyses described |
|  | a) | Describe how quantitative variables were handled in the analyses (i.e., scale, units, model) | 9 | Statement about how MR analysis units can be interpreted provided |
|  | b) | Describe how genetic variants were handled in the analyses and, if applicable, how their weights were selected | 8 | Description of the main analysis method (i.e., inverse variance weighted, IVW) provided |
|  | c) | Describe the MR estimator (e.g. two-stage least squares, Wald ratio) and related statistics. Detail the included covariates and, in case of two-sample MR, whether the same covariate set was used for adjustment in the two samples | 8 | Description of Wald ratio and IVW method provided |
|  | d) | Explain how missing data were addressed | 8 | Use of proxy single nucleotide polymorphisms (SNPs) were used where original SNPs were not present in outcome data |
|  | e) | If applicable, indicate how multiple testing was addressed | 9-10 | Overall comment on how p-values and effect estimates were used as continuous indicators of evidence strength provided on page 9; however, multiple testing corrections were made for MR analyses of metabolites on BC (page 9) and with linear regression analyses (on page 10) |
| 7 | **Assessment of assumptions** | Describe any methods or prior knowledge used to assess the assumptions or justify their validity | 11 | First paragraph in “Sensitivity analyses” section describes all analyses undertaken to test robustness of estimates to MR assumptions |
| 8 | **Sensitivity analyses and additional analyses** | Describe any sensitivity analyses or additional analyses performed (e.g. comparison of effect estimates from different approaches, independent replication, bias analytic techniques, validation of instruments, simulations) | 11-13 | All sensitivity analyses described |
| 9 | **Software and pre-registration** |  |  |  |
|  | a) | Name statistical software and package(s), including version and settings used | 13 | Software and versions provided alongside a Github page for scripts |
|  | b) | State whether the study protocol and details were pre-registered (as well as when and where) | NA | Not provided |
|  | **RESULTS** |  |  |  |
| 10 | **Descriptive data** |  |  |  |
|  | a) | Report the numbers of individuals at each stage of included studies and reasons for exclusion. Consider use of a flow diagram | 13 | Number of SNPs provided and Figure 2 provides numbers of individuals included in analyses |
|  | b) | Report summary statistics for phenotypic exposure(s), outcome(s), and other relevant variables (e.g. means, SDs, proportions) | NA | Not provided but SNPs used as instruments provided in Supplementary Tables 1 and 2 |
|  | c) | If the data sources include meta-analyses of previous studies, provide the assessments of heterogeneity across these studies | NA | Not provided |
|  | d) | For two-sample MR:  i.  Provide justification of the similarity of the genetic variant-exposure associations between the exposure and outcome samples  ii.  Provide information on the number of individuals who overlap between the exposure and outcome studies | 13 | Explicit statement about the independence and homogeneity of data sources provided |
| 11 | **Main results** |  |  |  |
|  | a) | Report the associations between genetic variant and exposure, and between genetic variant and outcome, preferably on an interpretable scale | Tables | The SNP-exposure associations provided in Supplementary Tables 1 and 2 |
|  | b) | Report MR estimates of the relationship between exposure and outcome, and the measures of uncertainty from the MR analysis, on an interpretable scale, such as odds ratio or relative risk per SD difference | 13-17 | Main results provided and interpreted |
|  | c) | If relevant, consider translating estimates of relative risk into absolute risk for a meaningful time period | NA | Not provided |
|  | d) | Consider plots to visualize results (e.g. forest plot, scatterplot of associations between genetic variants and outcome versus between genetic variants and exposure) | Figures | Main MR analyses visualised in Figures 3-5. |
| 12 | **Assessment of assumptions** |  |  |  |
|  | a) | Report the assessment of the validity of the assumptions | 18-21 | Sensitivity analysis results presented and in Supplementary Tables 10-30 |
|  | b) | Report any additional statistics (e.g., assessments of heterogeneity across genetic variants, such as *I^2^*, Q statistic or E-value) | Tables | Q statistics provided in Supplementary Table 31 |
| 13 | **Sensitivity analyses and additional analyses** |  |  |  |
|  | a) | Report any sensitivity analyses to assess the robustness of the main results to violations of the assumptions | 18-21 | Sensitivity analysis results presented and in Supplementary Tables 10-30 |
|  | b) | Report results from other sensitivity analyses or additional analyses | 18-21 | Sensitivity analysis results presented and in Supplementary Tables 10-30 |
|  | c) | Report any assessment of direction of causal relationship (e.g., bidirectional MR) | NA | Not provided |
|  | d) | When relevant, report and compare with estimates from non-MR analyses | 17-18 | Explicit comparison with multivariable linear MR provided and in Supplementary Tables 7-9 |
|  | e) | Consider additional plots to visualize results (e.g., leave-one-out analyses) | Figures | Leave-one-out plots provided in Supplementary Figure 3 |
|  | **DISCUSSION** |  |  |  |
| 14 | **Key results** | Summarize key results with reference to study objectives | 21-23 | Key findings provided with caution in causal interpretation |
| 15 | **Limitations** | Discuss limitations of the study, taking into account the validity of the IV assumptions, other sources of potential bias, and imprecision. Discuss both direction and magnitude of any potential bias and any efforts to address them | 25-27 | Comprehensive limitations provided |
| 16 | **Interpretation** |  |  |  |
|  | a) | Meaning: Give a cautious overall interpretation of results in the context of their limitations and in comparison with other studies | 22-25 | Caution in interpretation and comparisons with other studies provided |
|  | b) | Mechanism: Discuss underlying biological mechanisms that could drive a potential causal relationship between the investigated exposure and the outcome, and whether the gene-environment equivalence assumption is reasonable. Use causal language carefully, clarifying that IV estimates may provide causal effects only under certain assumptions | 23-25 | Mechanisms by which bias can be induced in analyses and thus caution being required in interpreting MR estimates is provided |
|  | c) | Clinical relevance: Discuss whether the results have clinical or public policy relevance, and to what extent they inform effect sizes of possible interventions | 28 | Relevance of sensitivity analyses and caution in interpreting results are provided in conclusive statement |
| 17 | **Generalizability** | Discuss the generalizability of the study results (a) to other populations, (b) across other exposure periods/timings, and (c) across other levels of exposure | 26-27 | Generalisability of studies used described |
|  | **OTHER INFORMATION** |  |  |  |
| 18 | **Funding** | Describe sources of funding and the role of funders in the present study and, if applicable, sources of funding for the databases and original study or studies on which the present study is based | 28 | Funding provided in “Acknowledgements” statement |
| 19 | **Data and data sharing** | Provide the data used to perform all analyses or report where and how the data can be accessed, and reference these sources in the article. Provide the statistical code needed to reproduce the results in the article, or report whether the code is publicly accessible and if so, where | 29 | Data availability statement provided |
| 20 | **Conflicts of Interest** | All authors should declare all potential conflicts of interest | 29 | Conflicts of interest provided in “Additional information” section |

This checklist is copyrighted by the Equator Network under the Creative Commons Attribution 3.0 Unported (CC BY 3.0) license.

1.	Skrivankova VW, Richmond RC, Woolf BAR, Yarmolinsky J, Davies NM, Swanson SA, et al. Strengthening the Reporting of Observational Studies in Epidemiology using Mendelian Randomization (STROBE-MR) Statement. JAMA. 2021;under review.

2.	Skrivankova VW, Richmond RC, Woolf BAR, Davies NM, Swanson SA, VanderWeele TJ, et al. Strengthening the Reporting of Observational Studies in Epidemiology using Mendelian Randomisation (STROBE-MR): Explanation and Elaboration. BMJ. 2021;375:n2233.
